# Reliability and Validity of Bifactor Models of Dimensional Psychopathology in Youth from three Continents

**DOI:** 10.1101/2021.06.27.21259601

**Authors:** Maurício Scopel Hoffmann, Tyler Maxwell Moore, Luiza Kvitko Axelrud, Nim Tottenham, Xi-Nian Zuo, Luis Augusto Rohde, Michael Peter Milham, Theodore Daniel Satterthwaite, Giovanni Abrahão Salum

## Abstract

Bifactor models are a promising strategy to parse general from specific aspects of psychopathology in youth. Currently, there are multiple configurations of bifactor models originating from different theoretical and empirical perspectives. Our aim is to identify and test the reliability, validity, measurement invariance, and the correlation of different bifactor models of psychopathology using the Child Behavior Checklist (CBCL). We used data from the Reproducible Brain Charts (RBC) initiative (N=7,011, ages 5 to 22 years, 40.2% females). Factor models were tested using the baseline data. To address our aim, we a) mapped the published bifactor models using the CBCL; b) tested their global model fit; c) calculated model-based reliability indices. d) tested associations with symptoms’ impact in everyday life; e) tested measurement invariance across many characteristics and f) analyzed the observed factor correlation across the models. We found 11 bifactor models ranging from 39 to 116 items. Their global model fit was broadly similar. Factor determinacy and H index were acceptable for the p-factors, internalizing, externalizing and somatic specific factors in most models. However, only p- and attention factors were predictors of symptoms’ impact in all models. Models were broadly invariant across different characteristics. P-factors were highly correlated across models (r = 0.88 to 0.99). Homotypic specific factors were also highly correlated. Regardless of item selection and strategy to compose CBCL bifactor models, results suggest that they all assess very similar constructs. Our results provide support for the robustness of the bifactor of psychopathology and distinct study characteristics.

**General Scientific Summaries:** This study supports the notion that models of psychopathology that separate what is general from what is specific in mental health problems have little impact from item selection and types of specific dimensions. The general dimensions are highly correlated among different models, valid to predict symptom impact in daily life and are not influenced by demographic and clinical characteristics, time and information.

---

Symptoms of mental health conditions in youth tend to co-occur and having one condition increases the chances of having another (Caspi et al., 2020; McGrath et al., 2020). This has led to the application of bifactor models in psychopathology (Caspi et al., 2014; Lahey et al., 2012), which can explicitly model what is shared (or general, transdiagnostic) and what are the unique (or specific) aspects of psychopathology. This type of modelling is advantageous because shared and unique dimensions of psychopathology can be linked with shared and unique biological factors (Caspi & Moffitt, 2018; Elliott et al., 2018; Grotzinger et al., 2020; Kaczkurkin et al., 2019; Romer et al., 2020; Sprooten et al., 2021; Xia et al., 2018) and this might result in better specificity when examining biological underpinnings of mental conditions in youth in comparison with correlated models. Whereas research has demonstrated the value of bifactor models, major issues in their implementation remain.

First, many different bifactor models of psychopathology have been suggested, even for the same questionnaire. Using this framework, the shared factor of psychopathology is often referred to as the “p-factor” (Caspi et al., 2014). However, the fact we name the shared latent factor as p-factor does not mean everything that comes out of bifactor models in a Confirmatory Factor Analysis (CFA) will have the same meaning irrespective of the theoretical or empirical model selection/generation. Therefore, the choice of the underlying theoretical or empirical model may have an impact on meaning of the shared and unique factors, that might in turn lead to inconsistent interpretations of these latent factors in the literature (Caspi & Moffitt, 2018; Eid et al., 2017; Smith et al., 2020; Watts et al., 2020).

Second, models might vary in their levels of reliability, i.e., in how cohesive are selected items for informing about shared and unique aspects of psychopathology. Indicator selection and model configuration (e.g., which and how many specific factors a bifactor model has) may have some impact on how they fit the data (i.e, global fit) and also on factor reliability, determinacy and how well the factors are defined by its items (i.e., model-based reliability). Specific factors of some models show low reliability and low determinacy, the latter indicating that the factor score is a poor representation of the modeled factor from which it was calculated (Grice, 2001). Indeed, it is possible for a score calculated from a poorly determined factor to correlate ∼0 (or even negatively) with the theoretical construct on which the factor is based, making any type of inference about a person’s relative standing on that factor difficult or impossible (Bornovalova et al., 2020; Forbes et al., 2021; Moore et al., 2020). Therefore, assessing how different models perform in terms of reliability might allow research to select models that are more likely to provide reliable information and prevent replication problems between studies. Nonetheless, p- and specific factors can simultaneously predict a given variable to investigate if each of these factors independently accounts for unique variance in the outcome variable (Moore et al., 2020). Thus, aside from reliability, factors derived from bifactor models should be able to explain external variables.

Third, it is not certain whether the models are comparable (i.e., invariant) across distinct sample characteristics, often being a neglected indicator of a model’s quality and utility. Published bifactor models using the Child Behavior Checklist (CBCL), a widely used and extensive assessment of multiple signs and symptoms in youth, have shown that mean level differences of the eight-syndrome model are comparable across 30 countries (Ivanova et al., 2007). Nonetheless, we do not know if CBCL models are invariant according to other characteristics while using the bifactor modelling. This would inform whether CBCL score differences are given by changes in the general and/or specific factors only, not by other potential sources of variation, such as study site, but also age, gender, race/ethnicity, intelligence, educational level, mental health condition status and informant. For that purpose, data sets containing harmonized individual and contextual characteristics could be very useful. Testing bifactor models in harmonized data sets could help understand which model holds comparability across groups of different characteristics, informants, and stages of human development.

Finally, the extent to which existing models compare to one another and inform us about the same latent factors is inconsistent. There is some important debate on whether bifactor models measured the same construct in a context of varying indicator selection (Bornovalova et al., 2020; Clark et al., 2021; Watts et al., 2020). The selection of items that compose a model is frequently theory- or empirically-driven. Applied to CBCL, a recent study tested four bifactor models derived from theoretical and empirical indicator selection approaches and found that the p-factors, internalizing and externalizing specific factors were highly correlated (Clark et al., 2021). However, another study reviewed models from previous literature and found that bifactor models are highly sensitive to indicator selection (Watts et al., 2020). Thus, comparability between previous studies would benefit from an investigation on whether their factors are correlated, despite distinct item selection and model configuration.

The understanding of what the bifactor model of psychopathology represents must go through the identification of the different published models, the estimation of their global fit, model-based reliability and criterion validity, whether the models are influenced by other sample characteristics and whether the factors derived from different models inform a similar construct. This would ultimately help to elucidate whether different models represent similar or distinct constructs while reliably representing psychopathology. To answer these questions, the present study aimed to compile published bifactor models of psychopathology originating from different methodologies and compare them in their global model fit, model-based reliability, invariance and correlations among the constructs of these models. Doing so allowed us to understand the extent to which the model specifications and item selection impacted model quality, and if the models are distinguishable, to select the best model to represent the bifactor model of psychopathology. To address our aims, we have, a) reviewed the literature and mapped the published item-level bifactor models using the CBCL, b) tested models’ global fit and c) model-based reliability indices, d) tested criterion validity by means of each factor’s association with symptom impact in everyday life; e) measured model invariance, and f) analyzed factor correlation across the models. Based on previous studies, we hypothesized that models will have similar global fit, specific factors will have low reliability, they will be broadly invariant, and p-factors as well as homotypic specific factors will be moderately to highly correlated.

## Methods

### Sample

The sample was obtained from the Reproducible Brain Charts (RBC) initiative. The aim of the initiative is to harmonize phenotypic and neuroimaging data to build highly reproducible growth charts of youth brain development. It contains harmonized demographic, educational, cognitive and clinical data across samples from eight large-scale developmental imaging cohorts. The studies composing the RBC are the Philadelphia Neurodevelopmental Cohort (PNC) (Satterthwaite et al., 2016), the Brazilian High-Risk Cohort Study for Mental Conditions (BHRCS) (Salum et al., 2015), the Healthy Brain Network (HBN) (Alexander et al., 2017), the Nathan Kline Institute-Rockland Sample (NKI-RS) (Nooner et al., 2012), the developmental component of the Chinese Color Nest Project (devCCNP) (Liu et al., 2020), the Parents and Children Coming Together (PACCT; PIs: Tottenham & Milham), the Human Connectome Project-Development (HCP-D) (Somerville et al., 2018), and the Pediatric Neuroimaging and Genetics Study (PING) (Jernigan et al., 2016). For the purposes of the present study, we have included five studies that used the CBCL (BHRCS, HBN, NKI, devCCNP and PACCT, baseline N=7,011, aged 5 to 22 years, 40.2% females). Three studies have prospective CBCL data (BHRCS, NKI and devCCNP) which were used to assess time invariance testing (baseline n=3,066, aged 5 to 18, 45.6% females; second follow-up n=2,154, aged 7 to 23, 45.8% females). Characteristics of each study sample and the total sample are described in Table S1.

### CBCL

CBCL is a parent-reported assessment of 120 emotional and behavioral symptoms in subjects aged 6 to 18 over the past 6 months, answered in a 3-point scale (0 = not true; 1 = somewhat/sometimes true; 2 = very true/often). It was developed and is applied within the Achenbach System of Empirically Based Assessment (ASEBA), which aggregate the scores in eight syndromes (anxious-depressed, withdrawn-depressed, somatic complaints, rule-breaking behavior, aggressive behavior, social problems, thought problems, attention problems). Another operationalization is to combine the scores of anxious-depressed, withdrawn-depressed and somatic complaints into an internalizing problems score, and rule-breaking behavior and aggressive behavior into an externalizing problems score (Achenbach & Rescorla, 2001). In studies with low sample sizes, a few items were not scored as “very true/often”. Therefore, items were rescored to indicate the presence of symptoms (i.e., symptoms rated as 1 or 2 were coded as 1), which is in line with previous item-level analyses of the CBCL (Achenbach & Rescorla, 2001; Deutz et al., 2020; McElroy et al., 2018). ASEBA’s Youth Self Report (YSR) and Teacher Report Form (TRF) were also available in HBN and were used to assess informant invariance (100 compatible items across all instruments) (Achenbach & Rescorla, 2001).

### Symptom impact in daily life

Symptom impact was measured using the Strengths and Difficulties Questionnaire (SDQ) impact questions. Parents were asked whether they think that their child has difficulties in emotions, concentration, behavior or being able to get on with other people. If they have, they were further asked whether these difficulties a) cause distress, b) interfere in their everyday life regarding family life, c) friendships, d) learning and e) leisure activities. For HBN, answers were 0=Not at all/Only a little, 1=A medium amount, 2=A great deal. For the BHRCS, options were 0=Not at all, 1= Only a little, 2=A lot, 3=More than a lot. A previous study demonstrated good internal and external validity of this assessment (Stringaris & Goodman, 2013). We estimated impact using a unidimensional solution as previously described using confirmatory factor analysis (CFA) with the same specifications as described in the statistical analysis section for the CBCL.

### Grouping variables

Subjects were classified in their race/ethnicity groups as Asian, black, mixed, native, white, other and unknown. For the measurement invariance purposes, race/ethnic groups were classified as white and non-white. Educational attainment was assessed in years of schooling and classified if enrolled in primary (currently) or secondary school (current or completed). Intelligence quotient (IQ) was assessed using the Wechsler Abbreviated Scale of Intelligence II (NKI), Wechsler Intelligence Scale for Children IV (HBN) and III (BHRCS) and, for those aged 18 and older, the Wechsler Adult Intelligence Scale IV (HBN) (Grizzle, 2011; Wechsler, 1999, 2002, 2008). IQ was standardized for age and gender within each study sample.

Current mental health condition status (present vs. not present) was evaluated according to each study methodology, all generating DSM-IV-based diagnosis. Briefly, HBN used the Kiddie Schedule for Affective Disorders and Schizophrenia computerized web-based clinical-consensus version (K-SADS-COMP). NKI used the present and lifetime version of the same instrument (K-SADS-PL) (Kaufman et al., 1997). BHRCS applied clinical consensus rating based on diagnostic probabilities obtained with the Development and Well-being Assessment (Goodman et al., 2000). The other two studies (PACCT and devCCNP) did not present diagnostic, educational, or cognitive information. PACCT did not present information on age and gender and was further excluded from the invariance testing analysis.

### Literature review on CBCL bifactor models

The review was anchored on a working paper by Constantinou and Fonagy (2019), which is a methodological review on the reliability of bifactor model estimates for psychopathology. They found 49 studies using different instruments, age-range and methods. We built on these findings and, using their search terms ((bifactor OR bi-factor OR “nested factor” OR “p factor”) AND (psychopathology OR psychiatr* OR disorder OR symptom OR diagnosis OR mental health)) added terms for our target population (AND (child* or adolesc* or youth or young)). Search was carried out using Pubmed and Google Scholar, restricting for publications from 2019 up until July 2020. For the purposes of the present study, we selected item-level CBCL models and samples that included children and adolescents.

## Data analysis

### Global fit and model-based reliability testing

The CBCL bifactor models were estimated with CFA. In a bifactor model, all items of CBCL were configured to load on a general factor of psychopathology, at the same time that some of the items were residually loaded by specific factors, depending on the model specification mentioned in Figure 1. Specific factors were not allowed to correlate between each other, nor with the general factor. CFA was carried out using delta parameterization and weighted least squares with diagonal weight matrix with standard errors and mean- and variance-adjusted chi-square test statistics (WLSMV) estimators. To evaluate model fit, we used root mean square error of approximation (RMSEA), comparative fit index (CFI), Tucker–Lewis index (TLI) and standardized root mean-square residual (SRMR). Values of RMSEA lower than 0.060 and CFI or TLI values higher than 0.950 indicate a good-to-excellent model (Hu & Bentler, 1999). SRMR lower or equal than 0.100 indicate adequate fit, and lower than 0.060 in combination with previous indices indicate good fit (Hu & Bentler, 1999; Kline, 2015).

**Figure1:**
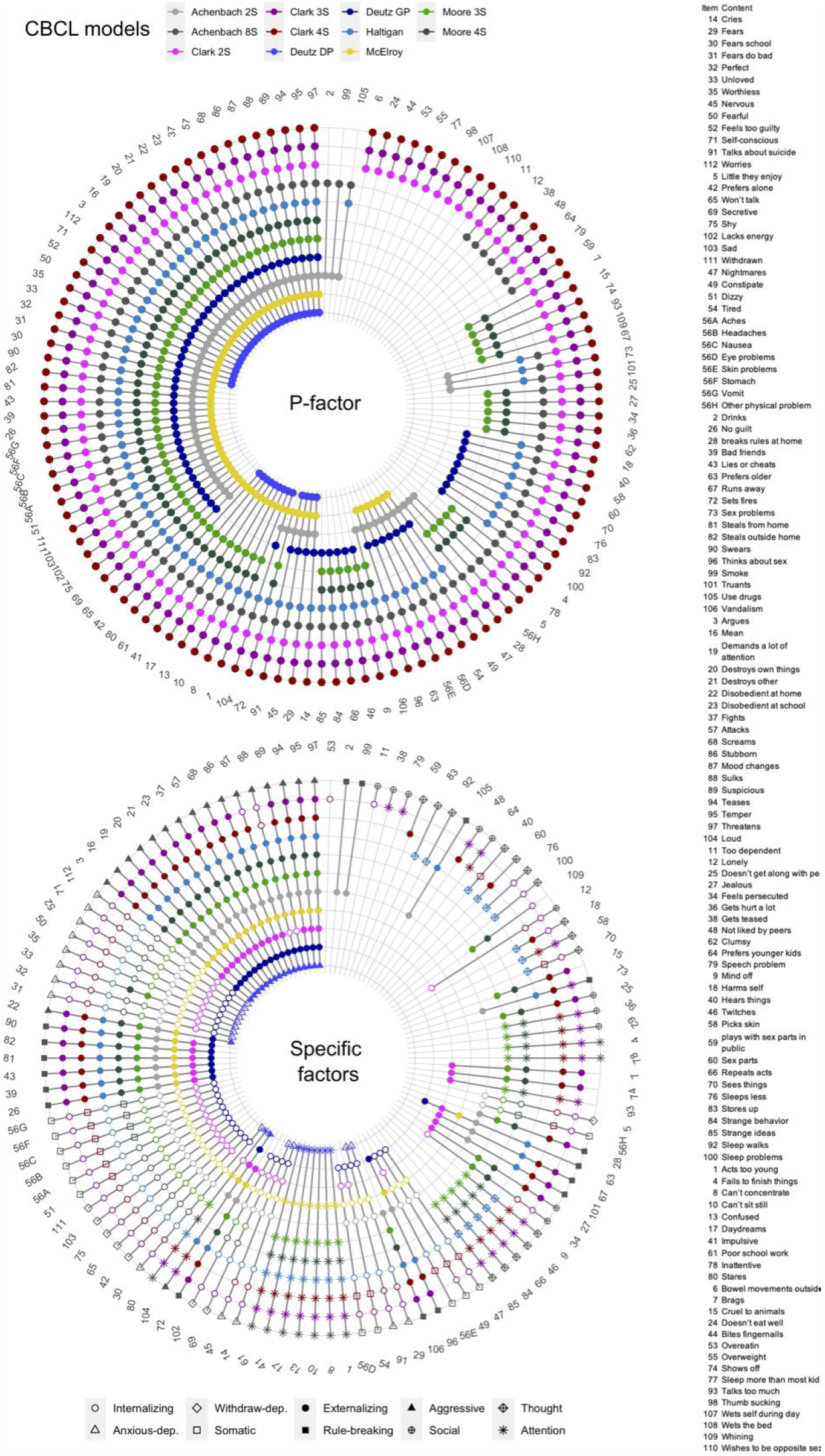
CBCL items included in each model. Items included in the p-factor are depicted in the upper plot and in the specific factors in the bottom. nS, number of specific factors in the model; p-factor, the general factor of psychopathology; GP, General psychopathology model; DP, dysregulation profile model; CBCL, Child and Behavior Checklist. * this model was first described by Deutz et. al. (2018) using the Strengths and Difficulties questionnaire and further tested by Haltigan et. al. in 2018 and Deutz et. al. in 2020.

We used 10 model-based reliability indices to evaluate the bifactor models, which are described in full in supplemental material (page 3). Briefly, they were omega (ω), hierarchical omega (ωH), factor determinacy (FD), H index, explained common variance (ECV), ECV of a specific factor due to itself (ECV-SS), ECV of a specific factor with respect to the general factor (ECV SG), ECV of the general factor with respect to a specific factor (ECV GS), percent uncontaminated correlations (PUC) and item explained common variance (IECV) (Dueber, 2017; Rodriguez et al., 2016). When ωH is > 0.8 and ECV and PUC are > 0.7, the construct can be interpreted as unidimensional (Rodriguez et al., 2016). We have also included the percentage of items on specific factors with negative factor loading (PINFL) as an index. Negative factor loadings can be an indicator of anomalous results in the bifactor model because either, the negative correlations among some set of items are strong enough to cause the formation of their own “contrast” factor defined by said items loading in opposite directions, or more commonly, the item variance is being completely captured by the general factor and the contribution of the specific factor is therefore null to explain item variance (Eid et al., 2017).

### Factor correlation

We estimated the factor correlation across the CBCL models to further understand if and to what extent the models are convergent to inform general and specific factors. To that purpose, we extracted the factor scores from the CBCL models and plotted the Pearson correlation coefficients and estimated the 95% confidence intervals.

### Criterion validity

To test criterion validity, we estimated the SDQ symptom impact using CFA. After estimating this model, we combined it with each CBCL model while regressing impact on each CBCL factor, adjusting for age, gender and IQ, in a structural equation model (SEM), so the associations between CBCL factors and impact are not vulnerable to the hazards of factor score calculation (Forbes et al., 2021; Grice, 2001; Kline, 2015). These SEMs were estimated for each CBCL model in BHRC and HBN samples separately (22 SEMs). We performed supplementary generalized additive models to test if between-model regression coefficient differences of impact latent variable on p- and paired specific factors was associated with the between-model 1) factors scores correlation and 2) the number and proportion of shared items (supplemental material, page 4).

### Measurement invariance

Invariance testing allows us to understand if the mean differences of a given test/questionnaire score across different groups are due to true differences in the mean levels of the latent construct which the test/questionnaire aims to assess (Meredith, 1993). In other words, it provides information about whether score differences are given by differences in the latent construct only and not influenced by exogenous sources of variation. In this sense, a measurement invariance violation is simply unmodeled multidimensionality in the data.

We tested if the CBCL models are invariant in terms of the following groups: 1) Age group (5 to 10 years old compared with 11 years old or older), 2) Gender (males and females), 3) Study site (two sites in the BHRCS and two sites in the HBN, which have a sample size to hold a 5:1 person/item ratio), 4) Race/ethnicity (white and non-white), 5) Educational level (primary versus at least some secondary school), 6) IQ (< 90, 90 to 109 and >109, considered below average, on average and above average) (Sattler, 2008), 7) Any psychiatric condition status (absent and present), 8) informant (parent-, self- and teacher-report) and 9) longitudinal invariance (three time-points). We included only cases with available data in each grouping variable for measurement invariance testing (see Table S1).

Invariance was tested using multigroup CFA (MG-CFA). It consists of applying a sequence of constraints and global model fit indices are compared between each constrained model. MG-CFA uses fixed-effects two parameters (thresholds and loadings) probit model, where configuration (configural invariance) and model parameters (scalar invariance) were not allowed to vary between groups (B. Muthén & Asparouhov, 2002; Ploubidis et al., 2019). Therefore, we tested if the models in each group are structurally similar (configural invariance) and if items are equally correlated with the latent factors and informing symptoms at equivalent level (scalar invariance). ΔCFI < 0.01 and ΔRMSEA < 0.015 or ΔSRMR < 0.010 between models with increasing levels of constraints indicate invariance (configural vs. scalar). If invariant, mean level differences among groups are due to differences in the latent trait (i.e., psychopathology) and not due to other sources of variation (Chen, 2007; Svetina et al., 2020).

All CFA and measurement invariance were carried out using Mplus version 8.6 (L. K. Muthén & Muthén, 2017) and implemented in R version 4.0.3 using the *MplusAutomation* package (Hallquist & Wiley, 2018), which was also used to extract factor scores generated in Mplus. For reproducibility in multiple data analysis languages, coding for the CFA was also implemented in R using the *lavaan* package (Rosseel et al., 2018). Notably, these approaches produce equivalent output (Narayanan, 2012). All bifactor reliability indices were calculated using the *BifactorIndicesCalculator* package in R (Dueber, 2017). Pearson correlation was estimated and plotted using the *rcorr* function in the Hmisc package (Harrell, 2021) and 95% confidence intervals using the cor.mtest function in the *corrplot* package (Wei et al., 2017). Code and supplemental tables can be found in https://osf.io/uwy5n/.

## Results

### Literature review on CBCL bifactor models

Based on the selection criteria described above, we found 283 studies, in which 81 presented non-duplicated relevant titles for abstract review (49 from Constantinou and Fonagy 2019, 26 from PubMed and 6 from Google Scholar). Five studies were included (Figure S1), for a total of nine selected models. We have further included two bifactor model versions of the two-syndrome and eight-syndrome models as recommended by the ASEBA (Achenbach & Rescorla, 2001). Therefore, we have estimated a total of 11 bifactor models (Figure 1), which were originated by different research groups and samples, and used different item selection approaches, ranging from 66 to 116 items. Henceforth, we name the models before the first author of each publication, specifying the number of specific factors (i.e., 2S, 3S, 4S, in which S stands for specific) (Achenbach & Rescorla, 2001; Clark et al., 2021; Deutz et al., 2020; Haltigan et al., 2018; McElroy et al., 2018; Moore et al., 2020). Full details of model structure and item loadings can be found in the appendix (Tables S2 to S12).

### Global fit

In the present analysis, the bifactor models presented acceptable to good fit to the data (Table 1). The Clark 3S and 4S models presented the best RMSEA index and acceptable CFI and TLI indices, and the Deutz-Haltigan DP model presented the best SRMR, CFI and TLI indexes. Clark 2S and 4S models present nonnegative loadings in the specific factors. The Clark 2S model is a derivative from the Achenbach 2S model, in which the general factor also loads on other items that are not included in the specific internalizing and externalizing factor. This is named a S-1 model (i.e., as if one specific factor is missing to explain a subset of item variance), in contrast to models in which all items load on the specific factors as well as the general factor (i.e., symmetric bifactor model).^18,19^ Therefore, all Clark models and the Deutz GP model are considered a S-1 bifactor model.

**Table 1.**
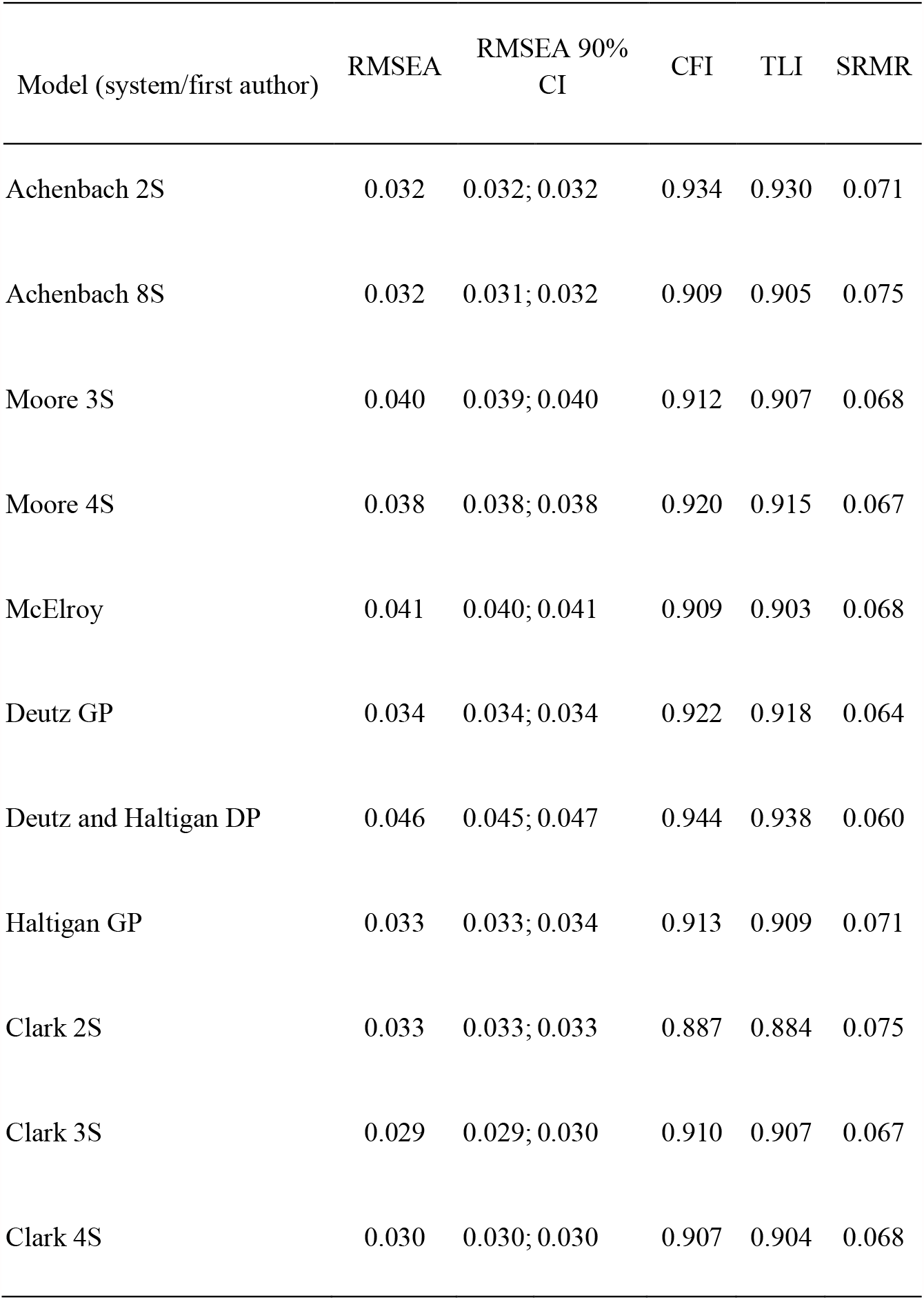
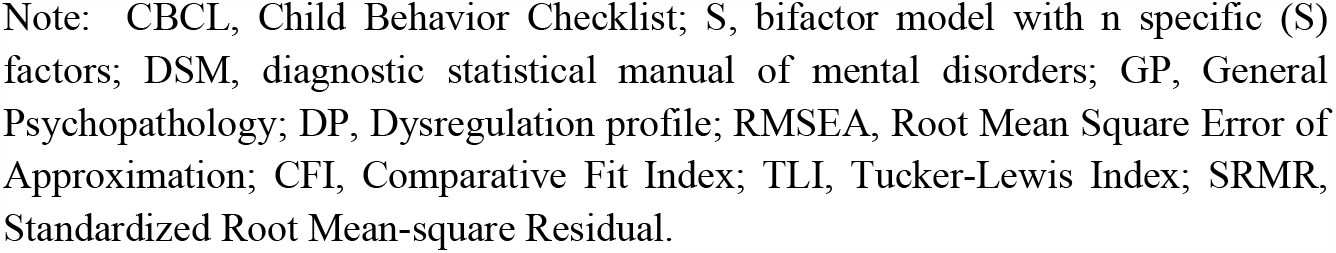
Global fit indices of CBCL models

### Model-based reliability indices

Table 2 shows complete model-based reliability results. P-factors from all models presented high reliability (mean ω = 0.952, range 0.935 to 0.962), H index (mean H = 0.976, range 0.967 to 0.983), FD (mean FD = 0.978, range 0.970 to 0.988) and represents 68.3% of the ECV (60.6% to 79.7%). The specific factors figures vary in a wider range. Reliability were generally low (mean ωH = 0.253, range 0.030 to 0.492), H index were acceptable in some cases (mean H = 0.797, range 0.584 to 0.919), as well as FD (0.915, range 0.814 to 0.951) and represents low ECV in relation to the general factor (mean ECV-SG = 9.2%, range 1.8% to 24.3%). Three models reached the minimum FD and H index threshold to consider all factors as well-defined (Achenbach 2S, Deutz GP and Clark 2S). Other models had at least one specific factor with H and FD indices below acceptable threshold.

**Table 2.**
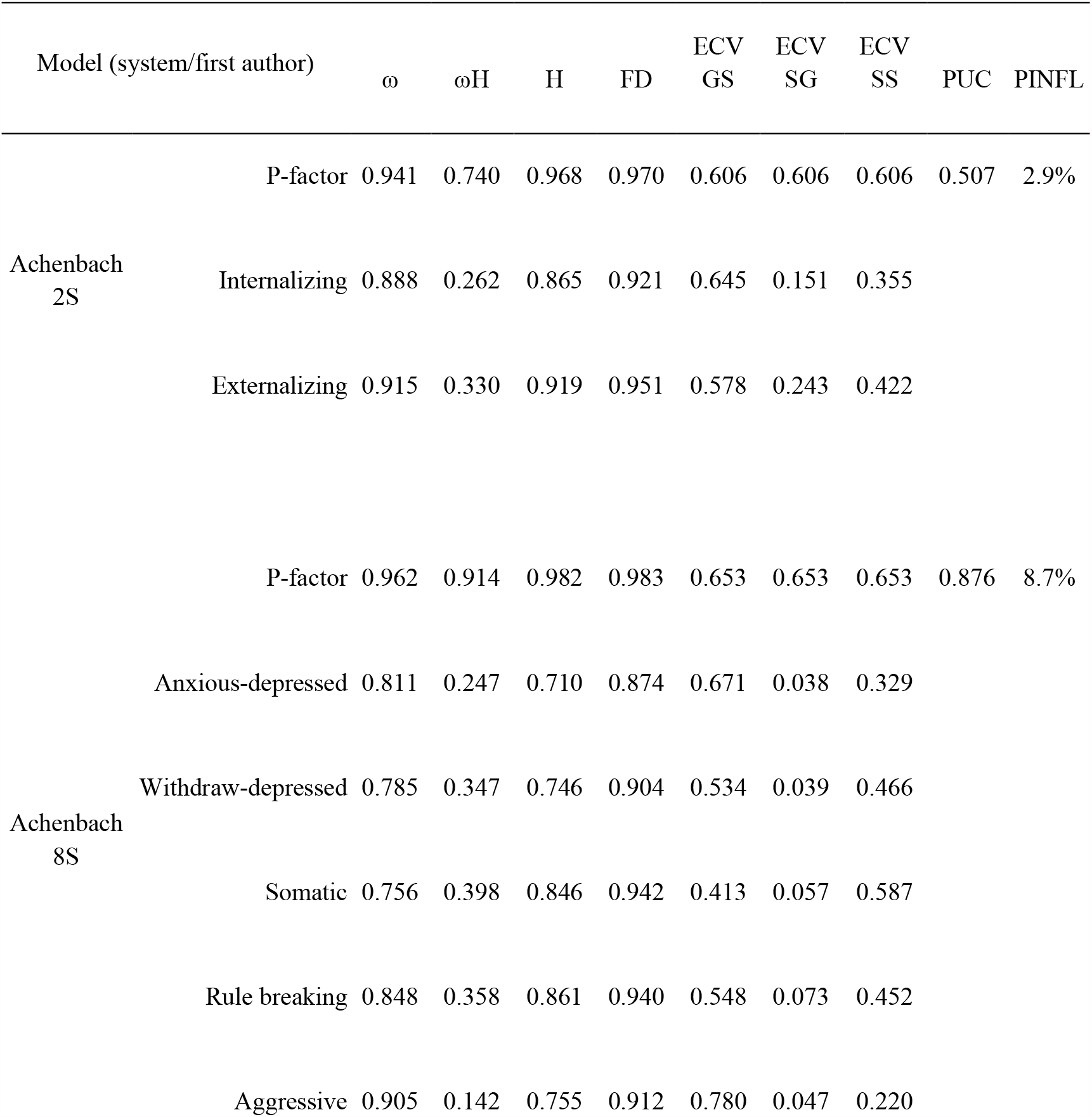
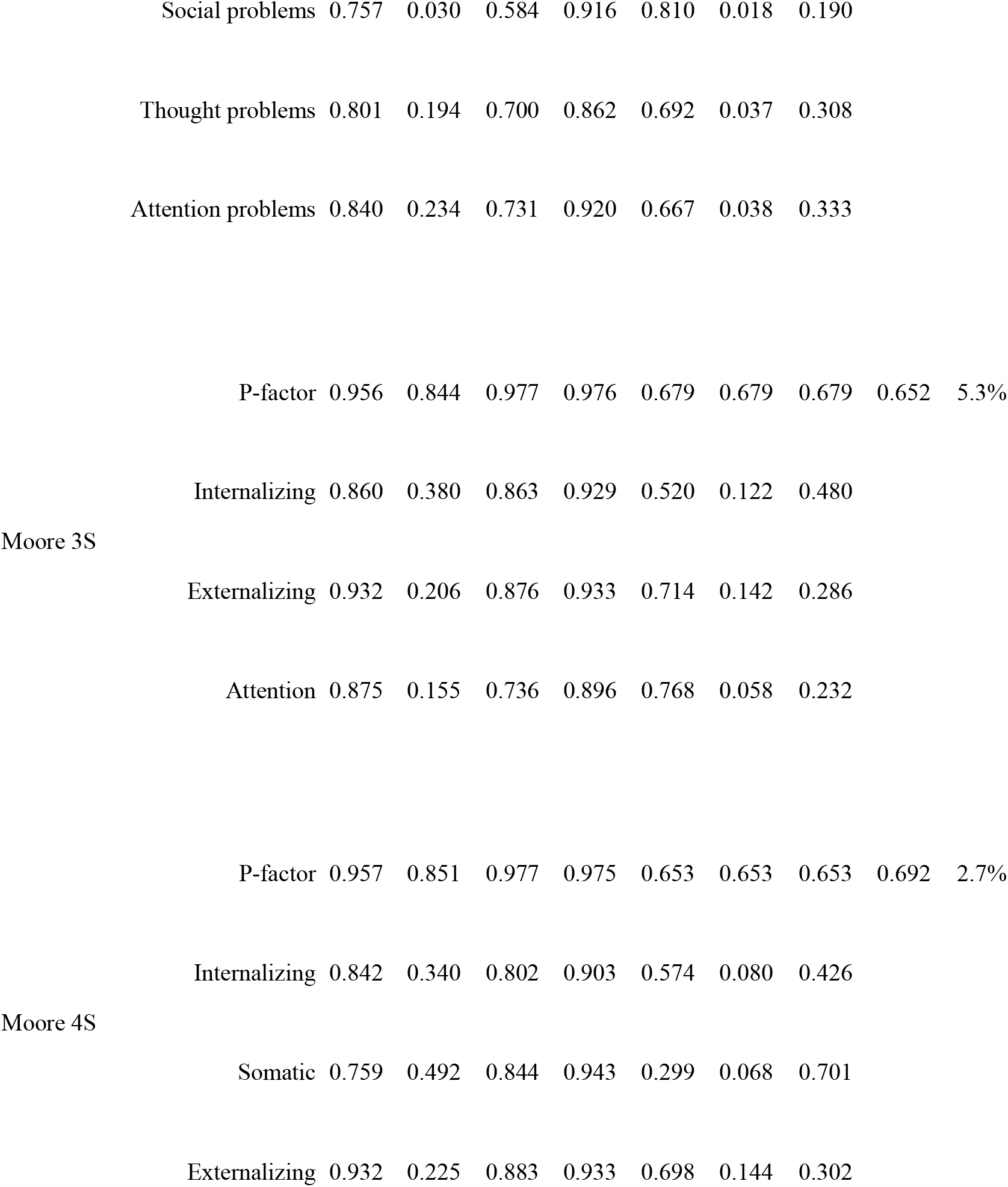
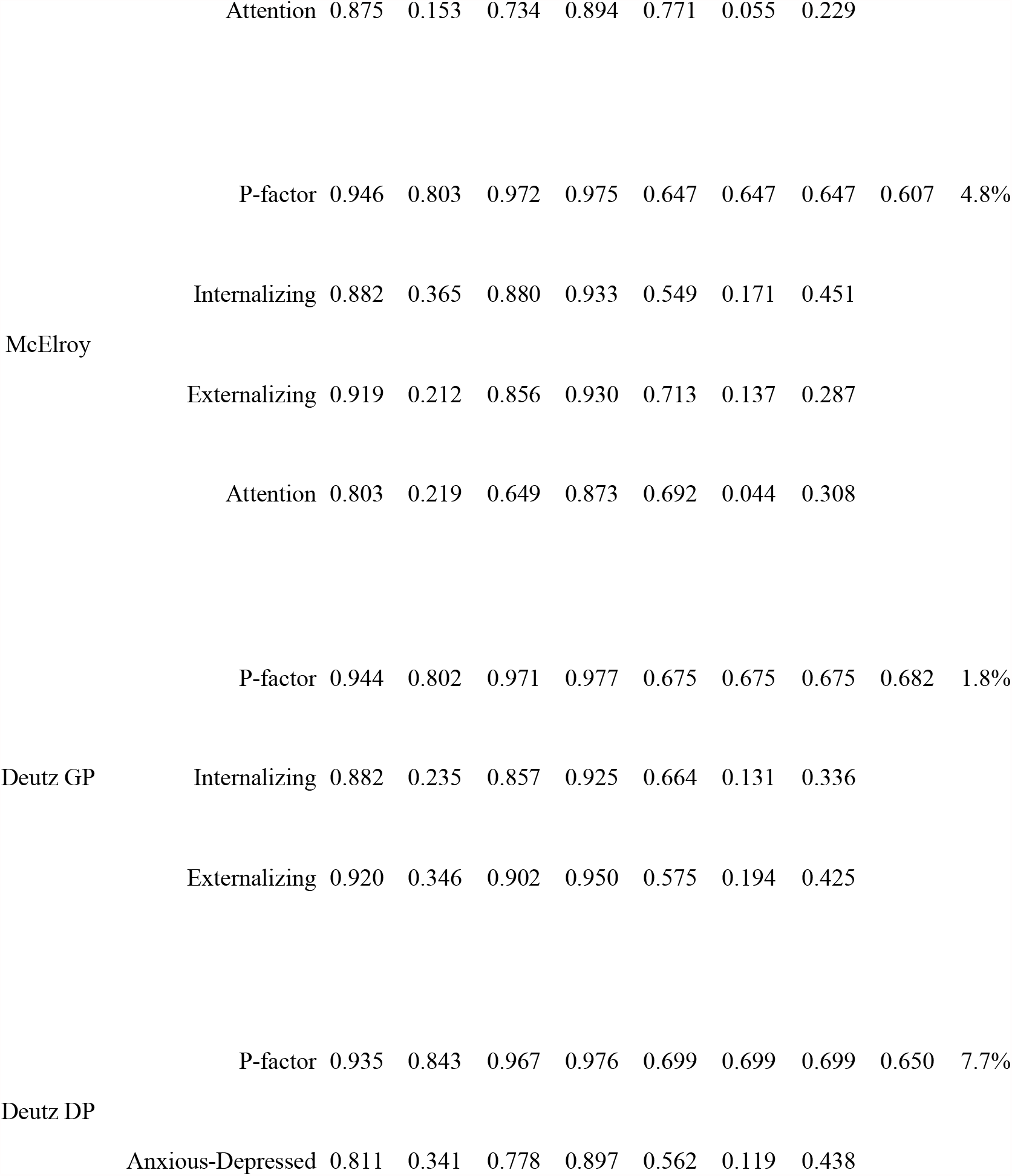
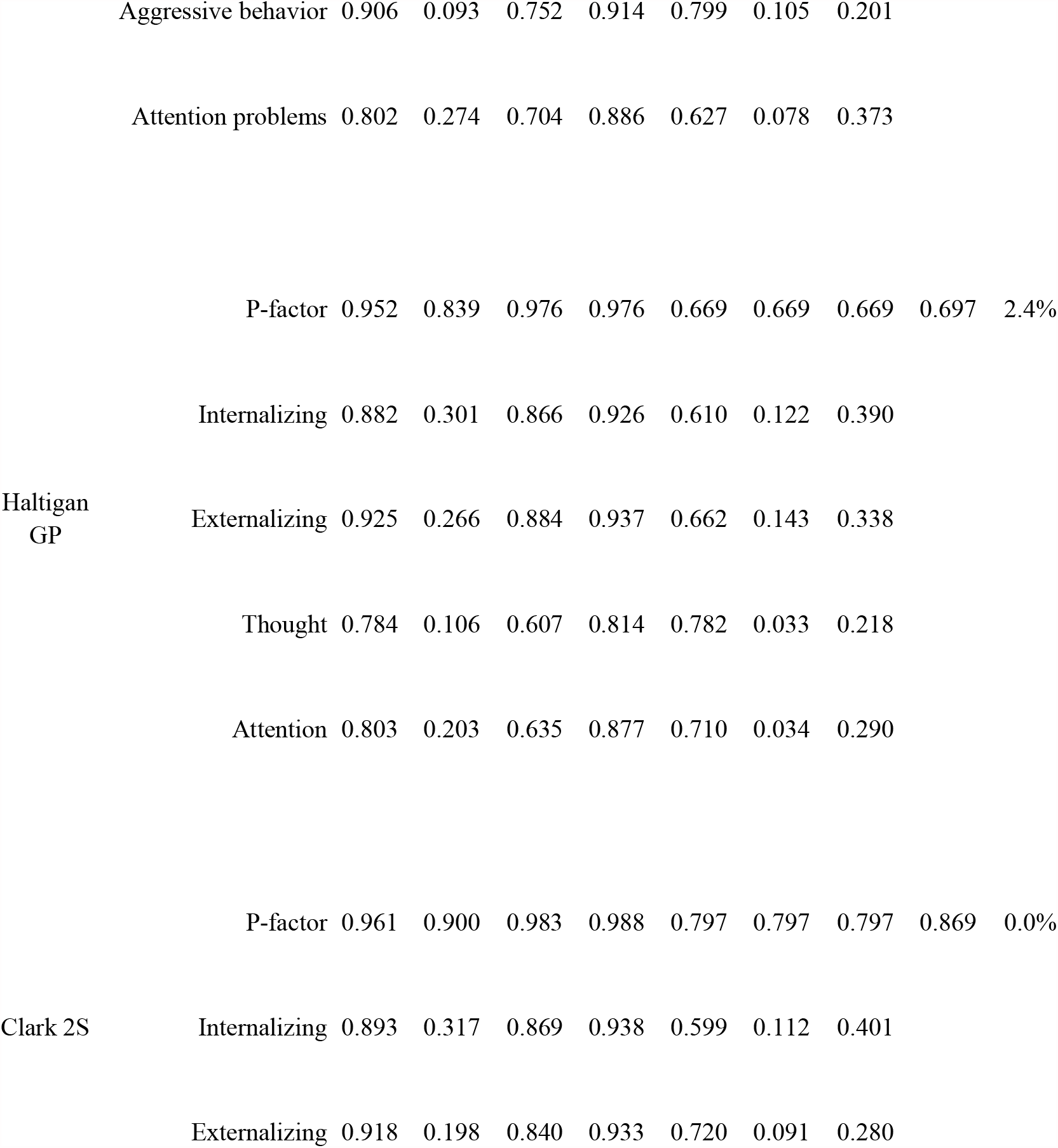
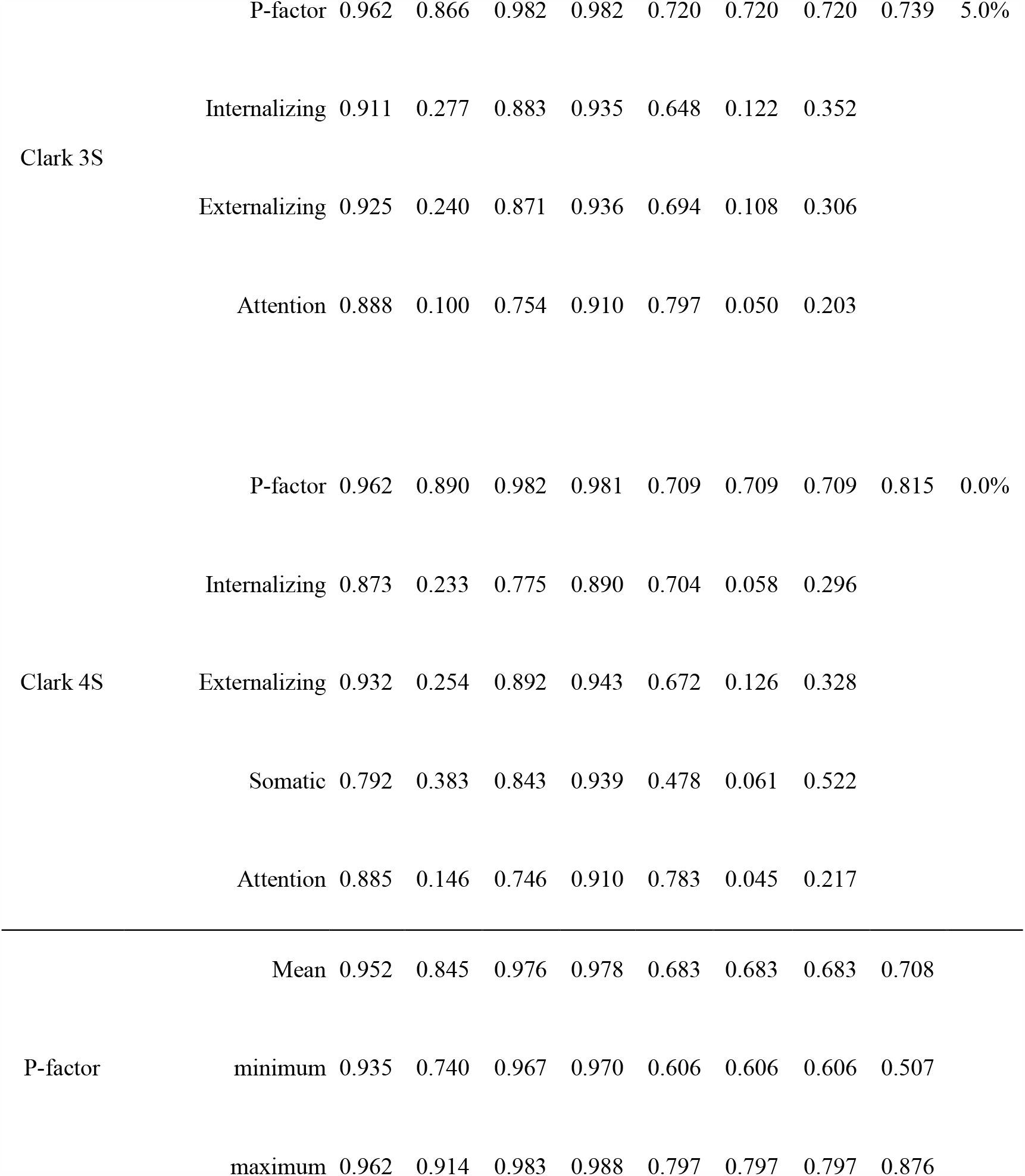
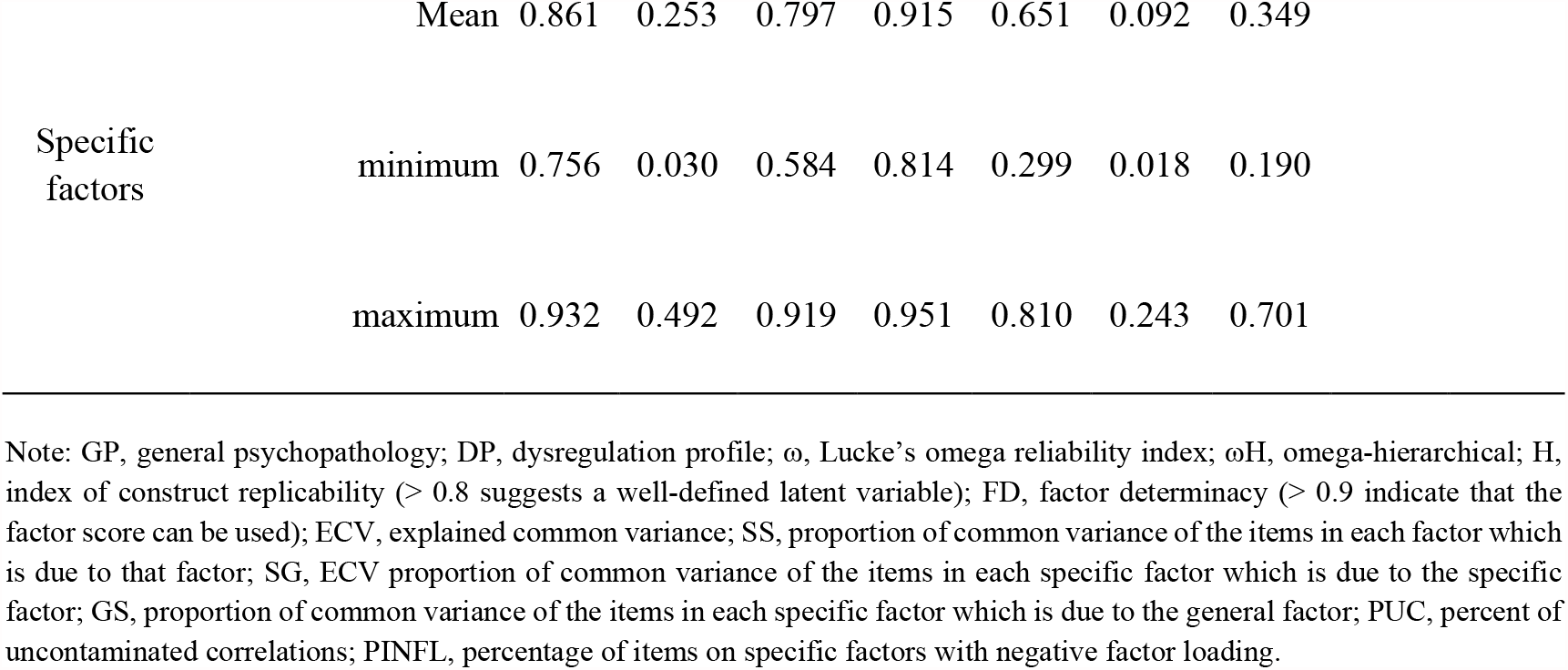
CBCL bifactor model-based reliability indices

Notably, IECV indices demonstrated that, when present in the model, factor loadings of items regarding “mood changes”, “suspicious”, “sulks”, “harms self”, “strange ideas” and “talks too much” tend to collapse in the specific factors and are good indicators of the p-factor across the models (Table S2 to Table S12).

### Criterion validity

Impact models fitted the data well (Table S13) and SEMs of impact regressed on CBCL models and covariates also presented acceptable global fit (Table S14). Impact was predicted by the p-factors of all models in both HBN and BHRCS (Figure 2). Aside from attention, the remaining specific factors were inconsistently related to daily life symptom impact, depending on the model or sample. All specific factors of the Moore 3S, McElroy, Deutz GP, Deutz-Haltigan DP, Clark 3S and Clark 4S were associated with impact in both tested samples. Complete latent regression results can be found in Table S15.

**Figure 2:**
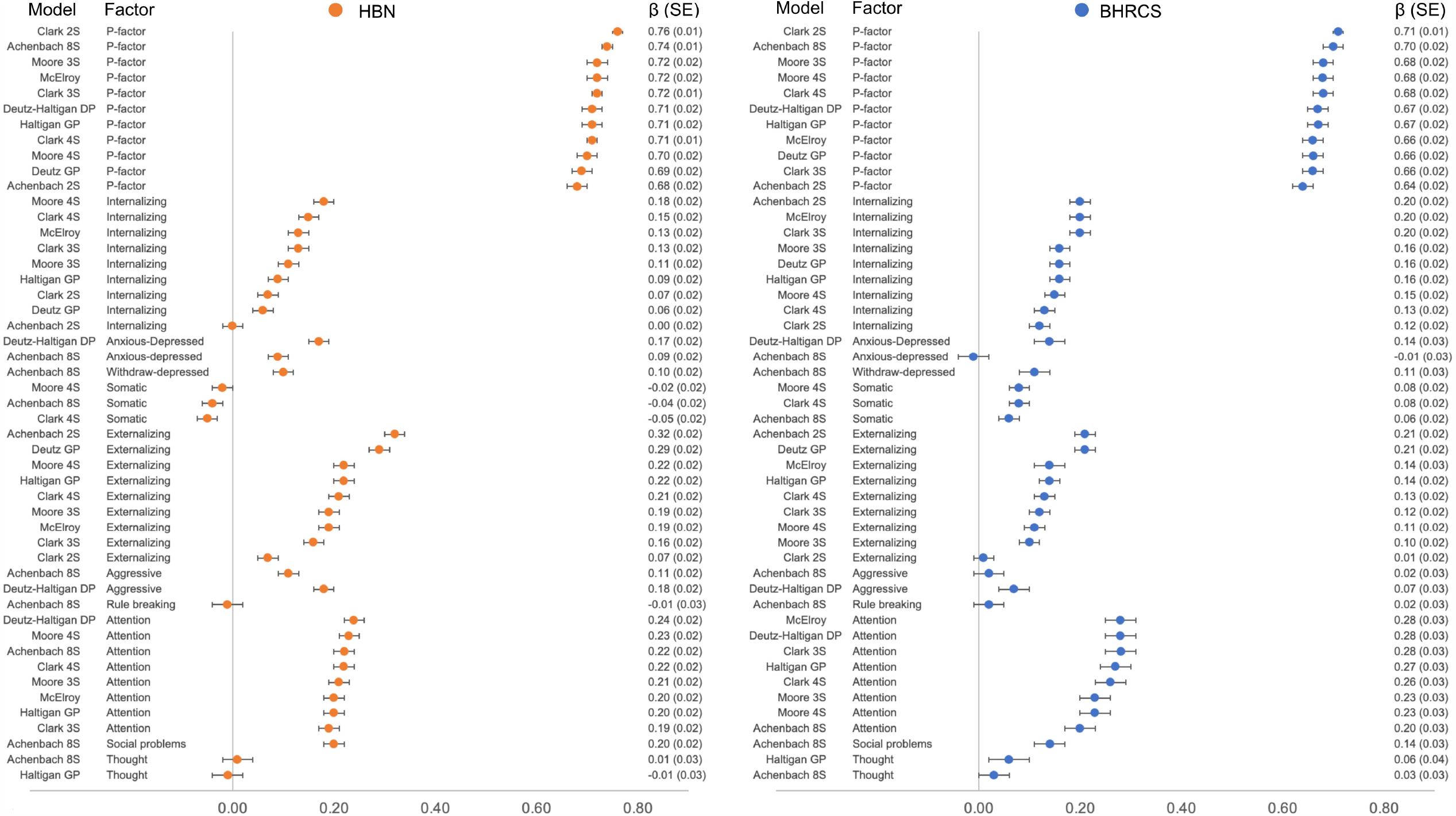
Latent regression models of symptom impact (SDQ) predicted by factors from each CBCL model. Regression models were carried out using structural equations where symptom impact factor was regressed on latent factors of each CBCL model plus age, gender and intelligence (one structural model for each CBCL model in each sample separately). β, standardized latent regression coefficient; SE, standard error. CBCL, Child Behavior Checklist; SDQ, Strengths and Difficulties Questionnaire; BHRCS, Brazilian High-Risk Cohort Study; HBN, Healthy Brain Network; S, bifactor model with n specific (S) factors; GP, General Psychopathology; DP, Dysregulation profile

Supplementary analysis (supplemental material, page 4) revealed that the differences of psychopathology latent factors with the external validator are a function of the factor scores correlation between models (Figure S2). Interestingly, the number and proportion of p-factor shared items between models are not associated with the differences of psychopathology latent factors with the external validator. The same does not hold for the specific factors (Figure S3).

### Invariance testing

Measurement invariance results are presented in Figure 3 and specific quantification and statistics can be seen in Tables S16 to S24. The models are broadly invariant across age groups, gender, study site, race/ethnicity, educational level, IQ, psychiatric condition status, informant and time-points. Therefore, for all models, a change of CBCL score was due solely to changes on the latent factor (i.e. p-factor, internalizing, etc.) and groups with the same levels (or changes) in the latent factor will present the same score on the observed indicators.

**Figure 3:**
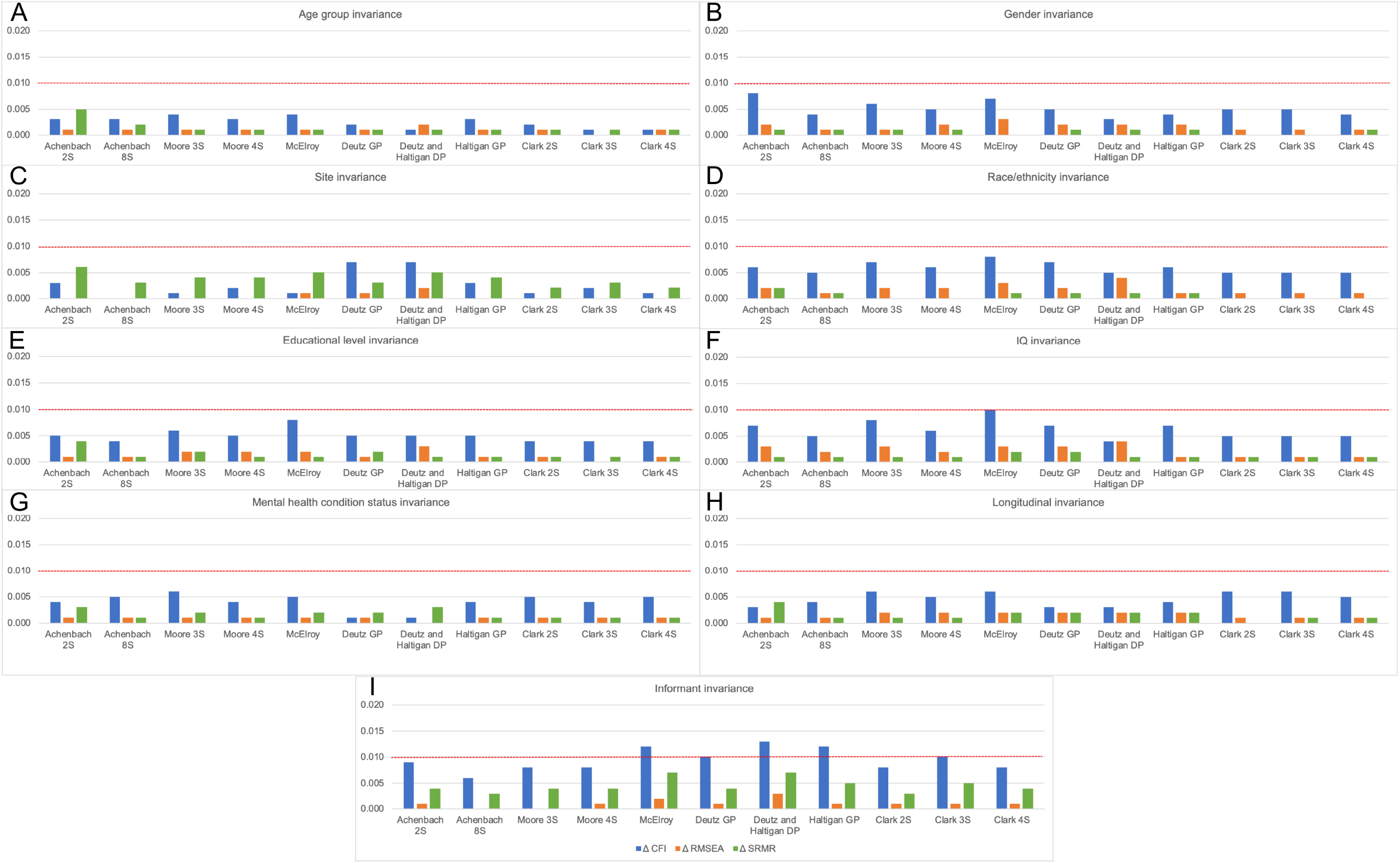
Y-axis represent the model fit differences between configural and scalar models (Δ) and X-axis represent each model. Dashed red line represents the threshold difference of 0.01. Blue bars represent Δ CFI, orange bars represent ΔRMSEA and green bars represent ΔSRMR. A) Age group (5 to 10 years old compared with 11 years old or older); B) Gender (males and females); C) Study site (two sites in the BHRCS study and two sites in the HBN study); D) Race/ethnicity (white and non-white); E) Educational level (primary and at least on the secondary school); F) IQ (below average, average and above average); G) Mental health condition status (absent and present); H) longitudinal invariance (three time-points) and I) Informant (parent-, self- and teacher-report) . CBCL, Child Behavior Checklist

### Factor correlation

Figure 4 depicts high correlation of the p-factor (Figure 4A) across the 11 models. Correlations between heterotypic specific factors are low, as expected when the general factor is taken into account, and high in homotypic factors between different models (Figure 4 B-D). Correlation coefficients, p-values and 95% confidence intervals between general and specific factors are shown in supplementary Tables S25 and S26 respectively.

**Figure 4:**
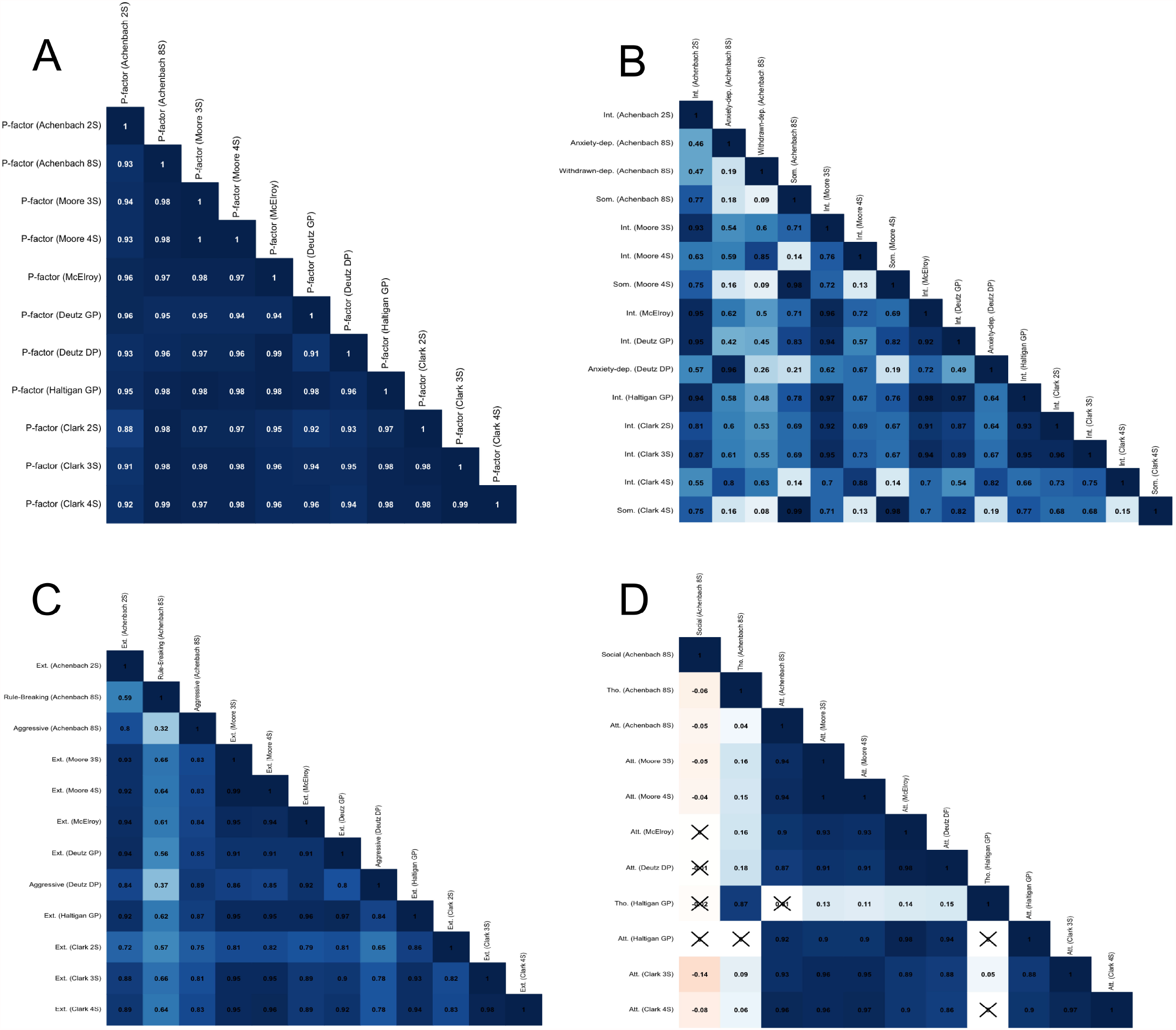
Correlation among p-factors (A), internalizing and somatic-related (B), externalizing-related (C) and attentional, thought and social specific factors (D). P-factor, the general factor of each model; nS, the number of specific factors in the model; GP, general psychopathology model; DP, dysregulation profile model; Int., internalizing; Som., somatic; -dep., depression; Ext., externalizing; Att., attention; Tho., thought.

## Discussion

Bifactor models of the CBCL, the most used instrument to measure dimensional psychopathology in youth, are a promising approach to improve the specificity of findings in the field of child and adolescent psychiatry. In this study, we searched for published item-level CBCL-based bifactor models of psychopathology, aiming to test their properties and advantages. We found 11 variations of CBCL bifactor models, using different numbers of items and both theory- and empirically-driven item selection and clustering procedures. Consistent with our hypothesis, the 11 models had similar global fit and invariance across different sample characteristics. However, reliability varied among the CBCL bifactor models. Aside from the p-factor, models with internalizing, externalizing and somatic specific factors and with less specific factors tend to produce acceptable H index and FD. P-factors and homotypic specific factors were found to be highly correlated regardless of the modelling specifications.

The interpretation of bifactor models of psychopathology have been controversial. It has been linked with non-specific liability to mental health problems, cognitive dysfunction, emotional dysregulation, psychosis, simply comorbidity or even an artifact (Achenbach, 2021; Caspi & Moffitt, 2018; Fried et al., 2021; Lahey et al., 2017, 2021; Smith et al., 2020; Watts et al., 2020). Nonetheless, to understand the meaning of these models, it is important to examine if the model’s configuration has any impact on the p-factor or specific factors. Previous studies have used different approaches to select items and specific factors to generate their model configuration (theory, researcher selection, data-driven, etc.). The first evidence that CBCL models’ configuration had little impact on the p-factor is raised by observing the similar global model fit among models. As previously stated, this should not be used as the sole criterion to select a given model (Bornovalova et al., 2020; Clark et al., 2021; Forbes et al., 2021; Moore et al., 2020). Accordingly, model-based reliability indexes were also evaluated.

Present findings demonstrated that these indices are within the range of recent meta-analysis using different types of assessments (Constantinou & Fonagy, 2019). Constantinoy and Fonagy (2019) found a wider range for the H index regarding the p-factor (mean=0.91, SD=0.07), and even wider for specific factors (mean = 0.69, SD=0.17) when compared to present findings. Tha same was observed for FD for the general (mean = 0.93, SD=0.16) and specific factors (mean = 0.84, SD=0.24). Overall, our CBCL findings demonstrate that while using CBCL, factors tend to be well-defined and factor scores are reliable. Regarding specific factors, we found that well-defined and reliable factor scores were the case only for internalizing, externalizing and somatic factors in all models, which consistently showed acceptable H index and FD, and for rule-breaking factors in the Achenbach’s 8 syndrome bifactor model. When present in the model, attention and thought specific factors tend to present H index below 0.80, rendering not well-defined latent constructs in the presence of the p-factor. Nonetheless, the attention factor was the only specific factor associated with symptom impact in daily life in all models and samples.

The second evidence that models’ specification had little impact on CBCL bifactor models came from testing invariance across groups with different characteristics. Previous evidence demonstrates that the CBCL correlated model is invariant across 30 cultures and, therefore, cross-country comparisons can be made (Ivanova et al., 2007; Rescorla et al., 2007). Moreover, previous evidence using different instruments already showed gender, age and race/ethnicity invariance of bifactor models of psychopathology in youth (Gluschkoff et al., 2019; He & Li, 2021; Pezzoli et al., 2017). However, bifactor models were shown to best represent psychopathology in community rather than clinical samples (Fernández de la Cruz et al., 2018). Nonetheless, our findings demonstrate that with respect to the clinical status, bifactor models with different configurations are invariant and mean levels of clinical vs. normative groups can be compared. Thus, our study expands this notion by examining invariance from a broad array of demographic, educational, cognitive and clinical characteristics, as well between raters and time-points. We have not tested, however, mean differences between these groups due to the focus on testing the quality of the models, via invariance testing. Therefore, groups differing in these characteristics probably do not impact mean CBCL scores, not even using the bifactor framework. Furthermore, the 11 tested models cannot be distinguished in their invariance properties, as they perform similarly well.

The third evidence that models’ specification had little impact on CBCL bifactor models comes from the high correlation between p-factors, even when some models, such as the Achenbach 2S model, do not use items from some domains, such as attention, thought and social problems. It is also evident by observing the high correlation among p-factors from symmetrical and S-1 models, as stated above and previously reported (Clark et al., 2021; Moore et al., 2020). The same results can be observed for the specific factors, when compared with homotypic factors from different models. The difference between models comes from different H index and FD of the specific factor as mentioned above, as well as the association with symtpom’s impact. In fact, factor correlation among models (p- and specific) are associated with the differences of factor coefficient on impact, as well as the number and proportion of shared items among specific factors between models. This suggests the relationship between models and external validators might not only be a function of items *per se* but the combination of items in any given model. Therefore, CBCL bifactor models are broadly similar and its specifications must be done depending on the research questions regarding the specific factors to improve its construct definition and determinacy for application of its factor scores.

Limitations should be noted. First, the analysis was carried out using different samples within the RBC, depending on data availability. This may have impacted especially on invariance testing, as some samples, including devCCNP, presented low sample size for the site invariance testing. Notwithstanding, due to the invariance across different characteristics, it is not likely that mean levels of the factors derived from the tested models would be caused by these external sources of variation. Second, we used CBCL only. Despite invariance across informants, correlation among factors may vary using different instruments even if applied to similar models. Future research must elucidate whether p- and specific factors correlate in such proportions when the same model configuration is applied to the same set of harmonized items. Third, we did not manipulate model specifications, such as using EFA-generated models or excluding items with low factor loadings to perform model optimization. Notably, previous studies that have used such strategies reported similar results (Clark et al., 2021).

Hierarchical and dimensional models of psychopathology are thriving as potential empirical classification of mental health problems. With its appropriate criticism and limitations, bifactor models present the possibility to distinguish general from specific dimensions of psychological functioning to improve reliability and validity of nosology. Within this framework, present findings demonstrated that regardless of model configuration and set of items, CBCL bifactor models generally present good fit to the data, are broadly invariant across different characteristics and the p-factor and homotypic specific factors are highly correlated between models. The differences between bifactor models arise when specific factors are analyzed. Internalizing, somatic and externalizing factors tend to be well-defined constructs, while attention and thought specific factors depend upon model specifications. Despite that, attention is consistently associated with symptom impact in daily life. Therefore, using CBCL, model selection should be based on the specific factors of interest. Regardless of the model used, the p-factor will most likely be similar and comparable between different studies. Future research should investigate if these findings hold using different instruments. This could help to generate harmonized data sets between different samples using different instruments and improve replicability while investigating phenotypes, biological and environmental underpinnings of psychopathology.

## Data Availability

Dr. Hoffmann have full access the phenotypic harmonized data set used in the study, as well as analysis code, and takes responsibility for the integrity of the data and the accuracy of the data analysis. Data is available upon request at the moment. However, the end product of the RBC project is to release a complete harmonized data set to the scientific community, combining phenotypic with neuroimaging data. Metadata, R and Mplus code to process raw variables used in this study can be found at https://osf.io/uwy5n/

## Authorship

Mauricio Hoffmann and Giovanni Salum developed the concept of this manuscript. Data collection and preparation was led by Mauricio Hoffmann, Giovanni Salum, Luis Rohde, Nim Tottenham, Xi-Nian Zuo, Michael Milham, Tyler Moore and Theodore Satterthwaite. Mauricio Hoffmann and Luiza Axelrud conducted the analyses and drafted the paper along with Giovanni Salum. All authors agree to the author listing and approve the final version of this paper for submission.

## Acknowledgments

This work was supported by the following grants from the United States National Institutes of Health: R01MH120482-01

