## supplemental material for "Reliability and Validity of Bifactor Models of Dimensional Psychopathology in Youth from three Continents"

### Supplementary Material

Figure S1: Flowchart of the bifactor models using Child Behavioural Checklist search

Figure S2: Correlation of between-model regression coefficient differences of symptom impact regressed on p- and specific factors and between-model p- and specific factor correlations

Figure S3: Correlation of between-model regression coefficient differences of symptom impact regressed on p- and specific factors and between-model p- and specific factor shared items

The supplemental tables below also can be found at https://osf.io/uwy5n/files/ in .xlsx files:

Table S1: Data description

Table S2: CBCL two-syndrome bifactor model (specific factors according to the ASEBA system - Achenbach 2S)

Table S3: CBCL eight-syndrome bifactor model (specific factors according to the ASEBA system - Achenbach 8S)

Table S4: CBCL Bifactor model according to Moore et al., using three specific factors

Table S5: CBCL Bifactor model according to Moore et al., using four specific factors

Table S6: CBCL Bifactor model according to McElroy et al., using three specific factors

Table S7: CBCL Bifactor model according to Deutz et al., using two specific factors (GP model)

Table S8: CBCL Bifactor model according to Deutz et al. and Haltigan et al., using three specific factors (DP model)

Table S9: CBCL Bifactor model according to Haltigan et al., using four specific factors

Table S10: CBCL Bifactor model according to Clark et al., using two specific factors

Table S11: CBCL Bifactor model according to Clark et al., using three specific factors

Table S12: CBCL Bifactor model according to Clark et al., using four specific factors

Table S13: Impact model - global fit and factor loading

Table S14: Global fit indices of structural equation models of CBCL factors predicting symptom impact

Table S15: Results of structural equation models of latent symptom impact (SDQ) predicted by latent factors of each CBCL model

Table S16: Measurement invariance testing of CBCL models across age groups

Table S17: Measurement invariance testing of CBCL models across gender groups

Table S18: Measurement invariance testing of CBCL models across study site

Table S19: Measurement invariance testing of CBCL models across race/ethnicity groups

Table S20: Measurement invariance testing of CBCL models across educational level groups

Table S21: Measurement invariance testing of CBCL models across IQ groups

Table S22: Measurement invariance testing of CBCL models across mental health condition status (presence vs. no condition)

Table S23: Measurement invariance testing of CBCL models across study waves

Table S24: Measurement invariance testing of CBCL models across informants (parent-CBCL, teacher-TRF and self-YSR)

Table S25: Correlation between p-factors derived from 11 CBCL bifactor models

Table S26: Correlation between specific factors derived from 11 CBCL bifactor models

**Model-based reliability indices**

We used 10 model-based reliability indices to evaluate the bifactor models (Constantinou & Fonagy, 2019; Rodriguez et al., 2016):

1. Reliability index omega (ω) is a model-based reliability estimate, analogous to the alpha coefficient, but appropriate for tests that have varying factor loadings;
2. Hierarchical omega (ωH), the proportion of total variance attributed to the general or specific factors (ω and ωH coefficients vary between 0 and 1, where higher scores indicate greater reliability);
3. Factor determinacy (FD): the correlation between the factor scores and the estimated factor, indicates that factor scores should be used if FD are at least 0.9;
4. H: a measure of construct replicability that quantifies how well each latent factor is represented by the items loading on it (H > 0.7 represents a well-defined latent variable (Hancock & Mueller, 2001);
5. Explained common variance (ECV): the proportion of the total variance in included items which is explained by the general factor rather than the specific factors;
6. ECV of a specific factor due to itself (ECV-SS) is the proportion of common variance of the items in each factor which is due to that factor;
7. ECV of a specific factor with respect to the general factor (ECV SG) is the proportion of common variance of the items in each specific factor which is due to the specific factor;
8. ECV of the general factor with respect to a specific factor (ECV GS) is the proportion of common variance of the items in each specific factor which is due to the general factor;
9. Percent uncontaminated correlations (PUC): the percent of all correlations among symptoms attributable purely to the general factor; and

10. The Item Explained Common Variance (IECV), which "provides the extent to which an item's responses are accounted for by variation on the latent general dimension alone, and this acts as an assessment of the unidimensionality at the individual item level" (IECV $\geq$ 0.85 yield unidimensional item sets that reflect the content of the general factor) (Stucky & Edelen, 2015).

**Criterion validity supplementary generalized additive models**

We performed supplementary generalized additive models (GAM) to test if between-model regression coefficient differences of impact latent variable on p- and paired specific factors was associated with the between-model 1) factors scores correlation and 2) the number and proportion of shared items. GAM was implemented in R using (Wood & Scheipl, 2020)

The observed between-models p-factor (HBN $F(1,54)=13.07$, $p=4.6\text{x}10^{-3}$; BHRCS $F(1,54)=2.63$, $p=0.108$; $R^2=0.124$; Figure S2) and specific factor correlation (HBN $F(1,106)=13.52$, $p=2.0\text{x}10^{-16}$; BHRCS $F(1,106)=5.69$, $p=0.001$; $R^2=0.277$; Figure S2) are associated with regression coefficient differences from each model on impact. Interestingly, the number (HBN $F(1,54)=2.12$, $p=0.087$; BHRCS $F(1,54)=2.20$, $p=0.141$; $R^2=0.04$; Figure S3) and proportion (HBN $F(1,54)=1.21$, $p=0.206$; BHRCS $F(1,54)=0.37$, $p=0.543$; $R^2=0.008$; Figure S3) of p-factor shared items between models are not associated with regression coefficient differences from each model on impact. The same does not hold for the specific factors, whether the number (HBN $F(1,105)=11.91$, $p=1.72\text{x}10^{-6}$; BHRCS $F(1,105)=4.96$, $p=9.67\text{x}10^{-3}$; $R^2=0.202$; Figure S4) and proportion (HBN $F(1,54)=3.47$, $p=0.014$; BHRCS $F(1,54)=3.04$, $p=0.029$; $R^2=0.08$; Figure S3) of shared items between models are associated with regression coefficient differences from each model on impact.

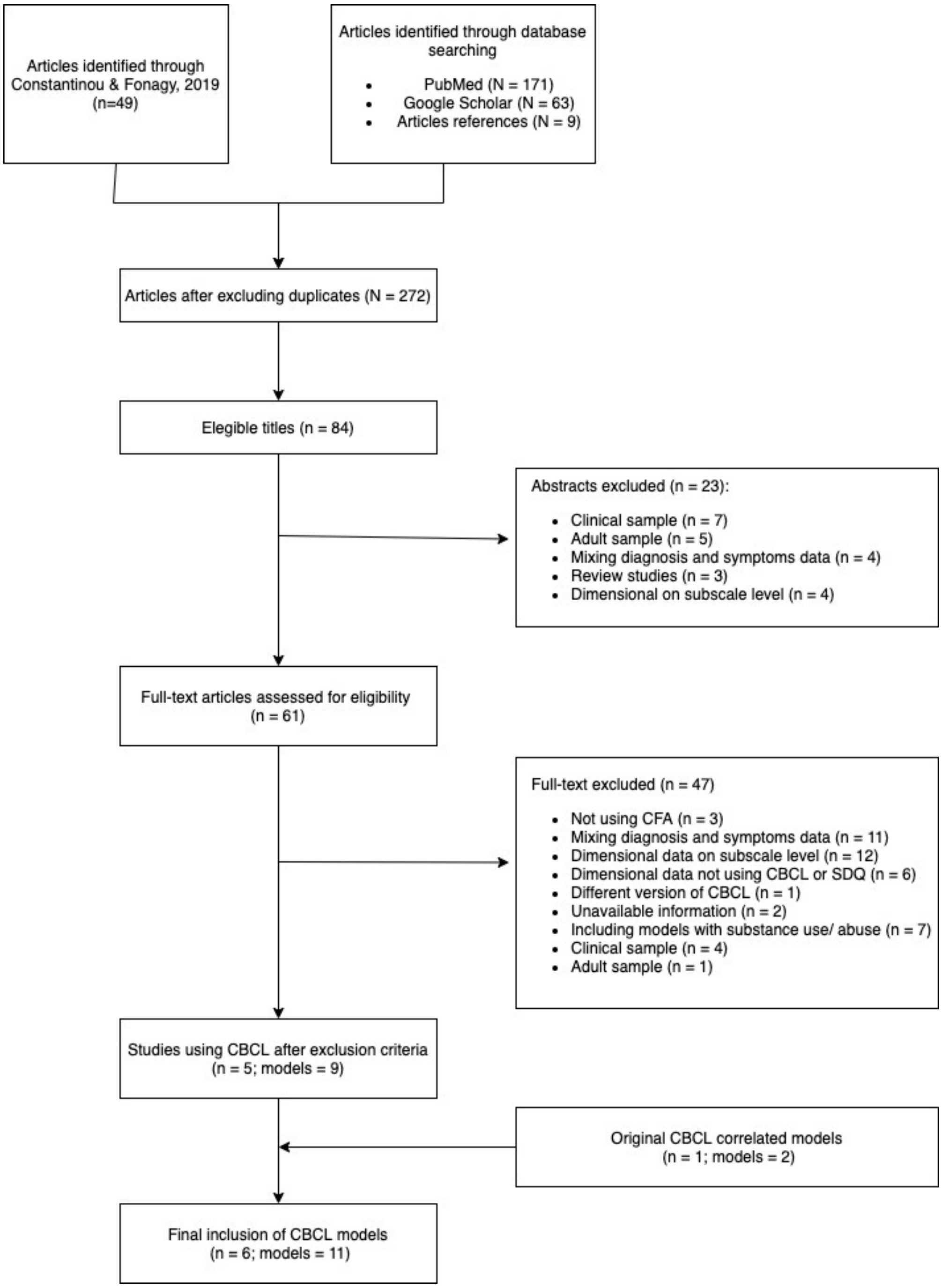


Figure S1: Flowchart of the bifactor models using Child Behavioural Checklist search

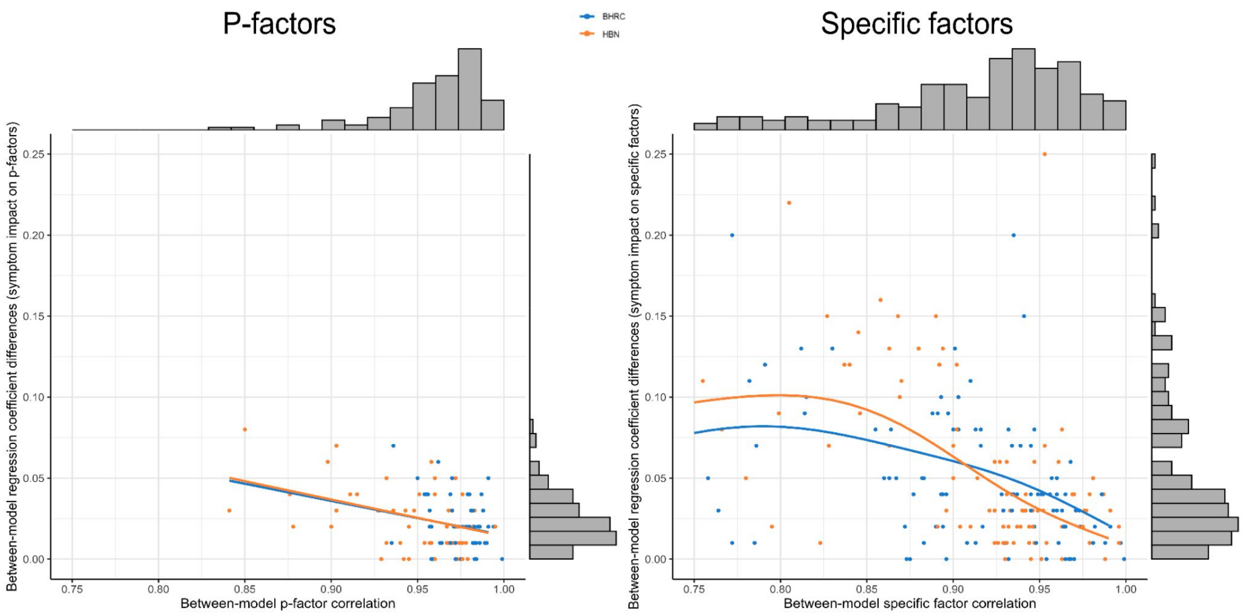


Figure S2: Correlation of between-model regression coefficient differences of symptom impact regressed on p- and specific factors and between-model p- and specific factor correlations. Models were fitted using generalized additive models. Marginal distributions are plotted referring to the Y and X axis. BHRC, Brazilian High-Risk Cohort; HBN, Healthy Brain Network.

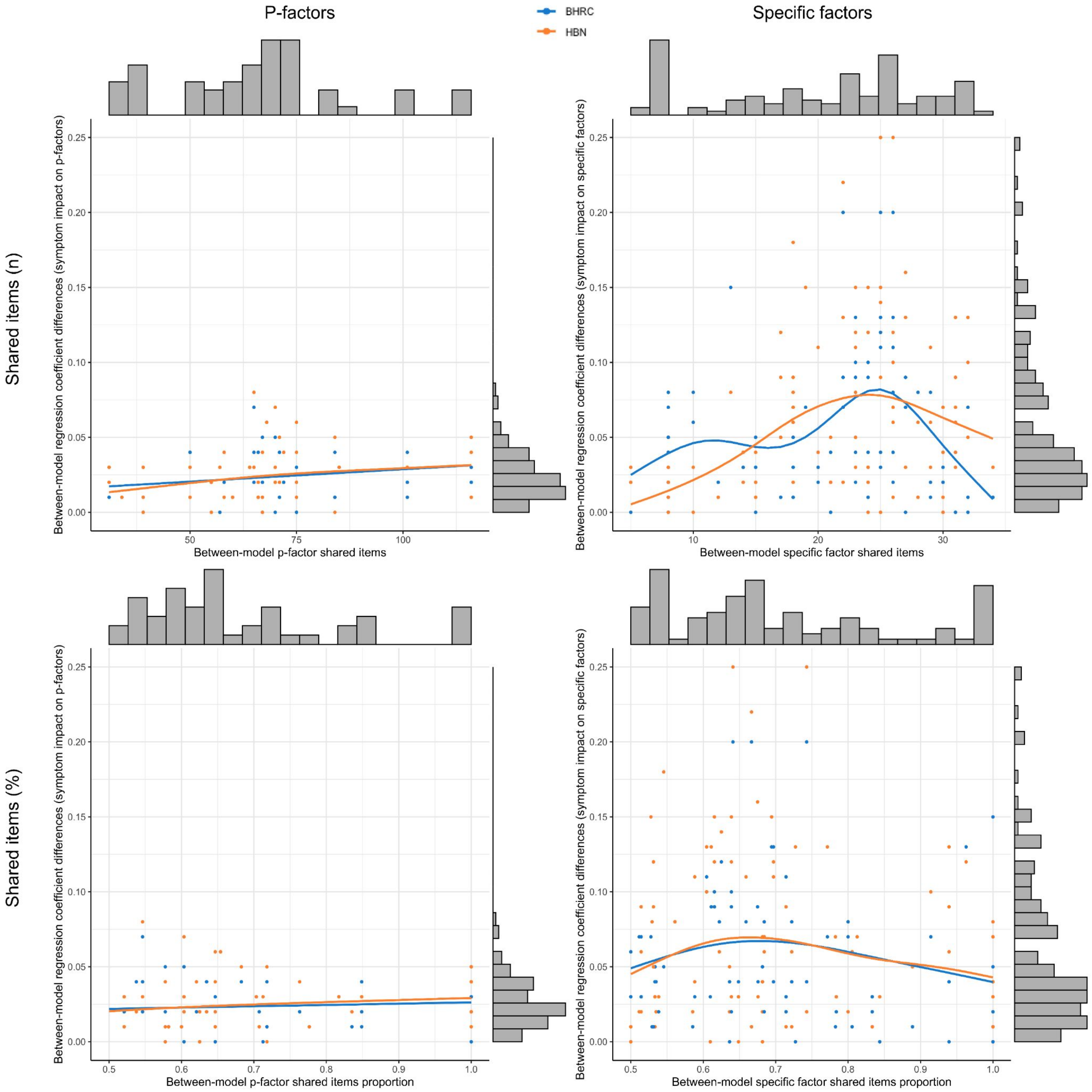


Figure S3: Correlation of between-model regression coefficient differences of symptom impact regressed on p- and specific factors and between-model p- (left plots) and specific factor (right plots) number (upper plots) and proportion (bottom plots) of shared items. Models were fitted using generalized additive models. Marginal distributions are plotted referring to the Y and X axis. BHRC, Brazilian High-Risk Cohort; HBN, Healthy Brain Network.

All tables can be found at https://osf.io/uwy5n/files/ in .xlsx files.

Table S1 - Descriptive table

| | | BHRCS (n=2511) | devCCNP (n=181) | PACCT (n=312) | HBN (n=3633) | NKI (n=374) | Total sample (N=7011) |
|---|---|---|---|---|---|---|---|
| Age (years) | | | | | | | |
| | Mean (SD) | 10 (± 1.9) | 12 (± 3.0) | NA | 10 (± 3.6) | 11 (± 3.2) | 10 (± 3.0) |
| | Missing | 0 (0%) | 0 (0%) | | 0 (0%) | 0 (0%) | 312 (4.5%) |
| Gender | | | | | | | |
| | Female | 1136 (45 %) | 95 (52 %) | NA | 1295 (36 %) | 168 (45 %) | 2694 (38 %) |
| | Male | 1375 (55 %) | 86 (48 %) | | 2338 (64 %) | 205 (55 %) | 4004 (57 %) |
| | Missing | 0 (0%) | 0 (0%) | | 0 (0%) | 1 (0.3%) | 313 (4.5%) |
| Child race/ethnicity | | | | | | | |
| | Asian | 5 (0 %) | NA | NA | 114 (3 %) | 25 (7 %) | 144 (2 %) |
| | Black | 264 (11 %) | | | 481 (13 %) | 78 (21 %) | 823 (12 %) |
| | Mixed | 706 (28 %) | | | 835 (23 %) | 0 (0 %) | 1541 (22 %) |

| | | | | | | |
|---|---|---|---|---|---|---|
| Native | 11 (0 %) | | | 8 (0 %) | 9 (2 %) | 28 (0 %) |
| White | 1519 (60 %) | | | 1595 (44 %) | 249 (67 %) | 3363 (48 %) |
| Other | 0 (0 %) | | | 74 (2 %) | 13 (3 %) | 87 (1 %) |
| Unknown | 0 (0 %) | | | 19 (1 %) | 0 (0 %) | 19 (0 %) |
| Missing | 6 (0.2%) | | | 507 (14.0%) | 0 (0%) | 1006 (14.3%) |
| IQ | | | | | | |
| Mean (SD) | 100 (± 15) | NA | NA | 100 (± 15) | 100 (± 15) | 100 (± 15) |
| Missing | 270 (10.8%) | | | 585 (16.1%) | 5 (1.3%) | 1353 (19.3%) |
| Educational attainment (years) | | | | | | |
| Mean (SD) | 7.5 (± 1.9) | NA | NA | 6.6 (± 3.5) | 5.8 (± 3.2) | 7.0 (± 2.8) |
| Missing | 41 (2.0%) | | | 2237 (61.6%) | 9 (2.4%) | 2780 (42.7%) |
| Educational attainment (binary) | | | | | | |
| Primary school | 1671 (83.1%) | NA | NA | 1030 (28.3 %) | 274 (73 %) | 2975 (45.7 %) |
| Secondary school | 298 (14.8 %) | | | 366 (10.0 %) | 91 (24 %) | 755 (11.6 %) |
| Missing | 41 (2.0%) | | | 2237 (61.6%) | 9 (2.4%) | 2780 (42.7%) |

| | | | | | | |
|---|---|---|---|---|---|---|
| Any psychiatric diagnosis | | | | | | |
| Not present | 1859 (74 %) | NA | NA | 308 (8 %) | 190 (51 %) | 2357 (34 %) |
| Present | 652 (26 %) | | | 2859 (79 %) | 177 (47 %) | 3688 (53 %) |
| Missing | 0 (0%) | | | 466 (12.8%) | 7 (1.9%) | 966 (13.8%) |
| CBCL Total score | | | | | | |
| Median [Min, Max] | 23.0 [0, 164] | 18.0 [0, 70.0] | 21.0 [0, 119] | 32.0 [0, 212] | 17.0 [0, 93.0] | 26.0 [0, 212] |
| CBCL Anxiety/depressive score | | | | | | |
| Median [Min, Max] | 3.00 [0, 24.0] | 1.00 [0, 12.0] | 3.00 [0, 23.0] | 3.00 [0, 101] | 2.00 [0, 18.0] | 3.00 [0, 101] |
| CBCL Withdraw/depressive score | | | | | | |
| Median [Min, Max] | 1.00 [0, 16.0] | 1.00 [0, 13.0] | 1.00 [0, 11.0] | 1.00 [0, 16.0] | 1.00 [0, 14.0] | 1.00 [0, 16.0] |
| CBCL Somatic score | | | | | | |
| Median [Min, Max] | 2.00 [0, 22.0] | 1.00 [0, 9.00] | 1.00 [0, 12.0] | 1.00 [0, 24.0] | 1.00 [0, 13.0] | 1.00 [0, 24.0] |
| CBCL Rule breaking score | | | | | | |
| Median [Min, Max] | 1.00 [0, 24.0] | 2.00 [0, 10.0] | 1.00 [0, 13.0] | 1.00 [0, 28.0] | 1.00 [0, 13.0] | 1.00 [0, 28.0] |
| CBCL Aggressive behavior score | | | | | | |

| | | | | | | |
|---|---|---|---|---|---|---|
| Median [Min, Max] | 4.00 [0, 36.0] | 3.00 [0, 18.0] | 4.00 [0, 32.0] | 5.00 [0, 107] | 2.00 [0, 24.0] | 4.00 [0, 107] |
| CBCL Social problems score | | | | | | |
| Median [Min, Max] | 2.00 [0, 20.0] | 2.00 [0, 11.0] | 2.00 [0, 16.0] | 3.00 [0, 20.0] | 1.00 [0, 15.0] | 2.00 [0, 20.0] |
| CBCL Thought problems score | | | | | | |
| Median [Min, Max] | 1.00 [0, 28.0] | 1.00 [0, 9.00] | 1.00 [0, 15.0] | 2.00 [0, 25.0] | 1.00 [0, 13.0] | 1.00 [0, 28.0] |
| CBCL Attentional problems score | | | | | | |
| Median [Min, Max] | 4.00 [0, 20.0] | 2.00 [0, 13.0] | 3.00 [0, 15.0] | 7.00 [0, 20.0] | 3.00 [0, 18.0] | 5.00 [0, 20.0] |
| CBCL Other problems score | | | | | | |
| Median [Min, Max] | 3.00 [0, 20.0] | 2.00 [0, 10.0] | 3.00 [0, 13.0] | 3.00 [0, 28.0] | 2.00 [0, 13.0] | 3.00 [0, 28.0] |

Note: CBCL, Child Behavior Checklist; NA, data not available; IQ, standardized Intelligence quotient adjusted by age and gender; BHRCS, Brazilian High-Risk Cohort Study; HBN, Healthy Brain Network; NKI, Nathan Kline Institute-Rockland Sample; devCCNP, developmental component of the Chinese Color Nest Project; PACCT, Parents and Children Coming Together. Educational attainment was collected in the second data collection for the BHRCS.

Table S2 - CBCL two-syndrome bifactor model (specific factors according the ASEBA system - Achenbach 2S model)

| Content | Item | Factors and factor loadings | | | IECV |
|---|---|---|---|---|---|
| | | P-factor | Internalizing | Externalizing | |
| Cries | CBCL_14 | 0.588 | 0.084 | | 0.980 |
| Fears | CBCL_29 | 0.380 | 0.242 | | 0.711 |
| Fears school | CBCL_30 | 0.507 | 0.280 | | 0.766 |
| Fears do bad | CBCL_31 | 0.522 | 0.160 | | 0.914 |
| Perfect | CBCL_32 | 0.342 | 0.264 | | 0.627 |
| Unloved | CBCL_33 | 0.743 | 0.028 | | 0.999 |
| Worthless | CBCL_35 | 0.698 | 0.207 | | 0.919 |
| Nervous | CBCL_45 | 0.705 | 0.268 | | 0.874 |
| Fearful | CBCL_50 | 0.556 | 0.423 | | 0.633 |
| Feels too guilty | CBCL_52 | 0.546 | 0.346 | | 0.713 |
| Self-conscious | CBCL_71 | 0.533 | 0.361 | | 0.686 |

| | | | | |
|---|---|---|---|---|
| Talks about suicide | CBCL_91 | 0.671 | -0.037 | 0.997 |
| Worries | CBCL_112 | 0.500 | 0.418 | 0.589 |
| Little they enjoy | CBCL_5 | 0.641 | 0.205 | 0.907 |
| Prefers alone | CBCL_42 | 0.448 | 0.407 | 0.548 |
| Won’t talk | CBCL_65 | 0.562 | 0.219 | 0.868 |
| Secretive | CBCL_69 | 0.554 | 0.261 | 0.818 |
| Shy | CBCL_75 | 0.296 | 0.426 | 0.326 |
| Lacks energy | CBCL_102 | 0.461 | 0.427 | 0.538 |
| Sad | CBCL_103 | 0.695 | 0.319 | 0.826 |
| Withdrawn | CBCL_111 | 0.481 | 0.481 | 0.500 |
| Nightmares | CBCL_47 | 0.489 | 0.190 | 0.869 |
| Constipate | CBCL_49 | 0.324 | 0.271 | 0.588 |
| Dizzy | CBCL_51 | 0.397 | 0.551 | 0.342 |
| Tired | CBCL_54 | 0.567 | 0.360 | 0.713 |
| Aches | CBCL_56A | 0.410 | 0.460 | 0.443 |

| | | | | | |
|---|---|---|---|---|---|
| Headaches | CBCL_56B | 0.326 | 0.522 | | 0.281 |
| Nausea | CBCL_56C | 0.351 | 0.754 | | 0.178 |
| Eye problems | CBCL_56D | 0.263 | 0.242 | | 0.542 |
| Skin problems | CBCL_56E | 0.279 | 0.223 | | 0.610 |
| Stomach | CBCL_56F | 0.358 | 0.623 | | 0.248 |
| Vomit | CBCL_56G | 0.328 | 0.615 | | 0.221 |
| Other physical problem | CBCL_56H | 0.280 | 0.252 | | 0.552 |
| Drinks | CBCL_2 | 0.215 | | 0.213 | 0.505 |
| No guilt | CBCL_26 | 0.521 | | 0.497 | 0.524 |
| Breaks rules at home | CBCL_28 | 0.548 | | 0.688 | 0.388 |
| Bad friends | CBCL_39 | 0.427 | | 0.510 | 0.412 |
| Lies or cheats | CBCL_43 | 0.486 | | 0.548 | 0.440 |
| Prefers older | CBCL_63 | 0.382 | | 0.170 | 0.835 |
| Runs away | CBCL_67 | 0.447 | | 0.368 | 0.596 |
| Sets fires | CBCL_72 | 0.380 | | 0.286 | 0.638 |

| | | | | |
|---|---|---|---|---|
| Sex problems | CBCL_73 | 0.404 | 0.224 | 0.765 |
| Steals from home | CBCL_81 | 0.359 | 0.649 | 0.234 |
| Steals outside home | CBCL_82 | 0.304 | 0.669 | 0.171 |
| Swears | CBCL_90 | 0.540 | 0.357 | 0.696 |
| Thinks about sex | CBCL_96 | 0.488 | 0.233 | 0.814 |
| Smoke | CBCL_99 | 0.301 | 0.217 | 0.658 |
| Truants | CBCL_101 | 0.514 | 0.161 | 0.911 |
| Use drugs | CBCL_105 | 0.348 | 0.300 | 0.574 |
| Vandalism | CBCL_106 | 0.480 | 0.574 | 0.412 |
| Argues | CBCL_3 | 0.634 | 0.397 | 0.718 |
| Mean | CBCL_16 | 0.500 | 0.604 | 0.407 |
| Demands a lot of attention | CBCL_19 | 0.655 | 0.293 | 0.833 |
| Destroys own things | CBCL_20 | 0.478 | 0.617 | 0.375 |
| Destroys other | CBCL_21 | 0.488 | 0.715 | 0.318 |
| Disobedient at home | CBCL_22 | 0.580 | 0.664 | 0.433 |

| | | | | | |
|---|---|---|---|---|---|
| Disobedient at school | CBCL_23 | 0.416 | | 0.676 | 0.275 |
| Fights | CBCL_37 | 0.528 | | 0.537 | 0.492 |
| Attacks | CBCL_57 | 0.497 | | 0.626 | 0.387 |
| Screams | CBCL_68 | 0.630 | | 0.383 | 0.730 |
| Stubborn | CBCL_86 | 0.778 | | 0.259 | 0.900 |
| Mood changes | CBCL_87 | 0.834 | | 0.089 | 0.989 |
| Sulks | CBCL_88 | 0.805 | | -0.034 | 0.998 |
| Suspicious | CBCL_89 | 0.702 | | 0.020 | 0.999 |
| Teases | CBCL_94 | 0.549 | | 0.416 | 0.635 |
| Temper | CBCL_95 | 0.705 | | 0.368 | 0.786 |
| Threatens | CBCL_97 | 0.560 | | 0.586 | 0.477 |
| Loud | CBCL_104 | 0.532 | | 0.390 | 0.650 |
| Index | | | | | |
| | PUC | 0.507 | | | |
| | ECV SS | 0.606 | 0.355 | 0.422 | |

| | | | |
|---|---|---|---|
| ECV SG | 0.606 | 0.151 | 0.243 |
| ECV GS | 0.606 | 0.645 | 0.578 |
| Omega | 0.941 | 0.888 | 0.915 |
| OmegaH | 0.740 | 0.262 | 0.330 |
| H | 0.968 | 0.865 | 0.919 |
| FD | 0.970 | 0.921 | 0.951 |

Note: IECV, item explained common variance (≥ 0.85 yield unidimensional item sets that reflect the content of the general factor); ECV, explained common variance; SS, proportion of common variance of the items in each factor which is due to that factor; SG, ECV proportion of common variance of the items in each specific factor which is due to the specific factor; GS, proportion of common variance of the items in each specific factor which is due to the general factor; PUC, percent of uncontaminated correlations; OmegaH, omega-hierarchical; H, index of construct replicability (>0.8 suggests a well-defined latent variable); FD, factor determinacy (>0.9 indicate that the factor score can be used)

Table S3 - CBCL eight-syndrome bifactor model (specific factors according the ASEBA system - Achenbach 8S model)

| Content | Item | Factors and factor loadings | | | | | | | | | IECV |
|---|---|---|---|---|---|---|---|---|---|---|---|
| | | P-factor | Anxious-depressed | Withdraw-depressed | Somatic | Rule breaking | Aggressive | Social problems | Thought problems | Attention problems | |
| Cries | CBCL_14 | 0.551 | 0.157 | | | | | | | | 0.925 |
| Fears | CBCL_29 | 0.382 | 0.346 | | | | | | | | 0.549 |
| Fears school | CBCL_30 | 0.513 | 0.321 | | | | | | | | 0.719 |
| Fears do bad | CBCL_31 | 0.486 | 0.408 | | | | | | | | 0.587 |
| Perfect | CBCL_32 | 0.318 | 0.504 | | | | | | | | 0.285 |
| Unloved | CBCL_33 | 0.654 | 0.179 | | | | | | | | 0.930 |
| Worthless | CBCL_35 | 0.668 | 0.323 | | | | | | | | 0.811 |
| Nervous | CBCL_45 | 0.673 | 0.371 | | | | | | | | 0.767 |
| Fearful | CBCL_50 | 0.551 | 0.585 | | | | | | | | 0.470 |
| Feels too guilty | CBCL_52 | 0.540 | 0.491 | | | | | | | | 0.547 |

| | | | | | | |
|---|---|---|---|---|---|---|
| Self-conscious | CBCL_71 | 0.554 | 0.324 | | | 0.745 |
| Talks about suicide | CBCL_91 | 0.577 | 0.108 | | | 0.966 |
| Worries | CBCL_112 | 0.520 | 0.514 | | | 0.506 |
| Little they enjoy | CBCL_5 | 0.609 | | 0.340 | | 0.762 |
| Prefers alone | CBCL_42 | 0.480 | | 0.603 | | 0.388 |
| Won’t talk | CBCL_65 | 0.524 | | 0.453 | | 0.572 |
| Secretive | CBCL_69 | 0.517 | | 0.461 | | 0.557 |
| Shy | CBCL_75 | 0.339 | | 0.464 | | 0.348 |
| Lacks energy | CBCL_102 | 0.518 | | 0.452 | | 0.568 |
| Sad | CBCL_103 | 0.664 | | 0.351 | | 0.782 |
| Withdrawn | CBCL_111 | 0.516 | | 0.712 | | 0.344 |
| Nightmares | CBCL_47 | 0.507 | | | 0.183 | 0.885 |
| Constipate | CBCL_49 | 0.352 | | | 0.255 | 0.656 |
| Dizzy | CBCL_51 | 0.446 | | | 0.548 | 0.398 |
| Tired | CBCL_54 | 0.589 | | | 0.275 | 0.821 |

| | | | | | |
|---|---|---|---|---|---|
| Aches | CBCL_56A | 0.447 | 0.492 | | 0.452 |
| Headaches | CBCL_56B | 0.339 | 0.656 | | 0.211 |
| Nausea | CBCL_56C | 0.423 | 0.805 | | 0.216 |
| Eye problems | CBCL_56D | 0.296 | 0.261 | | 0.563 |
| Skin problems | CBCL_56E | 0.314 | 0.204 | | 0.703 |
| Stomach | CBCL_56F | 0.413 | 0.697 | | 0.260 |
| Vomit | CBCL_56G | 0.380 | 0.682 | | 0.237 |
| Other physical problem | CBCL_56H | 0.331 | 0.177 | | 0.778 |
| Drinks | CBCL_2 | 0.179 | | 0.600 | 0.082 |
| No guilt | CBCL_26 | 0.658 | | 0.237 | 0.885 |
| Breaks rules at home | CBCL_28 | 0.790 | | 0.242 | 0.914 |
| Bad friends | CBCL_39 | 0.575 | | 0.367 | 0.711 |
| Lies or cheats | CBCL_43 | 0.640 | | 0.487 | 0.633 |
| Prefers older | CBCL_63 | 0.418 | | 0.056 | 0.982 |

| | | | | | |
|---|---|---|---|---|---|
| Runs away | CBCL_67 | 0.500 | 0.385 | | 0.628 |
| Sets fires | CBCL_72 | 0.424 | 0.286 | | 0.687 |
| Sex problems | CBCL_73 | 0.418 | 0.348 | | 0.591 |
| Steals from home | CBCL_81 | 0.535 | 0.666 | | 0.392 |
| Steals outside home | CBCL_82 | 0.484 | 0.690 | | 0.330 |
| Swears | CBCL_90 | 0.586 | 0.233 | | 0.863 |
| Thinks about sex | CBCL_96 | 0.489 | 0.373 | | 0.632 |
| Smoke | CBCL_99 | 0.220 | 0.692 | | 0.092 |
| Truants | CBCL_101 | 0.461 | 0.382 | | 0.593 |
| Use drugs | CBCL_105 | 0.290 | 0.772 | | 0.124 |
| Vandalism | CBCL_106 | 0.627 | 0.374 | | 0.738 |
| Argues | CBCL_3 | 0.708 | | 0.236 | 0.900 |
| Mean | CBCL_16 | 0.637 | | 0.454 | 0.663 |
| Demands a lot of attention | CBCL_19 | 0.712 | | 0.141 | 0.962 |

| | | | | |
|---|---|---|---|---|
| Destroys own things | CBCL_20 | 0.609 | 0.548 | 0.553 |
| Destroys other | CBCL_21 | 0.653 | 0.617 | 0.528 |
| Disobedient at home | CBCL_22 | 0.746 | 0.438 | 0.744 |
| Disobedient at school | CBCL_23 | 0.634 | 0.395 | 0.720 |
| Fights | CBCL_37 | 0.649 | 0.367 | 0.758 |
| Attacks | CBCL_57 | 0.628 | 0.537 | 0.578 |
| Screams | CBCL_68 | 0.680 | 0.291 | 0.845 |
| Stubborn | CBCL_86 | 0.769 | 0.145 | 0.966 |
| Mood changes | CBCL_87 | 0.784 | -0.025 | 0.999 |
| Sulks | CBCL_88 | 0.699 | -0.100 | 0.980 |
| Suspicious | CBCL_89 | 0.637 | -0.120 | 0.966 |
| Teases | CBCL_94 | 0.611 | 0.286 | 0.820 |
| Temper | CBCL_95 | 0.732 | 0.290 | 0.864 |
| Threatens | CBCL_97 | 0.664 | 0.495 | 0.643 |

| | | | | | | |
|---|---|---|---|---|---|---|
| Loud | CBCL_104 | 0.628 | 0.257 | | | 0.857 |
| Too dependent | CBCL_11 | 0.545 | | -0.108 | | 0.962 |
| Lonely | CBCL_12 | 0.596 | | 0.052 | | 0.992 |
| Doesn’t get along with peers | CBCL_25 | 0.689 | | 0.440 | | 0.710 |
| Jealous | CBCL_27 | 0.680 | | -0.020 | | 0.999 |
| Feels persecuted | CBCL_34 | 0.667 | | 0.191 | | 0.924 |
| Gets hurt a lot | CBCL_36 | 0.592 | | -0.127 | | 0.956 |
| Gets teased | CBCL_38 | 0.605 | | 0.394 | | 0.702 |
| Not liked by peers | CBCL_48 | 0.666 | | 0.685 | | 0.486 |
| Clumsy | CBCL_62 | 0.628 | | -0.138 | | 0.954 |
| Prefers younger kids | CBCL_64 | 0.398 | | 0.058 | | 0.979 |
| Speech problem | CBCL_79 | 0.349 | | -0.045 | | 0.984 |
| Mind off | CBCL_9 | 0.675 | | | 0.239 | 0.889 |
| Harms self | CBCL_18 | 0.528 | | | -0.040 | 0.994 |

| | | | | | |
|---|---|---|---|---|---|
| Hears things | CBCL_40 | 0.500 | 0.500 | | 0.500 |
| Twitches | CBCL_46 | 0.549 | 0.268 | | 0.808 |
| Picks skin | CBCL_58 | 0.499 | 0.170 | | 0.896 |
| Plays with sex parts in public | CBCL_59 | 0.421 | 0.554 | | 0.366 |
| Sex parts | CBCL_60 | 0.469 | 0.477 | | 0.492 |
| Repeats acts | CBCL_66 | 0.596 | 0.309 | | 0.788 |
| Sees things | CBCL_70 | 0.497 | 0.490 | | 0.507 |
| Sleeps less | CBCL_76 | 0.481 | 0.352 | | 0.651 |
| Stores up | CBCL_83 | 0.507 | 0.152 | | 0.918 |
| Strange behavior | CBCL_84 | 0.636 | 0.367 | | 0.750 |
| Strange ideas | CBCL_85 | 0.600 | 0.404 | | 0.688 |
| Sleep walks | CBCL_92 | 0.341 | 0.142 | | 0.852 |
| Sleep problems | CBCL_100 | 0.532 | 0.341 | | 0.709 |
| Acts too young | CBCL_1 | 0.527 | | 0.254 | 0.811 |

| | | | | | | | | | | | |
|---|---|---|---|---|---|---|---|---|---|---|---|
| Fails to finish things | CBCL_4 | 0.673 | | | | | | | | 0.443 | 0.698 |
| Can’t concentrate | CBCL_8 | 0.654 | | | | | | | | 0.663 | 0.493 |
| Can’t sit still | CBCL_10 | 0.605 | | | | | | | | 0.406 | 0.689 |
| Confused | CBCL_13 | 0.621 | | | | | | | | 0.320 | 0.790 |
| Daydreams | CBCL_17 | 0.574 | | | | | | | | 0.485 | 0.583 |
| Impulsive | CBCL_41 | 0.767 | | | | | | | | 0.258 | 0.898 |
| Poor school work | CBCL_61 | 0.565 | | | | | | | | 0.360 | 0.711 |
| Inattentive | CBCL_78 | 0.657 | | | | | | | | 0.609 | 0.538 |
| Stares | CBCL_80 | 0.552 | | | | | | | | 0.406 | 0.649 |
| Index | | | | | | | | | | | |
| PUC | | 0.876 | | | | | | | | | |
| ECV SS | | 0.653 | 0.329 | 0.466 | 0.587 | 0.452 | 0.220 | 0.190 | 0.308 | 0.333 | |
| ECV SG | | 0.653 | 0.038 | 0.039 | 0.057 | 0.073 | 0.047 | 0.018 | 0.037 | 0.038 | |
| ECV GS | | 0.653 | 0.671 | 0.534 | 0.413 | 0.548 | 0.780 | 0.810 | 0.692 | 0.667 | |
| Omega | | 0.962 | 0.811 | 0.785 | 0.756 | 0.848 | 0.905 | 0.757 | 0.801 | 0.840 | |

| | | | | | | | | | |
|---|---|---|---|---|---|---|---|---|---|
| OmegaH | 0.914 | 0.247 | 0.347 | 0.398 | 0.358 | 0.142 | 0.030 | 0.194 | 0.234 |
| H | 0.982 | 0.710 | 0.746 | 0.846 | 0.861 | 0.755 | 0.584 | 0.700 | 0.731 |
| FD | 0.983 | 0.874 | 0.904 | 0.942 | 0.940 | 0.912 | 0.916 | 0.862 | 0.920 |

Note: IECV, item explained common variance (≥ 0.85 yield unidimensional item sets that reflect the content of the general factor); ECV, explained common variance; SS, proportion of common variance of the items in each factor which is due to that factor; SG, ECV proportion of common variance of the items in each specific factor which is due to the specific factor; GS, proportion of common variance of the items in each specific factor which is due to the general factor; PUC, percent of uncontaminated correlations; OmegaH, omega-hierarchical; H, index of construct replicability (> 0.8 suggests a well-defined latent variable); FD, factor determinacy (> 0.9 indicate that the factor score can be used)

Table S4 - CBCL Bifactor model according to Moore et al., using three specific factors

| Content | Item | Factors and factor loadings | | | | IECV |
|---|---|---|---|---|---|---|
| | | P-factor | Internalizing | Externalizing | Attention | |
| Little they enjoy | CBCL_5 | 0.592 | 0.291 | | | 0.805 |
| Fears school | CBCL_30 | 0.480 | 0.315 | | | 0.699 |
| Fears do bad | CBCL_31 | 0.489 | 0.212 | | | 0.842 |
| Perfect | CBCL_32 | 0.306 | 0.307 | | | 0.498 |
| Worthless | CBCL_35 | 0.652 | 0.297 | | | 0.828 |
| Prefers alone | CBCL_42 | 0.453 | 0.436 | | | 0.519 |
| Fearful | CBCL_50 | 0.506 | 0.420 | | | 0.592 |
| Feels too guilty | CBCL_52 | 0.512 | 0.386 | | | 0.638 |
| Won’t talk | CBCL_65 | 0.511 | 0.317 | | | 0.722 |
| Secretive | CBCL_69 | 0.496 | 0.357 | | | 0.659 |
| Self-conscious | CBCL_71 | 0.528 | 0.396 | | | 0.640 |

| | | | | | | |
|---|---|---|---|---|---|---|
| Shy | CBCL_75 | 0.282 | 0.451 | | | 0.281 |
| Lacks energy | CBCL_102 | 0.473 | 0.418 | | | 0.561 |
| Sad | CBCL_103 | 0.613 | 0.441 | | | 0.659 |
| Withdrawn | CBCL_111 | 0.484 | 0.517 | | | 0.467 |
| Worries | CBCL_112 | 0.489 | 0.432 | | | 0.562 |
| Dizzy | CBCL_51 | 0.362 | 0.555 | | | 0.298 |
| Aches | CBCL_56A | 0.402 | 0.446 | | | 0.448 |
| Headaches | CBCL_56B | 0.282 | 0.528 | | | 0.222 |
| Nausea | CBCL_56C | 0.348 | 0.741 | | | 0.181 |
| Stomach | CBCL_56F | 0.355 | 0.597 | | | 0.261 |
| Vomit | CBCL_56G | 0.318 | 0.601 | | | 0.219 |
| Other physical problem | CBCL_56H | 0.301 | 0.214 | | | 0.664 |
| Argues | CBCL_3 | 0.708 | | 0.255 | | 0.885 |
| Brags | CBCL_7 | 0.481 | | 0.306 | | 0.712 |
| Cruel to animals | CBCL_15 | 0.453 | | 0.403 | | 0.558 |

| | | | | |
|---|---|---|---|---|
| Mean | CBCL_16 | 0.584 | 0.555 | 0.525 |
| Demands a lot of attention | CBCL_19 | 0.717 | 0.166 | 0.949 |
| Destroys own things | CBCL_20 | 0.573 | 0.532 | 0.537 |
| Destroys other | CBCL_21 | 0.602 | 0.629 | 0.478 |
| Disobedient at home | CBCL_22 | 0.703 | 0.511 | 0.654 |
| Disobedient at school | CBCL_23 | 0.591 | 0.495 | 0.588 |
| Doesn’t get along with peers | CBCL_25 | 0.640 | 0.296 | 0.824 |
| No guilt | CBCL_26 | 0.603 | 0.400 | 0.694 |
| Jealous | CBCL_27 | 0.660 | 0.206 | 0.911 |
| Breaks rules at home | CBCL_28 | 0.704 | 0.516 | 0.651 |
| Unloved | CBCL_33 | 0.680 | -0.009 | 1.0 |
| Feels persecuted | CBCL_34 | 0.668 | 0.072 | 0.989 |
| Fights | CBCL_37 | 0.591 | 0.479 | 0.604 |
| Bad friends | CBCL_39 | 0.519 | 0.419 | 0.605 |

| | | | | |
|---|---|---|---|---|
| Lies or cheats | CBCL_43 | 0.583 | 0.448 | 0.629 |
| Attacks | CBCL_57 | 0.580 | 0.560 | 0.518 |
| Screams | CBCL_68 | 0.681 | 0.247 | 0.884 |
| Sets fires | CBCL_72 | 0.388 | 0.236 | 0.730 |
| Shows off | CBCL_74 | 0.554 | 0.294 | 0.780 |
| Steals from home | CBCL_81 | 0.454 | 0.596 | 0.367 |
| Steals outside home | CBCL_82 | 0.390 | 0.634 | 0.275 |
| Stubborn | CBCL_86 | 0.783 | 0.140 | 0.969 |
| Mood changes | CBCL_87 | 0.811 | -0.037 | 0.998 |
| Sulks | CBCL_88 | 0.725 | -0.098 | 0.982 |
| Suspicious | CBCL_89 | 0.640 | -0.024 | 0.999 |
| Swears | CBCL_90 | 0.548 | 0.303 | 0.766 |
| Teases | CBCL_94 | 0.593 | 0.360 | 0.731 |
| Temper | CBCL_95 | 0.740 | 0.248 | 0.899 |
| Threatens | CBCL_97 | 0.616 | 0.534 | 0.571 |

| | | | | | | |
|---|---|---|---|---|---|---|
| Vandalism | CBCL_106 | 0.524 | | 0.539 | | 0.486 |
| Whining | CBCL_109 | 0.621 | | 0.094 | | 0.978 |
| Acts too young | CBCL_1 | 0.527 | | | 0.261 | 0.803 |
| Fails to finish things | CBCL_4 | 0.699 | | | 0.385 | 0.767 |
| Can’t concentrate | CBCL_8 | 0.669 | | | 0.620 | 0.538 |
| Mind off | CBCL_9 | 0.670 | | | 0.187 | 0.928 |
| Can’t sit still | CBCL_10 | 0.636 | | | 0.343 | 0.775 |
| Confused | CBCL_13 | 0.600 | | | 0.385 | 0.708 |
| Daydreams | CBCL_17 | 0.563 | | | 0.525 | 0.535 |
| Gets hurt a lot | CBCL_36 | 0.574 | | | 0.179 | 0.912 |
| Impulsive | CBCL_41 | 0.813 | | | 0.166 | 0.960 |
| Twitches | CBCL_46 | 0.541 | | | 0.170 | 0.910 |
| Poor school work | CBCL_61 | 0.584 | | | 0.307 | 0.783 |
| Clumsy | CBCL_62 | 0.579 | | | 0.342 | 0.741 |
| Repeats acts | CBCL_66 | 0.600 | | | 0.199 | 0.901 |

| | | | | | | |
|---|---|---|---|---|---|---|
| Inattentive | CBCL_78 | 0.672 | | | 0.586 | 0.568 |
| Stares | CBCL_80 | 0.534 | | | 0.470 | 0.563 |
| Strange behavior | CBCL_84 | 0.655 | | | 0.125 | 0.965 |
| Strange ideas | CBCL_85 | 0.631 | | | 0.031 | 0.998 |
| Talks too much | CBCL_93 | 0.579 | | | 0.024 | 0.998 |
| Index | | | | | | |
| PUC | | 0.652 | | | | |
| ECV SS | | 0.679 | 0.480 | 0.286 | 0.232 | |
| ECV SG | | 0.679 | 0.122 | 0.142 | 0.058 | |
| ECV GS | | 0.679 | 0.520 | 0.714 | 0.768 | |
| Omega | | 0.956 | 0.860 | 0.932 | 0.875 | |
| OmegaH | | 0.844 | 0.380 | 0.206 | 0.155 | |
| H | | 0.977 | 0.863 | 0.876 | 0.736 | |
| FD | | 0.976 | 0.929 | 0.933 | 0.896 | |

Note: IECV, item explained common variance (≥ 0.85 yield unidimensional item sets that reflect the content of the general factor); ECV, explained common variance; SS, proportion of common variance of the items in each factor which is due to that factor; SG, ECV proportion of common variance of the items in each specific factor which is due to the specific factor; GS, proportion of common variance of the items in each specific factor which is due to the general factor; PUC, percent of uncontaminated correlations; OmegaH, omega-hierarchical; H, index of construct replicability (>0.8 suggests a well-defined latent variable); FD, factor determinacy (>0.9 indicate that the factor score can be used)

Table S5 - CBCL Bifactor model according to Moore et al., using four specific factors

| Content | Item | Factors and factor loadings | | | | | IECV |
|---|---|---|---|---|---|---|---|
| | | P-factor | Internalizing | Somatic | Externalizing | Attention | |
| Little they enjoy | CBCL_5 | 0.598 | 0.302 | | | | 0.797 |
| Fears school | CBCL_30 | 0.490 | 0.321 | | | | 0.700 |
| Fears do bad | CBCL_31 | 0.493 | 0.233 | | | | 0.817 |
| Perfect | CBCL_32 | 0.308 | 0.366 | | | | 0.415 |
| Worthless | CBCL_35 | 0.658 | 0.316 | | | | 0.813 |
| Prefers alone | CBCL_42 | 0.446 | 0.549 | | | | 0.398 |
| Fearful | CBCL_50 | 0.525 | 0.386 | | | | 0.649 |
| Feels too guilty | CBCL_52 | 0.527 | 0.381 | | | | 0.657 |
| Won’t talk | CBCL_65 | 0.507 | 0.398 | | | | 0.619 |
| Secretive | CBCL_69 | 0.494 | 0.432 | | | | 0.567 |
| Self-conscious | CBCL_71 | 0.530 | 0.464 | | | | 0.566 |

| | | | | | | |
|---|---|---|---|---|---|---|
| Shy | CBCL_75 | 0.277 | 0.571 | | | 0.191 |
| Lacks energy | CBCL_102 | 0.482 | 0.450 | | | 0.534 |
| Sad | CBCL_103 | 0.624 | 0.454 | | | 0.654 |
| Withdrawn | CBCL_111 | 0.473 | 0.662 | | | 0.338 |
| Worries | CBCL_112 | 0.499 | 0.457 | | | 0.544 |
| Dizzy | CBCL_51 | 0.433 | | 0.533 | | 0.398 |
| Aches | CBCL_56A | 0.453 | | 0.477 | | 0.474 |
| Headaches | CBCL_56B | 0.334 | | 0.656 | | 0.206 |
| Nausea | CBCL_56C | 0.424 | | 0.823 | | 0.210 |
| Stomach | CBCL_56F | 0.416 | | 0.696 | | 0.263 |
| Vomit | CBCL_56G | 0.377 | | 0.686 | | 0.232 |
| Other physical problem | CBCL_56H | 0.330 | | 0.159 | | 0.817 |
| Argues | CBCL_3 | 0.700 | | | 0.274 | 0.867 |
| Brags | CBCL_7 | 0.473 | | | 0.320 | 0.686 |

| | | | | |
|---|---|---|---|---|
| Cruel to animals | CBCL_15 | 0.445 | 0.410 | 0.541 |
| Mean | CBCL_16 | 0.573 | 0.567 | 0.505 |
| Demands a lot of attention | CBCL_19 | 0.711 | 0.186 | 0.936 |
| Destroys own things | CBCL_20 | 0.564 | 0.540 | 0.522 |
| Destroys other | CBCL_21 | 0.591 | 0.637 | 0.463 |
| Disobedient at home | CBCL_22 | 0.692 | 0.527 | 0.633 |
| Disobedient at school | CBCL_23 | 0.581 | 0.505 | 0.570 |
| Doesn't get along with peers | CBCL_25 | 0.632 | 0.309 | 0.807 |
| No guilt | CBCL_26 | 0.594 | 0.414 | 0.673 |
| Jealous | CBCL_27 | 0.651 | 0.227 | 0.892 |
| Breaks rules at home | CBCL_28 | 0.694 | 0.528 | 0.633 |
| Unloved | CBCL_33 | 0.675 | 0.016 | 0.999 |
| Feels persecuted | CBCL_34 | 0.663 | 0.092 | 0.981 |
| Fights | CBCL_37 | 0.580 | 0.494 | 0.580 |

| | | | | |
|---|---|---|---|---|
| Bad friends | CBCL_39 | 0.511 | 0.426 | 0.590 |
| Lies or cheats | CBCL_43 | 0.576 | 0.455 | 0.616 |
| Attacks | CBCL_57 | 0.569 | 0.572 | 0.496 |
| Screams | CBCL_68 | 0.672 | 0.268 | 0.863 |
| Sets fires | CBCL_72 | 0.383 | 0.243 | 0.713 |
| Shows off | CBCL_74 | 0.546 | 0.309 | 0.757 |
| Steals from home | CBCL_81 | 0.446 | 0.598 | 0.357 |
| Steals outside home | CBCL_82 | 0.381 | 0.634 | 0.265 |
| Stubborn | CBCL_86 | 0.775 | 0.167 | 0.956 |
| Mood changes | CBCL_87 | 0.806 | -0.009 | 1.0 |
| Sulks | CBCL_88 | 0.719 | -0.065 | 0.992 |
| Suspicious | CBCL_89 | 0.636 | 0.001 | 1.0 |
| Swears | CBCL_90 | 0.540 | 0.319 | 0.741 |
| Teases | CBCL_94 | 0.583 | 0.378 | 0.704 |
| Temper | CBCL_95 | 0.731 | 0.274 | 0.877 |

| | | | | | |
|---|---|---|---|---|---|
| Threatens | CBCL_97 | 0.605 | 0.549 | | 0.548 |
| Vandalism | CBCL_106 | 0.513 | 0.550 | | 0.465 |
| Whining | CBCL_109 | 0.616 | 0.112 | | 0.968 |
| Acts too young | CBCL_1 | 0.527 | | 0.260 | 0.804 |
| Fails to finish things | CBCL_4 | 0.699 | | 0.385 | 0.767 |
| Can’t concentrate | CBCL_8 | 0.669 | | 0.621 | 0.537 |
| Mind off | CBCL_9 | 0.671 | | 0.183 | 0.931 |
| Can’t sit still | CBCL_10 | 0.635 | | 0.344 | 0.773 |
| Confused | CBCL_13 | 0.602 | | 0.381 | 0.714 |
| Daydreams | CBCL_17 | 0.565 | | 0.523 | 0.539 |
| Gets hurt a lot | CBCL_36 | 0.575 | | 0.176 | 0.914 |
| Impulsive | CBCL_41 | 0.814 | | 0.165 | 0.961 |
| Twitches | CBCL_46 | 0.543 | | 0.165 | 0.915 |
| Poor school work | CBCL_61 | 0.584 | | 0.308 | 0.782 |
| Clumsy | CBCL_62 | 0.581 | | 0.338 | 0.747 |

| | | | | | | | |
|---|---|---|---|---|---|---|---|
| Repeats acts | CBCL_66 | 0.601 | | | | 0.194 | 0.906 |
| Inattentive | CBCL_78 | 0.673 | | | | 0.586 | 0.569 |
| Stares | CBCL_80 | 0.536 | | | | 0.466 | 0.570 |
| Strange behavior | CBCL_84 | 0.656 | | | | 0.120 | 0.968 |
| Strange ideas | CBCL_85 | 0.633 | | | | 0.025 | 0.998 |
| Talks too much | CBCL_93 | 0.580 | | | | 0.022 | 0.999 |
| Index | | | | | | | |
| PUC | | 0.692 | | | | | |
| ECV SS | | 0.653 | 0.426 | 0.701 | 0.302 | 0.229 | |
| ECV SG | | 0.653 | 0.080 | 0.068 | 0.144 | 0.055 | |
| ECV GS | | 0.653 | 0.574 | 0.299 | 0.698 | 0.771 | |
| Omega | | 0.957 | 0.842 | 0.759 | 0.932 | 0.875 | |
| OmegaH | | 0.851 | 0.340 | 0.492 | 0.225 | 0.153 | |
| H | | 0.977 | 0.802 | 0.844 | 0.883 | 0.734 | |
| FD | | 0.975 | 0.903 | 0.943 | 0.933 | 0.894 | |

Note: IECV, item explained common variance (≥ 0.85 yield unidimensional item sets that reflect the content of the general factor); ECV, explained common variance; SS, proportion of common variance of the items in each factor which is due to that factor; SG, ECV proportion of common variance of the items in each specific factor which is due to the specific factor; GS, proportion of common variance of the items in each specific factor which is due to the general factor; PUC, percent of uncontaminated correlations; OmegaH, omega-hierarchical; H, index of construct replicability (> 0.8 suggests a well-defined latent variable); FD, factor determinacy (> 0.9 indicate that the factor score can be used)

Table S6 - CBCL Bifactor model according to McElroy et al., using three specific factors

| Content | Item | Factors and factor loadings | | | | IECV |
|---|---|---|---|---|---|---|
| | | P-factor | Internalizing | Externalizing | Attention | |
| Cries | CBCL_14 | 0.546 | 0.172 | | | 0.910 |
| Fears | CBCL_29 | 0.343 | 0.299 | | | 0.568 |
| Fears school | CBCL_30 | 0.467 | 0.345 | | | 0.647 |
| Fears do bad | CBCL_31 | 0.469 | 0.263 | | | 0.761 |
| Perfect | CBCL_32 | 0.284 | 0.349 | | | 0.398 |
| Unloved | CBCL_33 | 0.656 | 0.185 | | | 0.926 |
| Worthless | CBCL_35 | 0.634 | 0.335 | | | 0.782 |
| Nervous | CBCL_45 | 0.650 | 0.365 | | | 0.760 |
| Fearful | CBCL_50 | 0.498 | 0.511 | | | 0.487 |
| Feels too guilty | CBCL_52 | 0.487 | 0.443 | | | 0.547 |
| Self-conscious | CBCL_71 | 0.499 | 0.413 | | | 0.593 |

| | | | | |
|---|---|---|---|---|
| Talks about suicide | CBCL_91 | 0.585 | 0.122 | 0.958 |
| Worries | CBCL_112 | 0.457 | 0.483 | 0.472 |
| Prefers alone | CBCL_42 | 0.430 | 0.413 | 0.520 |
| Won't talk | CBCL_65 | 0.518 | 0.279 | 0.775 |
| Secretive | CBCL_69 | 0.497 | 0.332 | 0.691 |
| Shy | CBCL_75 | 0.276 | 0.440 | 0.282 |
| Lacks energy | CBCL_102 | 0.454 | 0.424 | 0.534 |
| Sad | CBCL_103 | 0.614 | 0.430 | 0.671 |
| Withdrawn | CBCL_111 | 0.461 | 0.485 | 0.475 |
| Nightmares | CBCL_47 | 0.457 | 0.250 | 0.770 |
| Constipate | CBCL_49 | 0.307 | 0.285 | 0.537 |
| Dizzy | CBCL_51 | 0.356 | 0.570 | 0.281 |
| Tired | CBCL_54 | 0.527 | 0.408 | 0.625 |
| Aches | CBCL_56A | 0.378 | 0.474 | 0.389 |
| Headaches | CBCL_56B | 0.279 | 0.539 | 0.211 |

| | | | | | |
|---|---|---|---|---|---|
| Nausea | CBCL_56C | 0.320 | 0.738 | | 0.158 |
| Eye problems | CBCL_56D | 0.249 | 0.245 | | 0.508 |
| Skin problems | CBCL_56E | 0.272 | 0.227 | | 0.589 |
| Stomach | CBCL_56F | 0.336 | 0.608 | | 0.234 |
| Vomit | CBCL_56G | 0.303 | 0.601 | | 0.203 |
| No guilt | CBCL_26 | 0.596 | | 0.397 | 0.693 |
| Bad friends | CBCL_39 | 0.518 | | 0.418 | 0.606 |
| Lies or cheats | CBCL_43 | 0.585 | | 0.452 | 0.626 |
| Prefers older | CBCL_63 | 0.405 | | 0.105 | 0.937 |
| Sets fires | CBCL_72 | 0.406 | | 0.231 | 0.755 |
| Steals from home | CBCL_81 | 0.448 | | 0.620 | 0.343 |
| Steals outside home | CBCL_82 | 0.387 | | 0.653 | 0.260 |
| Swears | CBCL_90 | 0.577 | | 0.269 | 0.821 |
| Thinks about sex | CBCL_96 | 0.493 | | 0.149 | 0.916 |
| Argues | CBCL_3 | 0.707 | | 0.255 | 0.885 |

| | | | | |
|---|---|---|---|---|
| Mean | CBCL_16 | 0.581 | 0.540 | 0.537 |
| Demands a lot of attention | CBCL_19 | 0.717 | 0.148 | 0.959 |
| Destroys own things | CBCL_20 | 0.574 | 0.554 | 0.518 |
| Destroys other | CBCL_21 | 0.595 | 0.657 | 0.451 |
| Disobedient at home | CBCL_22 | 0.707 | 0.493 | 0.673 |
| Disobedient at school | CBCL_23 | 0.580 | 0.481 | 0.593 |
| Fights | CBCL_37 | 0.603 | 0.454 | 0.638 |
| Attacks | CBCL_57 | 0.583 | 0.556 | 0.524 |
| Screams | CBCL_68 | 0.695 | 0.249 | 0.886 |
| Stubborn | CBCL_86 | 0.810 | 0.105 | 0.983 |
| Mood changes | CBCL_87 | 0.842 | -0.070 | 0.993 |
| Sulks | CBCL_88 | 0.782 | -0.181 | 0.949 |
| Suspicious | CBCL_89 | 0.676 | -0.071 | 0.989 |
| Teases | CBCL_94 | 0.601 | 0.329 | 0.769 |

| | | | | | | |
|---|---|---|---|---|---|---|
| Temper | CBCL_95 | 0.762 | | 0.228 | | 0.918 |
| Threatens | CBCL_97 | 0.630 | | 0.519 | | 0.596 |
| Loud | CBCL_104 | 0.633 | | 0.233 | | 0.881 |
| Acts too young | CBCL_1 | 0.516 | | | 0.260 | 0.798 |
| Can't concentrate | CBCL_8 | 0.664 | | | 0.625 | 0.530 |
| Can't sit still | CBCL_10 | 0.623 | | | 0.361 | 0.749 |
| Confused | CBCL_13 | 0.618 | | | 0.393 | 0.712 |
| Daydreams | CBCL_17 | 0.571 | | | 0.520 | 0.547 |
| Impulsive | CBCL_41 | 0.793 | | | 0.192 | 0.945 |
| Poor school work | CBCL_61 | 0.581 | | | 0.292 | 0.798 |
| Stares | CBCL_80 | 0.547 | | | 0.483 | 0.562 |
| Index | | | | | | |
| | PUC | 0.607 | | | | |
| | ECV SS | 0.647 | 0.451 | 0.287 | 0.308 | |
| | ECV SG | 0.647 | 0.171 | 0.137 | 0.044 | |

| | | | | |
|---|---|---|---|---|
| ECV GS | 0.647 | 0.549 | 0.713 | 0.692 |
| Omega | 0.946 | 0.882 | 0.919 | 0.803 |
| OmegaH | 0.803 | 0.365 | 0.212 | 0.219 |
| H | 0.972 | 0.880 | 0.856 | 0.649 |
| FD | 0.975 | 0.933 | 0.930 | 0.873 |

Note: IECV, item explained common variance (≥ 0.85 yield unidimensional item sets that reflect the content of the general factor); ECV, explained common variance; SS, proportion of common variance of the items in each factor which is due to that factor; SG, ECV proportion of common variance of the items in each specific factor which is due to the specific factor; GS, proportion of common variance of the items in each specific factor which is due to the general factor; PUC, percent of uncontaminated correlations; OmegaH, omega-hierarchical; H, index of construct replicability (>0.8 suggests a well-defined latent variable); FD, factor determinacy (>0.9 indicate that the factor score can be used)

Table S7 - CBCL Bifactor model according to Deutz et al., using two specific factors (GP model)

| Content | Item | Factors and factor loadings | | | IECV |
|---|---|---|---|---|---|
| | | P-factor | Internalizing | Externalizing | |
| Cries | CBCL_14 | 0.580 | 0.076 | | 0.983 |
| Fears | CBCL_29 | 0.416 | 0.186 | | 0.833 |
| Fears school | CBCL_30 | 0.521 | 0.248 | | 0.815 |
| Fears do bad | CBCL_31 | 0.539 | 0.117 | | 0.955 |
| Perfect | CBCL_32 | 0.375 | 0.209 | | 0.763 |
| Unloved | CBCL_33 | 0.702 | 0.058 | | 0.993 |
| Worthless | CBCL_35 | 0.691 | 0.197 | | 0.925 |
| Nervous | CBCL_45 | 0.718 | 0.234 | | 0.904 |
| Fearful | CBCL_50 | 0.591 | 0.366 | | 0.723 |
| Feels too guilty | CBCL_52 | 0.575 | 0.297 | | 0.789 |
| Self-conscious | CBCL_71 | 0.552 | 0.324 | | 0.744 |

| | | | | |
|---|---|---|---|---|
| Talks about suicide | CBCL_91 | 0.635 | -0.011 | 1.0 |
| Worries | CBCL_112 | 0.548 | 0.349 | 0.711 |
| Prefers alone | CBCL_42 | 0.475 | 0.358 | 0.638 |
| Won’t talk | CBCL_65 | 0.551 | 0.213 | 0.870 |
| Secretive | CBCL_69 | 0.540 | 0.262 | 0.809 |
| Shy | CBCL_75 | 0.322 | 0.389 | 0.407 |
| Lacks energy | CBCL_102 | 0.481 | 0.395 | 0.597 |
| Sad | CBCL_103 | 0.680 | 0.316 | 0.822 |
| Withdrawn | CBCL_111 | 0.512 | 0.425 | 0.592 |
| Nightmares | CBCL_47 | 0.529 | 0.137 | 0.937 |
| Constipate | CBCL_49 | 0.345 | 0.247 | 0.661 |
| Dizzy | CBCL_51 | 0.411 | 0.550 | 0.358 |
| Tired | CBCL_54 | 0.570 | 0.353 | 0.723 |
| Aches | CBCL_56A | 0.426 | 0.456 | 0.466 |
| Headaches | CBCL_56B | 0.317 | 0.559 | 0.243 |

| | | | | | |
|---|---|---|---|---|---|
| Nausea | CBCL_56C | 0.365 | 0.772 | | 0.183 |
| Eye problems | CBCL_56D | 0.282 | 0.221 | | 0.620 |
| Skin problems | CBCL_56E | 0.301 | 0.197 | | 0.700 |
| Stomach | CBCL_56F | 0.369 | 0.636 | | 0.252 |
| Vomit | CBCL_56G | 0.336 | 0.628 | | 0.223 |
| Argues | CBCL_3 | 0.623 | | 0.412 | 0.696 |
| Mean | CBCL_16 | 0.497 | | 0.613 | 0.397 |
| Demands a lot of attention | CBCL_19 | 0.655 | | 0.295 | 0.831 |
| Destroys own things | CBCL_20 | 0.495 | | 0.610 | 0.397 |
| Destroys other | CBCL_21 | 0.502 | | 0.711 | 0.333 |
| Disobedient at home | CBCL_22 | 0.578 | | 0.651 | 0.441 |
| Disobedient at school | CBCL_23 | 0.425 | | 0.639 | 0.307 |
| Fights | CBCL_37 | 0.509 | | 0.561 | 0.452 |
| Attacks | CBCL_57 | 0.504 | | 0.625 | 0.394 |
| Screams | CBCL_68 | 0.625 | | 0.398 | 0.711 |

| | | | | |
|---|---|---|---|---|
| Stubborn | CBCL_86 | 0.748 | 0.299 | 0.862 |
| Mood changes | CBCL_87 | 0.804 | 0.127 | 0.976 |
| Sulks | CBCL_88 | 0.744 | 0.046 | 0.996 |
| Suspicious | CBCL_89 | 0.665 | 0.067 | 0.990 |
| Teases | CBCL_94 | 0.529 | 0.444 | 0.587 |
| Temper | CBCL_95 | 0.683 | 0.406 | 0.739 |
| Threatens | CBCL_97 | 0.552 | 0.602 | 0.457 |
| Loud | CBCL_104 | 0.550 | 0.383 | 0.673 |
| No guilt | CBCL_26 | 0.517 | 0.496 | 0.521 |
| Bad friends | CBCL_39 | 0.427 | 0.502 | 0.420 |
| Lies or cheats | CBCL_43 | 0.488 | 0.538 | 0.451 |
| Prefers older | CBCL_63 | 0.381 | 0.176 | 0.824 |
| Steals from home | CBCL_81 | 0.386 | 0.625 | 0.276 |
| Steals outside home | CBCL_82 | 0.330 | 0.641 | 0.210 |
| Swears | CBCL_90 | 0.514 | 0.381 | 0.645 |

| | | | | |
|---|---|---|---|---|
| Thinks about sex | CBCL_96 | 0.478 | 0.210 | 0.838 |
| Mind off | CBCL_9 | 0.700 | | 1.0 |
| Harms self | CBCL_18 | 0.569 | | 1.0 |
| Hears things | CBCL_40 | 0.586 | | 1.0 |
| Twitches | CBCL_46 | 0.588 | | 1.0 |
| Picks skin | CBCL_58 | 0.508 | | 1.0 |
| Sex parts | CBCL_60 | 0.502 | | 1.0 |
| Repeats acts | CBCL_66 | 0.630 | | 1.0 |
| Sees things | CBCL_70 | 0.584 | | 1.0 |
| Sleeps less | CBCL_76 | 0.540 | | 1.0 |
| Stores up | CBCL_83 | 0.549 | | 1.0 |
| Strange behavior | CBCL_84 | 0.682 | | 1.0 |
| Strange ideas | CBCL_85 | 0.668 | | 1.0 |
| Sleep walks | CBCL_92 | 0.385 | | 1.0 |
| Sleep problems | CBCL_100 | 0.593 | | 1.0 |

Index

| | | | |
|---|---|---|---|
| PUC | 0.682 | | |
| ECV SS | 0.675 | 0.336 | 0.425 |
| ECV SG | 0.675 | 0.131 | 0.194 |
| ECV GS | 0.675 | 0.664 | 0.575 |
| Omega | 0.944 | 0.882 | 0.920 |
| OmegaH | 0.802 | 0.235 | 0.346 |
| H | 0.971 | 0.857 | 0.902 |
| FD | 0.977 | 0.925 | 0.950 |

Note: GP, General Psychopathology; IECV, item explained common variance (≥ 0.85 yield unidimensional item sets that reflect the content of the general factor); ECV, explained common variance; SS, proportion of common variance of the items in each factor which is due to that factor; SG, ECV proportion of common variance of the items in each specific factor which is due to the specific factor; GS, proportion of common variance of the items in each specific factor which is due to the general factor; PUC, percent of uncontaminated correlations; OmegaH, omega-hierarchical; H, index of construct replicability (>0.8 suggests a well-defined latent variable); FD, factor determinacy (>0.9 indicate that the factor score can be used)

Table S8 - CBCL Bifactor model according to Deutz et al. and Haltigan et al., using three specific factors (DP model)

| Content | Item | Factors and factor loadings | | | | IECV |
|---|---|---|---|---|---|---|
| | | P-factor | Anxious-Depressed | Aggressive behavior | Attention problems | |
| Cries | CBCL_14 | 0.549 | 0.202 | | | 0.881 |
| Fears | CBCL_29 | 0.335 | 0.388 | | | 0.427 |
| Fears school | CBCL_30 | 0.451 | 0.399 | | | 0.561 |
| Fears do bad | CBCL_31 | 0.443 | 0.451 | | | 0.491 |
| Perfect | CBCL_32 | 0.264 | 0.531 | | | 0.198 |
| Unloved | CBCL_33 | 0.644 | 0.248 | | | 0.871 |
| Worthless | CBCL_35 | 0.606 | 0.428 | | | 0.667 |
| Nervous | CBCL_45 | 0.640 | 0.432 | | | 0.687 |
| Fearful | CBCL_50 | 0.488 | 0.632 | | | 0.374 |
| Feels too guilty | CBCL_52 | 0.462 | 0.569 | | | 0.397 |
| Self-conscious | CBCL_71 | 0.483 | 0.414 | | | 0.576 |

| | | | | | |
|---|---|---|---|---|---|
| Talks about suicide | CBCL_91 | 0.558 | 0.185 | | 0.901 |
| Worries | CBCL_112 | 0.436 | 0.591 | | 0.352 |
| Argues | CBCL_3 | 0.736 | | 0.176 | 0.946 |
| Mean | CBCL_16 | 0.622 | | 0.480 | 0.627 |
| Demands a lot of attention | CBCL_19 | 0.741 | | 0.092 | 0.985 |
| Destroys own things | CBCL_20 | 0.612 | | 0.557 | 0.547 |
| Destroys other | CBCL_21 | 0.639 | | 0.653 | 0.489 |
| Disobedient at home | CBCL_22 | 0.762 | | 0.406 | 0.779 |
| Disobedient at school | CBCL_23 | 0.639 | | 0.401 | 0.717 |
| Fights | CBCL_37 | 0.644 | | 0.369 | 0.753 |
| Attacks | CBCL_57 | 0.637 | | 0.525 | 0.595 |
| Screams | CBCL_68 | 0.732 | | 0.178 | 0.944 |
| Stubborn | CBCL_86 | 0.820 | | 0.015 | 1.0 |
| Mood changes | CBCL_87 | 0.821 | | -0.129 | 0.976 |

| | | | | | |
|---|---|---|---|---|---|
| Sulks | CBCL_88 | 0.766 | -0.283 | | 0.880 |
| Suspicious | CBCL_89 | 0.652 | -0.188 | | 0.923 |
| Teases | CBCL_94 | 0.632 | 0.238 | | 0.876 |
| Temper | CBCL_95 | 0.797 | 0.151 | | 0.965 |
| Threatens | CBCL_97 | 0.675 | 0.475 | | 0.669 |
| Loud | CBCL_104 | 0.676 | 0.165 | | 0.944 |
| Acts too young | CBCL_1 | 0.509 | | 0.280 | 0.768 |
| Can’t concentrate | CBCL_8 | 0.645 | | 0.616 | 0.523 |
| Can’t sit still | CBCL_10 | 0.648 | | 0.331 | 0.793 |
| Confused | CBCL_13 | 0.550 | | 0.490 | 0.558 |
| Daydreams | CBCL_17 | 0.513 | | 0.602 | 0.421 |
| Impulsive | CBCL_41 | 0.782 | | 0.226 | 0.923 |
| Poor school work | CBCL_61 | 0.544 | | 0.344 | 0.714 |
| Stares | CBCL_80 | 0.480 | | 0.572 | 0.413 |

Index

| | | | | |
|---|---|---|---|---|
| PUC | 0.650 | | | |
| ECV SS | 0.699 | 0.438 | 0.201 | 0.373 |
| ECV SG | 0.699 | 0.119 | 0.105 | 0.078 |
| ECV GS | 0.699 | 0.562 | 0.799 | 0.627 |
| Omega | 0.935 | 0.811 | 0.906 | 0.802 |
| OmegaH | 0.843 | 0.341 | 0.093 | 0.274 |
| H | 0.967 | 0.778 | 0.752 | 0.704 |
| FD | 0.976 | 0.897 | 0.914 | 0.886 |

Note: DP, Dysregulation profile; IECV, item explained common variance (≥ 0.85 yield unidimensional item sets that reflect the content of the general factor); ECV, explained common variance; SS, proportion of common variance of the items in each factor which is due to that factor; SG, ECV proportion of common variance of the items in each specific factor which is due to the specific factor; GS, proportion of common variance of the items in each specific factor which is due to the general factor; PUC, percent of uncontaminated correlations; OmegaH, omega-hierarchical; H, index of construct replicability (>0.8 suggests a well-defined latent variable); FD, factor determinacy (>0.9 indicate that the factor score can be used)

Table S9 - CBCL Bifactor model according to Haltigan et al., using four specific factors

| Content | Item | Factors and factor loadings | | | | | IECV |
|---|---|---|---|---|---|---|---|
| | | P-factor | Internalizing | Externalizing | Thought | Attention | |
| Cries | CBCL_14 | 0.554 | 0.137 | | | | 0.942 |
| Fears | CBCL_29 | 0.379 | 0.250 | | | | 0.697 |
| Fears school | CBCL_30 | 0.496 | 0.299 | | | | 0.733 |
| Fears do bad | CBCL_31 | 0.496 | 0.210 | | | | 0.848 |
| Perfect | CBCL_32 | 0.320 | 0.302 | | | | 0.529 |
| Unloved | CBCL_33 | 0.653 | 0.156 | | | | 0.946 |
| Worthless | CBCL_35 | 0.655 | 0.283 | | | | 0.843 |
| Nervous | CBCL_45 | 0.679 | 0.310 | | | | 0.828 |
| Fearful | CBCL_50 | 0.540 | 0.454 | | | | 0.586 |
| Feels too guilty | CBCL_52 | 0.530 | 0.383 | | | | 0.657 |
| Self-conscious | CBCL_71 | 0.522 | 0.376 | | | | 0.658 |

| | | | | |
|---|---|---|---|---|
| Talks about suicide | CBCL_91 | 0.596 | 0.076 | 0.984 |
| Worries | CBCL_112 | 0.507 | 0.421 | 0.592 |
| Prefers alone | CBCL_42 | 0.466 | 0.366 | 0.618 |
| Won’t talk | CBCL_65 | 0.533 | 0.245 | 0.826 |
| Secretive | CBCL_69 | 0.513 | 0.301 | 0.744 |
| Shy | CBCL_75 | 0.301 | 0.415 | 0.345 |
| Lacks energy | CBCL_102 | 0.491 | 0.380 | 0.625 |
| Sad | CBCL_103 | 0.638 | 0.385 | 0.733 |
| Withdrawn | CBCL_111 | 0.505 | 0.430 | 0.580 |
| Nightmares | CBCL_47 | 0.499 | 0.191 | 0.872 |
| Constipate | CBCL_49 | 0.333 | 0.259 | 0.623 |
| Dizzy | CBCL_51 | 0.391 | 0.554 | 0.332 |
| Tired | CBCL_54 | 0.554 | 0.373 | 0.688 |
| Aches | CBCL_56A | 0.406 | 0.460 | 0.438 |
| Headaches | CBCL_56B | 0.294 | 0.554 | 0.220 |

| | | | | | | |
|---|---|---|---|---|---|---|
| Nausea | CBCL_56C | 0.351 | 0.748 | | | 0.180 |
| Eye problems | CBCL_56D | 0.277 | 0.221 | | | 0.611 |
| Skin problems | CBCL_56E | 0.296 | 0.201 | | | 0.684 |
| Stomach | CBCL_56F | 0.358 | 0.617 | | | 0.252 |
| Vomit | CBCL_56G | 0.325 | 0.610 | | | 0.221 |
| Argues | CBCL_3 | 0.677 | | 0.315 | | 0.822 |
| Mean | CBCL_16 | 0.548 | | 0.573 | | 0.478 |
| Demands a lot of attention | CBCL_19 | 0.694 | | 0.198 | | 0.925 |
| Destroys own things | CBCL_20 | 0.557 | | 0.558 | | 0.499 |
| Destroys other | CBCL_21 | 0.573 | | 0.662 | | 0.428 |
| Disobedient at home | CBCL_22 | 0.667 | | 0.546 | | 0.599 |
| Disobedient at school | CBCL_23 | 0.542 | | 0.519 | | 0.522 |
| Fights | CBCL_37 | 0.562 | | 0.514 | | 0.545 |
| Attacks | CBCL_57 | 0.553 | | 0.588 | | 0.469 |

| | | | | |
|---|---|---|---|---|
| Screams | CBCL_68 | 0.666 | 0.309 | 0.823 |
| Stubborn | CBCL_86 | 0.771 | 0.204 | 0.935 |
| Mood changes | CBCL_87 | 0.812 | 0.030 | 0.999 |
| Sulks | CBCL_88 | 0.733 | -0.036 | 0.998 |
| Suspicious | CBCL_89 | 0.648 | 0.021 | 0.999 |
| Teases | CBCL_94 | 0.563 | 0.397 | 0.668 |
| Temper | CBCL_95 | 0.720 | 0.314 | 0.840 |
| Threatens | CBCL_97 | 0.593 | 0.566 | 0.523 |
| Loud | CBCL_104 | 0.614 | 0.275 | 0.833 |
| No guilt | CBCL_26 | 0.570 | 0.433 | 0.634 |
| Bad friends | CBCL_39 | 0.493 | 0.444 | 0.552 |
| Lies or cheats | CBCL_43 | 0.559 | 0.475 | 0.581 |
| Prefers older | CBCL_63 | 0.394 | 0.135 | 0.895 |
| Runs away | CBCL_67 | 0.464 | 0.345 | 0.644 |
| Sets fires | CBCL_72 | 0.401 | 0.261 | 0.702 |

| | | | | | |
|---|---|---|---|---|---|
| Sex problems | CBCL_73 | 0.422 | 0.199 | | 0.818 |
| Steals from home | CBCL_81 | 0.439 | 0.613 | | 0.339 |
| Steals outside home | CBCL_82 | 0.381 | 0.639 | | 0.262 |
| Swears | CBCL_90 | 0.542 | 0.339 | | 0.719 |
| Thinks about sex | CBCL_96 | 0.494 | 0.207 | | 0.851 |
| Truants | CBCL_101 | 0.482 | 0.149 | | 0.913 |
| Use drugs | CBCL_105 | 0.312 | 0.283 | | 0.549 |
| Vandalism | CBCL_106 | 0.524 | 0.547 | | 0.479 |
| Mind off | CBCL_9 | 0.697 | | 0.141 | 0.961 |
| Harms self | CBCL_18 | 0.556 | | -0.103 | 0.967 |
| Hears things | CBCL_40 | 0.528 | | 0.530 | 0.498 |
| Twitches | CBCL_46 | 0.577 | | 0.151 | 0.936 |
| Picks skin | CBCL_58 | 0.516 | | 0.051 | 0.990 |
| Sex parts | CBCL_60 | 0.499 | | 0.098 | 0.963 |
| Repeats acts | CBCL_66 | 0.624 | | 0.189 | 0.916 |

| | | | | | |
|---|---|---|---|---|---|
| Sees things | CBCL_70 | 0.525 | 0.511 | | 0.514 |
| Sleeps less | CBCL_76 | 0.500 | 0.456 | | 0.546 |
| Stores up | CBCL_83 | 0.526 | 0.118 | | 0.952 |
| Strange behavior | CBCL_84 | 0.668 | 0.262 | | 0.867 |
| Strange ideas | CBCL_85 | 0.637 | 0.334 | | 0.784 |
| Sleep walks | CBCL_92 | 0.364 | 0.108 | | 0.919 |
| Sleep problems | CBCL_100 | 0.556 | 0.416 | | 0.641 |
| Acts too young | CBCL_1 | 0.522 | | 0.249 | 0.815 |
| Can’t concentrate | CBCL_8 | 0.668 | | 0.640 | 0.521 |
| Can’t sit still | CBCL_10 | 0.622 | | 0.367 | 0.742 |
| Confused | CBCL_13 | 0.635 | | 0.358 | 0.759 |
| Daydreams | CBCL_17 | 0.592 | | 0.480 | 0.603 |
| Impulsive | CBCL_41 | 0.789 | | 0.192 | 0.944 |
| Poor school work | CBCL_61 | 0.576 | | 0.298 | 0.789 |
| Stares | CBCL_80 | 0.572 | | 0.439 | 0.629 |

| Index | | | | | |
|---|---|---|---|---|---|
| PUC | 0.697 | | | | |
| ECV SS | 0.669 | 0.390 | 0.338 | 0.218 | 0.290 |
| ECV SG | 0.669 | 0.122 | 0.143 | 0.033 | 0.034 |
| ECV GS | 0.669 | 0.610 | 0.662 | 0.782 | 0.710 |
| Omega | 0.952 | 0.882 | 0.925 | 0.784 | 0.803 |
| OmegaH | 0.839 | 0.301 | 0.266 | 0.106 | 0.203 |
| H | 0.976 | 0.866 | 0.884 | 0.607 | 0.635 |
| FD | 0.976 | 0.926 | 0.937 | 0.814 | 0.877 |

Note: IECV, item explained common variance (≥ 0.85 yield unidimensional item sets that reflect the content of the general factor); ECV, explained common variance; SS, proportion of common variance of the items in each factor which is due to that factor; SG, ECV proportion of common variance of the items in each specific factor which is due to the specific factor; GS, proportion of common variance of the items in each specific factor which is due to the general factor; PUC, percent of uncontaminated correlations; OmegaH, omega-hierarchical; H, index of construct replicability (>0.8 suggests a well-defined latent variable); FD, factor determinacy (>0.9 indicate that the factor score can be used)

Table S10 - CBCL bifactor model according to Clark et al., using two specific factors

| Content | Item | Factors and factor loadings | | | IECV |
|---|---|---|---|---|---|
| | | P-factor | Internalizing | Externalizing | |
| Lonely | CBCL_12 | 0.552 | 0.269 | | 0.808 |
| Cries | CBCL_14 | 0.542 | 0.168 | | 0.912 |
| Fears do bad | CBCL_31 | 0.481 | 0.232 | | 0.811 |
| Perfect | CBCL_32 | 0.303 | 0.324 | | 0.467 |
| Unloved | CBCL_33 | 0.626 | 0.245 | | 0.867 |
| Feels persecuted | CBCL_34 | 0.640 | 0.185 | | 0.923 |
| Worthless | CBCL_35 | 0.627 | 0.353 | | 0.759 |
| Prefers alone | CBCL_42 | 0.448 | 0.399 | | 0.558 |
| Nervous | CBCL_45 | 0.638 | 0.370 | | 0.748 |
| Fearful | CBCL_50 | 0.519 | 0.453 | | 0.568 |
| Dizzy | CBCL_51 | 0.378 | 0.546 | | 0.324 |

| | | | | |
|---|---|---|---|---|
| Feels too guilty | CBCL_52 | 0.503 | 0.426 | 0.582 |
| Tired | CBCL_54 | 0.532 | 0.409 | 0.629 |
| Aches | CBCL_56A | 0.401 | 0.451 | 0.442 |
| Headaches | CBCL_56B | 0.282 | 0.553 | 0.206 |
| Nausea | CBCL_56C | 0.352 | 0.712 | 0.196 |
| Eye problems | CBCL_56D | 0.278 | 0.214 | 0.628 |
| Skin problems | CBCL_56E | 0.302 | 0.173 | 0.753 |
| Stomach | CBCL_56F | 0.361 | 0.579 | 0.280 |
| Vomit | CBCL_56G | 0.330 | 0.576 | 0.247 |
| Won’t talk | CBCL_65 | 0.494 | 0.317 | 0.708 |
| Secretive | CBCL_69 | 0.475 | 0.374 | 0.617 |
| Self-conscious | CBCL_71 | 0.511 | 0.398 | 0.622 |
| Shy | CBCL_75 | 0.286 | 0.438 | 0.299 |
| Stares | CBCL_80 | 0.571 | 0.147 | 0.938 |
| Sulks | CBCL_88 | 0.641 | 0.274 | 0.846 |

| | | | | | |
|---|---|---|---|---|---|
| Suspicious | CBCL_89 | 0.577 | 0.260 | | 0.831 |
| Lacks energy | CBCL_102 | 0.474 | 0.410 | | 0.572 |
| Sad | CBCL_103 | 0.594 | 0.460 | | 0.625 |
| Withdrawn | CBCL_111 | 0.478 | 0.477 | | 0.501 |
| Worries | CBCL_112 | 0.486 | 0.432 | | 0.559 |
| Argues | CBCL_3 | 0.703 | | 0.237 | 0.898 |
| Brags | CBCL_7 | 0.467 | | 0.378 | 0.604 |
| Mean | CBCL_16 | 0.603 | | 0.520 | 0.574 |
| Demands a lot of attention | CBCL_19 | 0.711 | | 0.180 | 0.940 |
| Destroys own things | CBCL_20 | 0.598 | | 0.514 | 0.575 |
| Destroys other | CBCL_21 | 0.632 | | 0.599 | 0.527 |
| Disobedient at school | CBCL_23 | 0.638 | | 0.316 | 0.803 |
| No guilt | CBCL_26 | 0.611 | | 0.369 | 0.733 |
| Jealous | CBCL_27 | 0.653 | | 0.211 | 0.905 |
| Fights | CBCL_37 | 0.614 | | 0.452 | 0.649 |

| | | | | |
|---|---|---|---|---|
| Bad friends | CBCL_39 | 0.530 | 0.399 | 0.638 |
| Lies or cheats | CBCL_43 | 0.599 | 0.415 | 0.676 |
| Attacks | CBCL_57 | 0.608 | 0.514 | 0.583 |
| Prefers older | CBCL_63 | 0.407 | 0.143 | 0.890 |
| Runs away | CBCL_67 | 0.477 | 0.295 | 0.723 |
| Screams | CBCL_68 | 0.680 | 0.258 | 0.874 |
| Sets fires | CBCL_72 | 0.403 | 0.243 | 0.733 |
| Shows off | CBCL_74 | 0.541 | 0.365 | 0.687 |
| Steals from home | CBCL_81 | 0.485 | 0.563 | 0.426 |
| Steals outside home | CBCL_82 | 0.431 | 0.601 | 0.340 |
| Stubborn | CBCL_86 | 0.760 | 0.157 | 0.959 |
| Mood changes | CBCL_87 | 0.776 | 0.013 | 1.0 |
| Swears | CBCL_90 | 0.546 | 0.324 | 0.740 |
| Talks too much | CBCL_93 | 0.554 | 0.199 | 0.886 |
| Teases | CBCL_94 | 0.584 | 0.428 | 0.651 |

| | | | | |
|---|---|---|---|---|
| Temper | CBCL_95 | 0.728 | 0.266 | 0.882 |
| Threatens | CBCL_97 | 0.633 | 0.518 | 0.599 |
| Truants | CBCL_101 | 0.461 | 0.098 | 0.957 |
| Loud | CBCL_104 | 0.634 | 0.272 | 0.845 |
| Acts too young | CBCL_1 | 0.559 | | 1.0 |
| Fails to finish things | CBCL_4 | 0.734 | | 1.0 |
| Little they enjoy | CBCL_5 | 0.622 | | 1.0 |
| Bowel movements outside toilet | CBCL_6 | 0.362 | | 1.0 |
| Can’t concentrate | CBCL_8 | 0.760 | | 1.0 |
| Mind off | CBCL_9 | 0.688 | | 1.0 |
| Can’t sit still | CBCL_10 | 0.668 | | 1.0 |
| Too dependent | CBCL_11 | 0.548 | | 1.0 |
| Confused | CBCL_13 | 0.650 | | 1.0 |
| Cruel to animals | CBCL_15 | 0.555 | | 1.0 |
| Daydreams | CBCL_17 | 0.632 | | 1.0 |

| | | | |
|---|---|---|---|
| Harms self | CBCL_18 | 0.521 | 1.0 |
| Disobedient at home | CBCL_22 | 0.805 | 1.0 |
| Doesn’t eat well | CBCL_24 | 0.423 | 1.0 |
| Doesn’t get along with peers | CBCL_25 | 0.707 | 1.0 |
| breaks rules at home | CBCL_28 | 0.804 | 1.0 |
| Fears | CBCL_29 | 0.410 | 1.0 |
| Fears school | CBCL_30 | 0.533 | 1.0 |
| Gets hurt a lot | CBCL_36 | 0.596 | 1.0 |
| Gets teased | CBCL_38 | 0.623 | 1.0 |
| Hears things | CBCL_40 | 0.531 | 1.0 |
| Impulsive | CBCL_41 | 0.799 | 1.0 |
| Bites fingernails | CBCL_44 | 0.357 | 1.0 |
| Twitches | CBCL_46 | 0.564 | 1.0 |
| Nightmares | CBCL_47 | 0.520 | 1.0 |
| Not liked by peers | CBCL_48 | 0.694 | 1.0 |

| | | | |
|---|---|---|---|
| Constipate | CBCL_49 | 0.373 | 1.0 |
| Overeatin | CBCL_53 | 0.459 | 1.0 |
| Overweight | CBCL_55 | 0.285 | 1.0 |
| Other physical problem | CBCL_56H | 0.342 | 1.0 |
| Picks skin | CBCL_58 | 0.521 | 1.0 |
| Plays with sex parts in public | CBCL_59 | 0.469 | 1.0 |
| Sex parts | CBCL_60 | 0.506 | 1.0 |
| Poor school work | CBCL_61 | 0.605 | 1.0 |
| Clumsy | CBCL_62 | 0.627 | 1.0 |
| Prefers younger kids | CBCL_64 | 0.405 | 1.0 |
| Repeats acts | CBCL_66 | 0.617 | 1.0 |
| Sees things | CBCL_70 | 0.529 | 1.0 |
| Sex problems | CBCL_73 | 0.444 | 1.0 |
| Sleeps less | CBCL_76 | 0.501 | 1.0 |
| Sleep more than most kids | CBCL_77 | 0.338 | 1.0 |

| | | | |
|---|---|---|---|
| Inattentive | CBCL_78 | 0.756 | 1.0 |
| Speech problem | CBCL_79 | 0.351 | 1.0 |
| Stores up | CBCL_83 | 0.521 | 1.0 |
| Strange behavior | CBCL_84 | 0.658 | 1.0 |
| Strange ideas | CBCL_85 | 0.628 | 1.0 |
| Talks about suicide | CBCL_91 | 0.581 | 1.0 |
| Sleep walks | CBCL_92 | 0.356 | 1.0 |
| Thinks about sex | CBCL_96 | 0.517 | 1.0 |
| Thumb sucking | CBCL_98 | 0.271 | 1.0 |
| Sleep problems | CBCL_100 | 0.550 | 1.0 |
| Vandalism | CBCL_106 | 0.663 | 1.0 |
| Wets self during day | CBCL_107 | 0.420 | 1.0 |
| Wets the bed | CBCL_108 | 0.325 | 1.0 |
| Whining | CBCL_109 | 0.630 | 1.0 |
| Wishes to be opposite sex | CBCL_110 | 0.409 | 1.0 |

Index

| | | | |
|---|---|---|---|
| PUC | 0.869 | | |
| ECV SS | 0.797 | 0.401 | 0.280 |
| ECV SG | 0.797 | 0.112 | 0.091 |
| ECV GS | 0.797 | 0.599 | 0.720 |
| Omega | 0.961 | 0.893 | 0.918 |
| OmegaH | 0.900 | 0.317 | 0.198 |
| H | 0.983 | 0.869 | 0.840 |
| FD | 0.988 | 0.938 | 0.933 |

Note: IECV, item explained common variance (≥ 0.85 yield unidimensional item sets that reflect the content of the general factor); ECV, explained common variance; SS, proportion of common variance of the items in each factor which is due to that factor; SG, ECV proportion of common variance of the items in each specific factor which is due to the specific factor; GS, proportion of common variance of the items in each specific factor which is due to the general factor; PUC, percent of uncontaminated correlations; OmegaH, omega-hierarchical; H, index of construct replicability (>0.8 suggests a well-defined latent variable); FD, factor determinacy (>0.9 indicate that the factor score can be used)

Table S11 - CBCL bifactor model according to Clark et al., using three specific factors (EFA-generated)

| Content | Item | Factors and factor loadings | | | | IECV |
|---|---|---|---|---|---|---|
| | | P-factor | Internalizing | Externalizing | Attention | |
| Little they enjoy | CBCL_5 | 0.568 | 0.321 | | | 0.758 |
| Too dependent | CBCL_11 | 0.529 | 0.159 | | | 0.917 |
| Lonely | CBCL_12 | 0.551 | 0.283 | | | 0.791 |
| Cries | CBCL_14 | 0.545 | 0.184 | | | 0.898 |
| Harms self | CBCL_18 | 0.508 | 0.126 | | | 0.942 |
| Fears | CBCL_29 | 0.364 | 0.277 | | | 0.633 |
| Fears school | CBCL_30 | 0.474 | 0.330 | | | 0.674 |
| Fears do bad | CBCL_31 | 0.481 | 0.243 | | | 0.797 |
| Perfect | CBCL_32 | 0.296 | 0.341 | | | 0.430 |
| Unloved | CBCL_33 | 0.635 | 0.232 | | | 0.882 |
| Feels persecuted | CBCL_34 | 0.653 | 0.163 | | | 0.941 |

| | | | | |
|---|---|---|---|---|
| Worthless | CBCL_35 | 0.627 | 0.353 | 0.759 |
| Prefers alone | CBCL_42 | 0.440 | 0.405 | 0.541 |
| Nervous | CBCL_45 | 0.638 | 0.375 | 0.743 |
| Nightmares | CBCL_47 | 0.488 | 0.221 | 0.830 |
| Constipate | CBCL_49 | 0.328 | 0.259 | 0.618 |
| Fearful | CBCL_50 | 0.502 | 0.503 | 0.499 |
| Dizzy | CBCL_51 | 0.360 | 0.568 | 0.287 |
| Feels too guilty | CBCL_52 | 0.497 | 0.433 | 0.568 |
| Overeatin | CBCL_53 | 0.440 | 0.153 | 0.892 |
| Tired | CBCL_54 | 0.525 | 0.421 | 0.609 |
| Won’t talk | CBCL_65 | 0.493 | 0.312 | 0.714 |
| Secretive | CBCL_69 | 0.475 | 0.358 | 0.636 |
| Sees things | CBCL_70 | 0.499 | 0.207 | 0.853 |
| Self-conscious | CBCL_71 | 0.509 | 0.393 | 0.627 |
| Shy | CBCL_75 | 0.274 | 0.446 | 0.274 |

| | | | | |
|---|---|---|---|---|
| Sleeps less | CBCL_76 | 0.476 | 0.181 | 0.874 |
| Mood changes | CBCL_87 | 0.753 | 0.219 | 0.922 |
| Sulks | CBCL_88 | 0.649 | 0.257 | 0.864 |
| Talks about suicide | CBCL_91 | 0.563 | 0.152 | 0.932 |
| Sleep problems | CBCL_100 | 0.514 | 0.234 | 0.828 |
| Lacks energy | CBCL_102 | 0.464 | 0.420 | 0.550 |
| Sad | CBCL_103 | 0.582 | 0.485 | 0.590 |
| Whining | CBCL_109 | 0.651 | -0.041 | 0.996 |
| Withdrawn | CBCL_111 | 0.468 | 0.482 | 0.485 |
| Worries | CBCL_112 | 0.476 | 0.453 | 0.525 |
| Aches | CBCL_56A | 0.393 | 0.451 | 0.432 |
| Headaches | CBCL_56B | 0.271 | 0.545 | 0.198 |
| Nausea | CBCL_56C | 0.333 | 0.711 | 0.180 |
| Eye problems | CBCL_56D | 0.276 | 0.215 | 0.622 |
| Stomach | CBCL_56F | 0.345 | 0.586 | 0.257 |

| | | | | | |
|---|---|---|---|---|---|
| Vomit | CBCL_56G | 0.315 | 0.579 | | 0.228 |
| Other physical problem | CBCL_56H | 0.307 | 0.203 | | 0.701 |
| Argues | CBCL_3 | 0.708 | | 0.250 | 0.889 |
| Brags | CBCL_7 | 0.485 | | 0.294 | 0.731 |
| Cruel to animals | CBCL_15 | 0.486 | | 0.364 | 0.641 |
| Mean | CBCL_16 | 0.586 | | 0.551 | 0.531 |
| Demands a lot of attention | CBCL_19 | 0.731 | | 0.132 | 0.968 |
| Destroys own things | CBCL_20 | 0.594 | | 0.504 | 0.581 |
| Destroys other | CBCL_21 | 0.617 | | 0.606 | 0.509 |
| Disobedient at home | CBCL_22 | 0.702 | | 0.516 | 0.649 |
| Disobedient at school | CBCL_23 | 0.585 | | 0.506 | 0.572 |
| Doesn’t get along with peers | CBCL_25 | 0.669 | | 0.249 | 0.878 |
| No guilt | CBCL_26 | 0.601 | | 0.402 | 0.691 |
| Jealous | CBCL_27 | 0.666 | | 0.186 | 0.928 |
| Breaks rules at home | CBCL_28 | 0.695 | | 0.530 | 0.632 |

| | | | | |
|---|---|---|---|---|
| Fights | CBCL_37 | 0.609 | 0.456 | 0.641 |
| Bad friends | CBCL_39 | 0.522 | 0.416 | 0.612 |
| Lies or cheats | CBCL_43 | 0.587 | 0.446 | 0.634 |
| Attacks | CBCL_57 | 0.592 | 0.543 | 0.543 |
| Runs away | CBCL_67 | 0.471 | 0.324 | 0.679 |
| Screams | CBCL_68 | 0.694 | 0.221 | 0.908 |
| Sets fires | CBCL_72 | 0.408 | 0.225 | 0.765 |
| Shows off | CBCL_74 | 0.561 | 0.276 | 0.805 |
| Steals from home | CBCL_81 | 0.467 | 0.587 | 0.388 |
| Steals outside home | CBCL_82 | 0.414 | 0.617 | 0.310 |
| Stubborn | CBCL_86 | 0.774 | 0.151 | 0.963 |
| Suspicious | CBCL_89 | 0.638 | -0.020 | 0.999 |
| Swears | CBCL_90 | 0.551 | 0.307 | 0.763 |
| Teases | CBCL_94 | 0.601 | 0.351 | 0.746 |
| Temper | CBCL_95 | 0.741 | 0.239 | 0.906 |

| | | | | | |
|---|---|---|---|---|---|
| Thinks about sex | CBCL_96 | 0.497 | 0.163 | | 0.903 |
| Threatens | CBCL_97 | 0.625 | 0.523 | | 0.588 |
| Truants | CBCL_101 | 0.458 | 0.142 | | 0.912 |
| Vandalism | CBCL_106 | 0.549 | 0.515 | | 0.532 |
| Acts too young | CBCL_1 | 0.534 | | 0.282 | 0.782 |
| Fails to finish things | CBCL_4 | 0.686 | | 0.400 | 0.746 |
| Can’t concentrate | CBCL_8 | 0.664 | | 0.620 | 0.534 |
| Mind off | CBCL_9 | 0.680 | | 0.181 | 0.934 |
| Can’t sit still | CBCL_10 | 0.631 | | 0.352 | 0.763 |
| Confused | CBCL_13 | 0.614 | | 0.361 | 0.743 |
| Daydreams | CBCL_17 | 0.569 | | 0.501 | 0.563 |
| Gets hurt a lot | CBCL_36 | 0.589 | | 0.163 | 0.929 |
| Gets teased | CBCL_38 | 0.638 | | -0.011 | 0.999 |
| Impulsive | CBCL_41 | 0.792 | | 0.201 | 0.939 |
| Twitches | CBCL_46 | 0.557 | | 0.152 | 0.931 |

| | | | | |
|---|---|---|---|---|
| Not liked by peers | CBCL_48 | 0.715 | -0.056 | 0.994 |
| Picks skin | CBCL_58 | 0.517 | 0.125 | 0.946 |
| Poor school work | CBCL_61 | 0.573 | 0.320 | 0.762 |
| Clumsy | CBCL_62 | 0.598 | 0.320 | 0.777 |
| Prefers younger kids | CBCL_64 | 0.399 | 0.124 | 0.912 |
| Repeats acts | CBCL_66 | 0.605 | 0.208 | 0.895 |
| Sex problems | CBCL_73 | 0.478 | -0.242 | 0.796 |
| Inattentive | CBCL_78 | 0.665 | 0.599 | 0.552 |
| Speech problem | CBCL_79 | 0.305 | 0.410 | 0.356 |
| Stares | CBCL_80 | 0.543 | 0.451 | 0.592 |
| Strange behavior | CBCL_84 | 0.655 | 0.139 | 0.957 |
| Strange ideas | CBCL_85 | 0.639 | 0.020 | 0.999 |
| Talks too much | CBCL_93 | 0.600 | 0.002 | 0.999 |
| Loud | CBCL_104 | 0.696 | 0.001 | 0.999 |

| | | | |
|---|---|---|---|
| Bowel movements outside toilet | CBCL_6 | 0.369 | 1.0 |
| Doesn’t eat well | CBCL_24 | 0.431 | 1.0 |
| Hears things | CBCL_40 | 0.542 | 1.0 |
| Bites fingernails | CBCL_44 | 0.365 | 1.0 |
| Overweight | CBCL_55 | 0.291 | 1.0 |
| Skin problems | CBCL_56E | 0.300 | 1.0 |
| Plays with sex parts in public | CBCL_59 | 0.479 | 1.0 |
| Sex parts | CBCL_60 | 0.516 | 1.0 |
| Prefers older | CBCL_63 | 0.440 | 1.0 |
| Sleep more than most kids | CBCL_77 | 0.346 | 1.0 |
| Stores up | CBCL_83 | 0.533 | 1.0 |
| Sleep walks | CBCL_92 | 0.365 | 1.0 |
| Thumb sucking | CBCL_98 | 0.277 | 1.0 |
| Wets self during day | CBCL_107 | 0.429 | 1.0 |

| | | | | | | |
|---|---|---|---|---|---|---|
| Wets the bed | CBCL_108 | 0.332 | | | | 1.0 |
| Wishes to be opposite sex | CBCL_110 | 0.416 | | | | 1.0 |
| Index | | | | | | |
| PUC | | 0.739 | | | | |
| ECV SS | | 0.720 | 0.352 | 0.306 | 0.203 | |
| ECV SG | | 0.720 | 0.122 | 0.108 | 0.050 | |
| ECV GS | | 0.720 | 0.648 | 0.694 | 0.797 | |
| Omega | | 0.962 | 0.911 | 0.925 | 0.888 | |
| OmegaH | | 0.866 | 0.277 | 0.240 | 0.100 | |
| H | | 0.982 | 0.883 | 0.871 | 0.754 | |
| FD | | 0.982 | 0.935 | 0.936 | 0.910 | |

Note: IECV, item explained common variance (≥ 0.85 yield unidimensional item sets that reflect the content of the general factor); ECV, explained common variance; SS, proportion of common variance of the items in each factor which is due to that factor; SG, ECV proportion of common variance of the items in each specific factor which is due to the specific factor; GS, proportion of common variance of the items in each specific factor which is due to the general factor; PUC, percent of uncontaminated correlations; OmegaH, omega-hierarchical; H, index of construct replicability (> 0.8 suggests a well-defined latent variable); FD, factor determinacy (> 0.9 indicate that the factor score can be used)

Table S12 - CBCL bifactor model according to Clark et al., using four specific factors (EFA-generated)

| Content | Item | Factors and factor loadings | | | | | IECV |
|---|---|---|---|---|---|---|---|
| | | P-factor | Internalizing | Externalizing | Somatic | Attention | |
| Little they enjoy | CBCL_5 | 0.602 | 0.256 | | | | 0.847 |
| Lonely | CBCL_12 | 0.577 | 0.246 | | | | 0.846 |
| Cries | CBCL_14 | 0.564 | 0.130 | | | | 0.950 |
| Fears | CBCL_29 | 0.387 | 0.271 | | | | 0.671 |
| Fears school | CBCL_30 | 0.500 | 0.335 | | | | 0.690 |
| Fears do bad | CBCL_31 | 0.492 | 0.292 | | | | 0.740 |
| Perfect | CBCL_32 | 0.315 | 0.420 | | | | 0.360 |
| Unloved | CBCL_33 | 0.656 | 0.181 | | | | 0.929 |
| Feels persecuted | CBCL_34 | 0.666 | 0.122 | | | | 0.968 |
| Worthless | CBCL_35 | 0.651 | 0.369 | | | | 0.757 |
| Prefers alone | CBCL_42 | 0.471 | 0.428 | | | | 0.548 |

| | | | | | |
|---|---|---|---|---|---|
| Nervous | CBCL_45 | 0.670 | 0.342 | | 0.793 |
| Fearful | CBCL_50 | 0.545 | 0.498 | | 0.545 |
| Feels too guilty | CBCL_52 | 0.531 | 0.444 | | 0.589 |
| Won’t talk | CBCL_65 | 0.521 | 0.292 | | 0.761 |
| Self-conscious | CBCL_71 | 0.536 | 0.427 | | 0.612 |
| Shy | CBCL_75 | 0.309 | 0.494 | | 0.281 |
| Sulks | CBCL_88 | 0.679 | 0.162 | | 0.946 |
| Talks about suicide | CBCL_91 | 0.576 | 0.114 | | 0.962 |
| Sad | CBCL_103 | 0.630 | 0.437 | | 0.675 |
| Withdrawn | CBCL_111 | 0.502 | 0.539 | | 0.465 |
| Worries | CBCL_112 | 0.508 | 0.492 | | 0.516 |
| Argues | CBCL_3 | 0.688 | | 0.292 | 0.847 |
| Brags | CBCL_7 | 0.465 | | 0.327 | 0.669 |
| Cruel to animals | CBCL_15 | 0.460 | | 0.404 | 0.565 |
| Mean | CBCL_16 | 0.556 | | 0.585 | 0.475 |

| | | | | |
|---|---|---|---|---|
| Harms self | CBCL_18 | 0.485 | 0.201 | 0.853 |
| Demands a lot of attention | CBCL_19 | 0.713 | 0.181 | 0.939 |
| Destroys own things | CBCL_20 | 0.572 | 0.523 | 0.545 |
| Destroys other | CBCL_21 | 0.591 | 0.626 | 0.471 |
| Disobedient at home | CBCL_22 | 0.677 | 0.544 | 0.608 |
| Disobedient at school | CBCL_23 | 0.560 | 0.527 | 0.530 |
| Doesn’t get along with peers | CBCL_25 | 0.640 | 0.324 | 0.796 |
| No guilt | CBCL_26 | 0.579 | 0.430 | 0.643 |
| Jealous | CBCL_27 | 0.648 | 0.230 | 0.888 |
| Breaks rules at home | CBCL_28 | 0.670 | 0.556 | 0.592 |
| Fights | CBCL_37 | 0.579 | 0.504 | 0.569 |
| Bad friends | CBCL_39 | 0.501 | 0.437 | 0.568 |
| Lies or cheats | CBCL_43 | 0.567 | 0.467 | 0.596 |
| Not liked by peers | CBCL_48 | 0.663 | 0.183 | 0.929 |

| | | | | |
|---|---|---|---|---|
| Attacks | CBCL_57 | 0.562 | 0.578 | 0.486 |
| Plays with sex parts in public | CBCL_59 | 0.415 | 0.250 | 0.734 |
| Sex parts | CBCL_60 | 0.466 | 0.198 | 0.847 |
| Runs away | CBCL_67 | 0.449 | 0.359 | 0.610 |
| Screams | CBCL_68 | 0.673 | 0.269 | 0.861 |
| Sets fires | CBCL_72 | 0.392 | 0.256 | 0.701 |
| Sex problems | CBCL_73 | 0.394 | 0.244 | 0.722 |
| Shows off | CBCL_74 | 0.541 | 0.308 | 0.755 |
| Steals from home | CBCL_81 | 0.445 | 0.593 | 0.360 |
| Steals outside home | CBCL_82 | 0.389 | 0.624 | 0.280 |
| Stubborn | CBCL_86 | 0.754 | 0.207 | 0.930 |
| Mood changes | CBCL_87 | 0.785 | 0.037 | 0.998 |
| Suspicious | CBCL_89 | 0.626 | 0.033 | 0.997 |
| Swears | CBCL_90 | 0.530 | 0.346 | 0.701 |

| | | | | | |
|---|---|---|---|---|---|
| Teases | CBCL_94 | 0.578 | 0.392 | | 0.685 |
| Temper | CBCL_95 | 0.719 | 0.292 | | 0.858 |
| Thinks about sex | CBCL_96 | 0.468 | 0.249 | | 0.779 |
| Threatens | CBCL_97 | 0.595 | 0.559 | | 0.531 |
| Truants | CBCL_101 | 0.449 | 0.165 | | 0.881 |
| Vandalism | CBCL_106 | 0.521 | 0.544 | | 0.478 |
| Hears things | CBCL_40 | 0.525 | | 0.242 | 0.825 |
| Nightmares | CBCL_47 | 0.520 | | 0.183 | 0.890 |
| Constipate | CBCL_49 | 0.367 | | 0.236 | 0.707 |
| Dizzy | CBCL_51 | 0.450 | | 0.547 | 0.404 |
| Tired | CBCL_54 | 0.600 | | 0.265 | 0.837 |
| Aches | CBCL_56A | 0.459 | | 0.481 | 0.477 |
| Headaches | CBCL_56B | 0.346 | | 0.653 | 0.219 |
| Nausea | CBCL_56C | 0.429 | | 0.800 | 0.223 |
| Eye problems | CBCL_56D | 0.305 | | 0.254 | 0.590 |

| | | | | | |
|---|---|---|---|---|---|
| Skin problems | CBCL_56E | 0.325 | 0.186 | | 0.753 |
| Stomach | CBCL_56F | 0.424 | 0.684 | | 0.278 |
| Vomit | CBCL_56G | 0.389 | 0.674 | | 0.250 |
| Other physical problem | CBCL_56H | 0.339 | 0.163 | | 0.820 |
| Sees things | CBCL_70 | 0.523 | 0.243 | | 0.822 |
| Acts too young | CBCL_1 | 0.531 | | 0.270 | 0.795 |
| Fails to finish things | CBCL_4 | 0.678 | | 0.419 | 0.724 |
| Can’t concentrate | CBCL_8 | 0.654 | | 0.643 | 0.508 |
| Mind off | CBCL_9 | 0.677 | | 0.177 | 0.936 |
| Can’t sit still | CBCL_10 | 0.618 | | 0.385 | 0.720 |
| Confused | CBCL_13 | 0.616 | | 0.343 | 0.763 |
| Daydreams | CBCL_17 | 0.567 | | 0.505 | 0.558 |
| Gets hurt a lot | CBCL_36 | 0.585 | | 0.166 | 0.925 |
| Impulsive | CBCL_41 | 0.782 | | 0.225 | 0.924 |
| Twitches | CBCL_46 | 0.556 | | 0.146 | 0.935 |

| | | | | |
|---|---|---|---|---|
| Picks skin | CBCL_58 | 0.512 | 0.131 | 0.939 |
| Poor school work | CBCL_61 | 0.567 | 0.335 | 0.741 |
| Clumsy | CBCL_62 | 0.597 | 0.314 | 0.783 |
| Prefers younger kids | CBCL_64 | 0.396 | 0.118 | 0.918 |
| Repeats acts | CBCL_66 | 0.603 | 0.192 | 0.908 |
| Inattentive | CBCL_78 | 0.657 | 0.610 | 0.537 |
| Stares | CBCL_80 | 0.545 | 0.436 | 0.610 |
| Strange behavior | CBCL_84 | 0.654 | 0.117 | 0.969 |
| Strange ideas | CBCL_85 | 0.637 | 0.004 | 1.0 |
| Talks too much | CBCL_93 | 0.590 | 0.034 | 0.997 |
| Loud | CBCL_104 | 0.685 | 0.034 | 0.998 |
| Bowel movements outside toilet | CBCL_6 | 0.364 | | 1.0 |
| Too dependent | CBCL_11 | 0.558 | | 1.0 |
| Doesn’t eat well | CBCL_24 | 0.430 | | 1.0 |

| | | | |
|---|---|---|---|
| Gets teased | CBCL_38 | 0.633 | 1.0 |
| Bites fingernails | CBCL_44 | 0.363 | 1.0 |
| Overeatin | CBCL_53 | 0.469 | 1.0 |
| Overweight | CBCL_55 | 0.295 | 1.0 |
| Prefers older | CBCL_63 | 0.436 | 1.0 |
| Secretive | CBCL_69 | 0.550 | 1.0 |
| Sleeps less | CBCL_76 | 0.511 | 1.0 |
| Sleep more than most kids | CBCL_77 | 0.349 | 1.0 |
| Speech problem | CBCL_79 | 0.353 | 1.0 |
| Stores up | CBCL_83 | 0.531 | 1.0 |
| Sleep walks | CBCL_92 | 0.364 | 1.0 |
| Thumb sucking | CBCL_98 | 0.274 | 1.0 |
| Sleep problems | CBCL_100 | 0.561 | 1.0 |
| Lacks energy | CBCL_102 | 0.557 | 1.0 |

| | | | | | | | |
|---|---|---|---|---|---|---|---|
| Wets self during day | CBCL_107 | 0.424 | | | | | 1.0 |
| Wets the bed | CBCL_108 | 0.327 | | | | | 1.0 |
| Whining | CBCL_109 | 0.639 | | | | | 1.0 |
| Wishes to be opposite sex | CBCL_110 | 0.415 | | | | | 1.0 |
| Index | | | | | | | |
| PUC | | 0.815 | | | | | |
| ECV SS | | 0.709 | 0.296 | 0.328 | 0.522 | 0.217 | |
| ECV SG | | 0.709 | 0.058 | 0.126 | 0.061 | 0.045 | |
| ECV GS | | 0.709 | 0.704 | 0.672 | 0.478 | 0.783 | |
| Omega | | 0.962 | 0.873 | 0.932 | 0.792 | 0.885 | |
| OmegaH | | 0.890 | 0.233 | 0.254 | 0.383 | 0.146 | |
| H | | 0.982 | 0.775 | 0.892 | 0.843 | 0.746 | |
| FD | | 0.981 | 0.890 | 0.943 | 0.939 | 0.910 | |

Note: IECV, item explained common variance (≥ 0.85 yield unidimensional item sets that reflect the content of the general factor); ECV, explained common variance; SS, proportion of common variance of the items in each factor which is due to that factor; SG, ECV proportion of common variance of the items in each specific factor which is due to the specific factor; GS, proportion of common variance of the items in each specific factor which is due to the general factor; PUC, percent of uncontaminated correlations; OmegaH, omega-hierarchical; H, index of construct replicability (> 0.8 suggests a well-defined latent variable); FD, factor determinacy (> 0.9 indicate that the factor score can be used)

Table S13 - Impact model - global fit and factor loading

| Content | Item | Factor loading of each study<br>BHRCS (n=2,511) | HBN (n=3,199) |
|---|---|---|---|
| *Do the difficulties in emotions, concentration, behavior or being able to get on with other people:* | | | |
| Upset or distress your child? | SDQ 26A | 0.856 | 0.756 |
| Interfere with your child's everyday life in family life | SDQ 26B | 0.880 | 0.770 |
| Interfere with your child's everyday life in friendships | SDQ 26C | 0.848 | 0.678 |
| Interfere with your child's everyday life in classrom learning | SDQ 26D | 0.829 | 0.636 |
| Interfere with your child's everyday life in leisure | SDQ 26E | 0.779 | 0.754 |
| Correlation (D and F) | | 0.391 | 0.349 |
| RMSEA (90%CI) | | 0.063 (0.047-0.081) | 0.045 (0.031 - 0.061) |
| CFI | | 0.998 | 0.997 |
| TLI | | 0.995 | 0.992 |
| SRMR | | 0.010 | 0.014 |

Note: BHRCS, Brazilian High-Risk Cohort Study; HBN, Healthy Brain Network; RMSEA, Root Mean Square Error of Approximation; CFI, Comparative Fit Index; TLI, Tucker-Lewis Index; SRMR, Standardized Root Mean-square Residual.

Table S14 - Global fit indices of structural equation models of CBCL factors predicting symptom impact (SDQ impact factor), adjusted by age, gender and intelligence

| Model (Study) | RMSEA | RMSEA 90% CI | CFI | TLI | SRMR |
|---|---|---|---|---|---|
| Achenbach 2S (BHRCS) | 0.025 | 0.024 0.025 | 0.958 | 0.956 | 0.080 |
| Achenbach 2S (HBN) | 0.033 | 0.032 0.034 | 0.921 | 0.916 | 0.099 |
| Achenbach 8S (BHRCS) | 0.023 | 0.022 0.023 | 0.948 | 0.946 | 0.083 |
| Achenbach 8S (HBN) | 0.031 | 0.031 0.032 | 0.893 | 0.888 | 0.098 |
| Moore 3S (BHRCS) | 0.030 | 0.030 0.031 | 0.944 | 0.941 | 0.080 |
| Moore 3S (HBN) | 0.040 | 0.040 0.041 | 0.889 | 0.884 | 0.093 |
| Moore 4S (BHRCS) | 0.029 | 0.028 0.030 | 0.948 | 0.945 | 0.079 |
| Moore 4S (HBN) | 0.038 | 0.038 0.039 | 0.900 | 0.895 | 0.091 |
| McElroy (BHRCS) | 0.031 | 0.030 0.031 | 0.946 | 0.943 | 0.079 |
| McElroy (HBN) | 0.041 | 0.041 0.042 | 0.889 | 0.883 | 0.093 |
| Deutz GP (BHRCS) | 0.026 | 0.025 0.027 | 0.951 | 0.949 | 0.075 |
| Deutz GP (HBN) | 0.035 | 0.034 0.035 | 0.908 | 0.904 | 0.084 |

| | | | | | |
|---|---|---|---|---|---|
| Deutz and Haltigan DP (BHRCS) | 0.038 | 0.036 0.039 | 0.960 | 0.956 | 0.085 |
| Deutz and Haltigan DP (HBN) | 0.048 | 0.047 0.049 | 0.921 | 0.914 | 0.099 |
| Haltigan GP (BHRCS) | 0.024 | 0.024 0.025 | 0.949 | 0.946 | 0.080 |
| Haltigan GP (HBN) | 0.033 | 0.032 0.033 | 0.899 | 0.894 | 0.094 |
| Clark 2S (BHRCS) | 0.023 | 0.023 0.024 | 0.937 | 0.936 | 0.083 |
| Clark 2S (HBN) | 0.033 | 0.032 0.033 | 0.865 | 0.862 | 0.096 |
| Clark 3S (BHRCS) | 0.022 | 0.021 0.022 | 0.946 | 0.944 | 0.079 |
| Clark 3S (HBN) | 0.029 | 0.029 0.030 | 0.892 | 0.889 | 0.090 |
| Clark 4S (BHRCS) | 0.022 | 0.021 0.022 | 0.945 | 0.943 | 0.079 |
| Clark 4S (HBN) | 0.029 | 0.028 0.029 | 0.897 | 0.894 | 0.089 |

Note: CBCL, Child Behavior Checklist; SDQ, Strengths and Difficulties Questionnaire; BHRCS, Brazilian High-Risk Cohort Study; HBN, Healthy Brain Network; S, bifactor model with n specific (S) factors; GP, General Psychopathology; DP, Dysregulation profile; RMSEA, Root Mean Square Error of Approximation; CFI, Comparative Fit Index; TLI, Tucker-Lewis Index; SRMR, Standardized Root Mean-square Residual.

Table S15 - Results of structural equation models of latent symptom impact (SDQ) predicted by latent factors of each CBCL model, adjusted for observed variables (age, gender and IQ) for BHRCS (n=2,241) and HBN (n=2,818) separately.

| Model | Predictors | BHRCS | | | HBN | | |
|---|---|---|---|---|---|---|---|
| | | β | SE | P-value | β | SE | P-value |
| Achenbach 2S | P-factor | 0.640 | 0.017 | < 0.001 | 0.681 | 0.015 | < 0.001 |
| | Internalizing | 0.201 | 0.024 | < 0.001 | 0.002 | 0.024 | 0.921 |
| | Externalizing | 0.212 | 0.023 | < 0.001 | 0.321 | 0.019 | < 0.001 |
| | Age | 0.084 | 0.024 | 0.001 | 0.031 | 0.022 | 0.150 |
| | Gender (Stdy) | -0.186 | 0.048 | < 0.001 | -0.220 | 0.045 | < 0.001 |
| | IQ | -0.114 | 0.023 | < 0.001 | -0.087 | 0.022 | < 0.001 |
| Achenbach 8S | P-factor | 0.702 | 0.015 | < 0.001 | 0.738 | 0.014 | < 0.001 |
| | Anxious-depressed | -0.010 | 0.027 | 0.714 | 0.089 | 0.020 | < 0.001 |
| | Withdraw-depressed | 0.109 | 0.025 | < 0.001 | 0.097 | 0.022 | < 0.001 |
| | Somatic | 0.056 | 0.024 | 0.020 | -0.039 | 0.022 | 0.075 |

| | | | | | | | |
|---|---|---|---|---|---|---|---|
| | Rule breaking | 0.023 | 0.031 | 0.458 | -0.013 | 0.026 | 0.619 |
| | Aggressive | 0.015 | 0.025 | 0.554 | 0.105 | 0.022 | < 0.001 |
| | Social problems | 0.137 | 0.033 | < 0.001 | 0.200 | 0.023 | < 0.001 |
| | Thought problems | 0.032 | 0.033 | 0.330 | 0.009 | 0.026 | 0.737 |
| | Attention problems | 0.203 | 0.025 | < 0.001 | 0.218 | 0.021 | < 0.001 |
| | Age | 0.084 | 0.024 | 0.001 | 0.031 | 0.022 | 0.157 |
| | Gender (Stdy) | -0.187 | 0.048 | < 0.001 | -0.224 | 0.045 | < 0.001 |
| | IQ | -0.115 | 0.023 | < 0.001 | -0.092 | 0.022 | < 0.001 |
| Moore 3S | P-factor | 0.684 | 0.015 | < 0.001 | 0.718 | 0.015 | < 0.001 |
| | Internalizing | 0.162 | 0.021 | < 0.001 | 0.114 | 0.021 | < 0.001 |
| | Externalizing | 0.098 | 0.023 | < 0.001 | 0.194 | 0.021 | < 0.001 |
| | Attention | 0.225 | 0.027 | < 0.001 | 0.209 | 0.023 | < 0.001 |
| | Age | 0.084 | 0.024 | 0.001 | 0.03 | 0.022 | 0.169 |
| | Gender (Stdy) | -0.187 | 0.048 | < 0.001 | -0.226 | 0.045 | < 0.001 |

| | | | | | | | |
|---|---|---|---|---|---|---|---|
| | IQ | -0.115 | 0.023 | < 0.001 | -0.092 | 0.022 | < 0.001 |
| Moore 4S | P-factor | 0.684 | 0.015 | < 0.001 | 0.701 | 0.015 | < 0.001 |
| | Internalizing | 0.147 | 0.023 | < 0.001 | 0.177 | 0.021 | < 0.001 |
| | Somatic | 0.075 | 0.024 | 0.002 | -0.023 | 0.023 | 0.313 |
| | Externalizing | 0.106 | 0.023 | < 0.001 | 0.215 | 0.02 | < 0.001 |
| | Attention | 0.225 | 0.027 | < 0.001 | 0.228 | 0.022 | < 0.001 |
| | Age | 0.084 | 0.024 | 0.001 | 0.03 | 0.022 | 0.169 |
| | Gender (Stdy) | -0.187 | 0.048 | < 0.001 | -0.226 | 0.045 | < 0.001 |
| | IQ | -0.115 | 0.023 | < 0.001 | -0.092 | 0.022 | < 0.001 |
| McElroy | P-factor | 0.661 | 0.017 | < 0.001 | 0.719 | 0.015 | < 0.001 |
| | Internalizing | 0.196 | 0.022 | < 0.001 | 0.126 | 0.021 | < 0.001 |
| | Externalizing | 0.143 | 0.025 | < 0.001 | 0.188 | 0.021 | < 0.001 |
| | Attention | 0.280 | 0.034 | < 0.001 | 0.195 | 0.024 | < 0.001 |
| | Age | 0.084 | 0.024 | 0.001 | 0.030 | 0.022 | 0.166 |

| | | | | | | | |
|---|---|---|---|---|---|---|---|
| | Gender (Stdy) | -0.186 | 0.048 | < 0.001 | -0.225 | 0.045 | < 0.001 |
| | IQ | -0.115 | 0.023 | < 0.001 | -0.091 | 0.022 | < 0.001 |
| Deutz GP | P-factor | 0.655 | 0.016 | < 0.001 | 0.686 | 0.015 | < 0.001 |
| | Internalizing | 0.158 | 0.023 | < 0.001 | 0.059 | 0.022 | 0.008 |
| | Externalizing | 0.205 | 0.022 | < 0.001 | 0.292 | 0.02 | < 0.001 |
| | Age | 0.084 | 0.024 | 0.001 | 0.032 | 0.022 | 0.146 |
| | Gender (Stdy) | -0.186 | 0.048 | < 0.001 | -0.22 | 0.045 | < 0.001 |
| | IQ | -0.114 | 0.023 | < 0.001 | -0.087 | 0.022 | < 0.001 |
| Deutz-Haltigan DP | P-factor | 0.671 | 0.016 | < 0.001 | 0.712 | 0.015 | < 0.001 |
| | Anxious-Depressed | 0.136 | 0.027 | < 0.001 | 0.171 | 0.021 | < 0.001 |
| | Aggressive behavior | 0.071 | 0.027 | 0.009 | 0.181 | 0.023 | < 0.001 |
| | Attention problems | 0.281 | 0.031 | < 0.001 | 0.239 | 0.022 | < 0.001 |
| | Age | 0.085 | 0.024 | < 0.001 | 0.03 | 0.022 | 0.171 |
| | Gender (Stdy) | -0.187 | 0.048 | < 0.001 | -0.226 | 0.045 | < 0.001 |

| | | | | | | | |
|---|---|---|---|---|---|---|---|
| | IQ | -0.115 | 0.023 | < 0.001 | -0.091 | 0.022 | < 0.001 |
| Haltigan GP | P-factor | 0.668 | 0.016 | < 0.001 | 0.713 | 0.015 | < 0.001 |
| | Internalizing | 0.162 | 0.022 | < 0.001 | 0.086 | 0.021 | < 0.001 |
| | Externalizing | 0.138 | 0.024 | < 0.001 | 0.221 | 0.020 | < 0.001 |
| | Thought | 0.064 | 0.039 | 0.102 | -0.013 | 0.028 | 0.644 |
| | Attention | 0.269 | 0.032 | < 0.001 | 0.195 | 0.023 | < 0.001 |
| | Age | 0.084 | 0.024 | 0.001 | 0.03 | 0.022 | 0.163 |
| | Gender (Stdy) | -0.186 | 0.048 | < 0.001 | -0.224 | 0.045 | < 0.001 |
| | IQ | -0.115 | 0.023 | < 0.001 | -0.09 | 0.022 | < 0.001 |
| Clark 2S | P-factor | 0.711 | 0.014 | < 0.001 | 0.764 | 0.012 | < 0.001 |
| | Internalizing | 0.124 | 0.022 | < 0.001 | 0.068 | 0.019 | < 0.001 |
| | Externalizing | 0.007 | 0.023 | 0.767 | 0.065 | 0.020 | 0.001 |
| | Age | 0.084 | 0.024 | 0.001 | 0.031 | 0.022 | 0.157 |
| | Gender (Stdy) | -0.187 | 0.048 | < 0.001 | -0.107 | 0.022 | < 0.001 |

| | | | | | | | |
|---|---|---|---|---|---|---|---|
| | IQ | -0.115 | 0.023 | < 0.001 | -0.006 | 0.001 | < 0.001 |
| Clark 3S | P-factor | 0.664 | 0.016 | < 0.001 | 0.724 | 0.014 | < 0.001 |
| | Internalizing | 0.201 | 0.021 | < 0.001 | 0.126 | 0.019 | < 0.001 |
| | Externalizing | 0.117 | 0.022 | < 0.001 | 0.158 | 0.019 | < 0.001 |
| | Attention | 0.284 | 0.026 | < 0.001 | 0.189 | 0.022 | < 0.001 |
| | Age | 0.084 | 0.024 | 0.001 | 0.031 | 0.022 | 0.157 |
| | Gender (Stdy) | -0.187 | 0.048 | < 0.001 | -0.224 | 0.045 | < 0.001 |
| | IQ | -0.115 | 0.023 | < 0.001 | -0.091 | 0.022 | < 0.001 |
| Clark 4S | P-factor | 0.676 | 0.015 | < 0.001 | 0.708 | 0.014 | < 0.001 |
| | Internalizing | 0.134 | 0.024 | < 0.001 | 0.145 | 0.019 | < 0.001 |
| | Externalizing | 0.127 | 0.022 | < 0.001 | 0.21 | 0.019 | < 0.001 |
| | Somatic | 0.075 | 0.024 | 0.001 | -0.045 | 0.022 | 0.046 |
| | Attention | 0.256 | 0.025 | < 0.001 | 0.219 | 0.021 | < 0.001 |
| | Age | 0.084 | 0.024 | 0.001 | 0.031 | 0.022 | 0.157 |

| | | | | | | |
|---|---|---|---|---|---|---|
| Gender (Stdy) | -0.187 | 0.048 | < 0.001 | -0.224 | 0.045 | < 0.001 |
| IQ | -0.115 | 0.023 | < 0.001 | -0.091 | 0.022 | < 0.001 |

Note: Predictors of each CBCL model and SDQ symptom impact were modelled as latent variables. Age, gender and IQ were observed variables in the SEM. Stdy, regression coefficient standardized for the dependent variable (symptom impact), interpreted as change of symptom impact standard deviation units when gender changes male to female. All other regression coefficients (β) are interpreted as change in standard deviation units of symptom impact for a standard deviation change in the predictor variable (Stdyx standardization). CBCL, Child Behavior Checklist; SDQ, Strengths and Difficulties Questionnaire; IQ, Intelligence quotient assessed using the Wechsler Intelligence Scale for Children; SE, standard error; BHRCS, Brazilian High-Risk Cohort Study; HBN, Healthy Brain Network; S, bifactor model with n specific (S) factors; GP, General Psychopathology; DP, Dysregulation profile.

**Table** S16 - Measurement invariance testing of CBCL models across age groups

| Sample in each group | Model | Invariance | RMSEA | CFI | TLI | SRMR | Δ CFI | Δ RMSEA | Δ SRMR | Decision |
|---|---|---|---|---|---|---|---|---|---|---|
| 5 to 10 years = 3,994<br>11 to 22 = 2,326 | Achenbach 2S | Configural | 0.028 | 0.945 | 0.941 | 0.073 | | | | |
| | | Scalar | 0.027 | 0.948 | 0.946 | 0.078 | 0.003 | 0.001 | 0.005 | Invariant |
| | Achenbach 8S | Configural | 0.028 | 0.922 | 0.919 | 0.080 | | | | |
| | | Scalar | 0.027 | 0.925 | 0.923 | 0.082 | 0.003 | 0.001 | 0.002 | Invariant |
| | Moore 3S | Configural | 0.036 | 0.924 | 0.920 | 0.071 | | | | |
| | | Scalar | 0.035 | 0.928 | 0.926 | 0.072 | 0.004 | 0.001 | 0.001 | Invariant |
| | Moore 4S | Configural | 0.034 | 0.931 | 0.928 | 0.068 | | | | |

| | | | | | | | | | |
|---|---|---|---|---|---|---|---|---|---|
| | Scalar | 0.033 | 0.934 | 0.932 | 0.069 | 0.003 | 0.001 | 0.001 | Invariant |
| McElroy | Configural | 0.037 | 0.920 | 0.914 | 0.070 | | | | |
| | Scalar | 0.036 | 0.924 | 0.921 | 0.071 | 0.004 | 0.001 | 0.001 | Invariant |
| Deutz GP | Configural | 0.031 | 0.930 | 0.926 | 0.068 | | | | |
| | Scalar | 0.030 | 0.932 | 0.930 | 0.069 | 0.002 | 0.001 | 0.001 | Invariant |
| Deutz and Haltigan DP | Configural | 0.045 | 0.947 | 0.940 | 0.063 | | | | |
| | Scalar | 0.043 | 0.948 | 0.945 | 0.064 | 0.001 | 0.002 | 0.001 | Invariant |
| Haltigan GP | Configural | 0.030 | 0.925 | 0.922 | 0.075 | | | | |
| | Scalar | 0.029 | 0.928 | 0.926 | 0.076 | 0.003 | 0.001 | 0.001 | Invariant |

| | | | | | | | | | |
|---|---|---|---|---|---|---|---|---|---|
| Clark 2S | Configural | 0.029 | 0.906 | 0.903 | 0.079 | | | | |
| | Scalar | 0.028 | 0.908 | 0.906 | 0.080 | 0.002 | 0.001 | 0.001 | Invariant |
| Clark 3S | Configural | 0.025 | 0.927 | 0.924 | 0.072 | | | | |
| | Scalar | 0.025 | 0.928 | 0.927 | 0.073 | 0.001 | 0.000 | 0.001 | Invariant |
| Clark 4S | Configural | 0.026 | 0.925 | 0.923 | 0.072 | | | | |
| | Scalar | 0.025 | 0.926 | 0.925 | 0.073 | 0.001 | 0.001 | 0.001 | Invariant |

Note: The models were estimated using samples that contained information on age (HBN, BHRC, NKI and CCNP). Invariance decision is based on ΔCFI < 0.010 supplemented by ΔRMSEA < 0.015 or ΔSRMR < 0.010. CBCL, Child Behavior Checklist; RMSEA, Root Mean Square Error of Approximation; CFI, Comparative Fit Index; TLI, Tucker-Lewis Index; SRMR, Standardized Root Mean-square Residual; Δ, differences between fit index.

**Table** S17 - Measurement invariance testing of CBCL models across gender groups

| Sample in each group | Model | Invariance | RMSEA | CFI | TLI | SRMR | Δ CFI | Δ RMSEA | Δ SRMR | Decision |
|---|---|---|---|---|---|---|---|---|---|---|
| Female = 2,550 Male = 3,769 | Achenbach 2S | Configural | 0.029 | 0.939 | 0.935 | 0.076 | | | | |
| | | Scalar | 0.027 | 0.947 | 0.945 | 0.077 | 0.008 | 0.002 | 0.001 | Invariant |
| | Achenbach 8S | Configural | 0.028 | 0.921 | 0.918 | 0.079 | | | | |
| | | Scalar | 0.027 | 0.925 | 0.924 | 0.080 | 0.004 | 0.001 | 0.001 | Invariant |
| | Moore 3S | Configural | 0.036 | 0.921 | 0.916 | 0.071 | | | | |
| | | Scalar | 0.035 | 0.927 | 0.925 | 0.072 | 0.006 | 0.001 | 0.001 | Invariant |
| | Moore 4S | Configural | 0.035 | 0.928 | 0.924 | 0.069 | | | | |

| | | | | | | | | | |
|---|---|---|---|---|---|---|---|---|---|
| | Scalar | 0.033 | 0.933 | 0.931 | 0.070 | 0.005 | 0.002 | 0.001 | Invariant |
| McElroy | Configural | 0.038 | 0.917 | 0.911 | 0.071 | | | | |
| | Scalar | 0.035 | 0.924 | 0.922 | 0.071 | 0.007 | 0.003 | 0.000 | Invariant |
| Deutz GP | Configural | 0.032 | 0.927 | 0.923 | 0.068 | | | | |
| | Scalar | 0.030 | 0.932 | 0.931 | 0.069 | 0.005 | 0.002 | 0.001 | Invariant |
| Deutz and Haltigan DP | Configural | 0.044 | 0.947 | 0.941 | 0.062 | | | | |
| | Scalar | 0.042 | 0.950 | 0.947 | 0.063 | 0.003 | 0.002 | 0.001 | Invariant |
| Haltigan GP | Configural | 0.030 | 0.924 | 0.920 | 0.075 | | | | |
| | Scalar | 0.028 | 0.928 | 0.927 | 0.076 | 0.004 | 0.002 | 0.001 | Invariant |

| | | | | | | | | | |
|---|---|---|---|---|---|---|---|---|---|
| Clark 2S | Configural | 0.029 | 0.902 | 0.899 | 0.080 | | | | |
| | Scalar | 0.028 | 0.907 | 0.905 | 0.080 | 0.005 | 0.001 | 0.000 | Invariant |
| Clark 3S | Configural | 0.026 | 0.922 | 0.919 | 0.073 | | | | |
| | Scalar | 0.025 | 0.927 | 0.925 | 0.073 | 0.005 | 0.001 | 0.000 | Invariant |
| Clark 4S | Configural | 0.026 | 0.921 | 0.918 | 0.073 | | | | |
| | Scalar | 0.025 | 0.925 | 0.923 | 0.074 | 0.004 | 0.001 | 0.001 | Invariant |

Note: The models were estimated using samples that contained information on gender (HBN, BHRC, NKI and CCNP). Invariance decision is based on ΔCFI < 0.010 supplemented by ΔRMSEA < 0.015 or ΔSRMR < 0.010. CBCL, Child Behavior Checklist; RMSEA, Root Mean Square Error of Approximation; CFI, Comparative Fit Index; TLI, Tucker-Lewis Index; SRMR, Standardized Root Mean-square Residual; Δ, differences between fit index.

Table S18 - Measurement invariance testing of CBCL models across study site

| Sample in each group | Model | Invariance | RMSEA | CFI | TLI | SRMR | Δ CFI | Δ RMSEA | Δ SRMR | Decision |
|---|---|---|---|---|---|---|---|---|---|---|
| HBN 1 = 1,776<br>HBN 3 = 1,099<br>BHRCS 1 = 1,255<br>BHRCS 2 = 1,256 | Achenbach 2S | Configural | 0.027 | 0.943 | 0.939 | 0.095 | | | | |
| | | Scalar | 0.027 | 0.940 | 0.939 | 0.101 | 0.003 | 0.000 | 0.006 | Invariant |
| | Achenbach 8S | Configural | 0.025 | 0.925 | 0.922 | 0.096 | | | | |
| | | Scalar | 0.025 | 0.925 | 0.924 | 0.099 | 0.000 | 0.000 | 0.003 | Invariant |
| | Moore 3S | Configural | 0.033 | 0.926 | 0.922 | 0.084 | | | | |
| | | Scalar | 0.033 | 0.925 | 0.923 | 0.088 | 0.001 | 0.000 | 0.004 | Invariant |
| | Moore 4S | Configural | 0.032 | 0.931 | 0.928 | 0.083 | | | | |

| | | | | | | | | | |
|---|---|---|---|---|---|---|---|---|---|
| | Scalar | 0.032 | 0.929 | 0.928 | 0.087 | 0.002 | 0.000 | 0.004 | Invariant |
| McElroy | Configural | 0.035 | 0.924 | 0.919 | 0.082 | | | | |
| | Scalar | 0.034 | 0.923 | 0.921 | 0.087 | 0.001 | 0.001 | 0.005 | Invariant |
| Deutz GP | Configural | 0.028 | 0.935 | 0.931 | 0.081 | | | | |
| | Scalar | 0.029 | 0.928 | 0.927 | 0.084 | 0.007 | 0.001 | 0.003 | Invariant |
| Deutz and Haltigan DP | Configural | 0.043 | 0.947 | 0.940 | 0.073 | | | | |
| | Scalar | 0.045 | 0.940 | 0.938 | 0.078 | 0.007 | 0.002 | 0.005 | Invariant |
| Haltigan GP | Configural | 0.026 | 0.930 | 0.926 | 0.089 | | | | |
| | Scalar | 0.026 | 0.927 | 0.926 | 0.093 | 0.003 | 0.000 | 0.004 | Invariant |

| | | | | | | | | | |
|---|---|---|---|---|---|---|---|---|---|
| Clark 2S | Configural | 0.026 | 0.907 | 0.905 | 0.095 | | | | |
| | Scalar | 0.026 | 0.906 | 0.905 | 0.097 | 0.001 | 0.000 | 0.002 | Invariant |
| Clark 3S | Configural | 0.023 | 0.927 | 0.925 | 0.088 | | | | |
| | Scalar | 0.023 | 0.925 | 0.925 | 0.091 | 0.002 | 0.000 | 0.003 | Invariant |
| Clark 4S | Configural | 0.023 | 0.924 | 0.922 | 0.089 | | | | |
| | Scalar | 0.023 | 0.923 | 0.923 | 0.091 | 0.001 | 0.000 | 0.002 | Invariant |

Note: We used only study site that would have sufficient size to test measurement invariance (two in BHRCS and two in HBN). Invariance decision is based on ΔCFI < 0.010 supplemented by ΔRMSEA < 0.015 or ΔSRMR < 0.010. CBCL, Child Behavior Checklist; RMSEA, Root Mean Square Error of Approximation; CFI, Comparative Fit Index; TLI, Tucker-Lewis Index; SRMR, Standardized Root Mean-square Residual; Δ, differences between fit index.

Table S19 - Measurement invariance testing of CBCL models across race/ethnicity groups

| Sample in each group | Model | Invariance | RMSEA | CFI | TLI | SRMR | Δ CFI | Δ RMSEA | Δ SRMR | Decision |
|---|---|---|---|---|---|---|---|---|---|---|
| White = 3,349<br>Non-white = 2,629 | Achenbach 2S | Configural | 0.029 | 0.942 | 0.938 | 0.078 | | | | |
| | | Scalar | 0.027 | 0.948 | 0.946 | 0.080 | 0.006 | 0.002 | 0.002 | Invariant |
| | Achenbach 8S | Configural | 0.029 | 0.919 | 0.916 | 0.082 | | | | |
| | | Scalar | 0.028 | 0.924 | 0.922 | 0.083 | 0.005 | 0.001 | 0.001 | Invariant |
| | Moore 3S | Configural | 0.037 | 0.921 | 0.916 | 0.073 | | | | |
| | | Scalar | 0.035 | 0.928 | 0.926 | 0.073 | 0.007 | 0.002 | 0.000 | Invariant |
| | Moore 4S | Configural | 0.036 | 0.927 | 0.923 | 0.071 | | | | |

| | | | | | | | | | |
|---|---|---|---|---|---|---|---|---|---|
| | Scalar | 0.034 | 0.933 | 0.931 | 0.071 | 0.006 | 0.002 | 0.000 | Invariant |
| McElroy | Configural | 0.039 | 0.916 | 0.910 | 0.072 | | | | |
| | Scalar | 0.036 | 0.924 | 0.922 | 0.073 | 0.008 | 0.003 | 0.001 | Invariant |
| Deutz GP | Configural | 0.032 | 0.927 | 0.923 | 0.069 | | | | |
| | Scalar | 0.030 | 0.934 | 0.932 | 0.070 | 0.007 | 0.002 | 0.001 | Invariant |
| Deutz and Haltigan DP | Configural | 0.046 | 0.945 | 0.938 | 0.064 | | | | |
| | Scalar | 0.042 | 0.950 | 0.946 | 0.065 | 0.005 | 0.004 | 0.001 | Invariant |
| Haltigan GP | Configural | 0.030 | 0.922 | 0.918 | 0.077 | | | | |
| | Scalar | 0.029 | 0.928 | 0.926 | 0.078 | 0.006 | 0.001 | 0.001 | Invariant |

| | | | | | | | | | |
|---|---|---|---|---|---|---|---|---|---|
| Clark 2S | Configural | 0.030 | 0.901 | 0.898 | 0.081 | | | | |
| | Scalar | 0.029 | 0.906 | 0.905 | 0.081 | 0.005 | 0.001 | 0.000 | Invariant |
| Clark 3S | Configural | 0.026 | 0.922 | 0.919 | 0.074 | | | | |
| | Scalar | 0.025 | 0.927 | 0.926 | 0.074 | 0.005 | 0.001 | 0.000 | Invariant |
| Clark 4S | Configural | 0.027 | 0.919 | 0.916 | 0.075 | | | | |
| | Scalar | 0.026 | 0.924 | 0.923 | 0.075 | 0.005 | 0.001 | 0.000 | Invariant |

Note: The models were estimated using samples that contained information on race and ethnicity (HBN, BHRC and NKI). Invariance decision is based on ΔCFI < 0.010 supplemented by ΔRMSEA < 0.015 or ΔSRMR < 0.010. CBCL, Child Behavior Checklist; RMSEA, Root Mean Square Error of Approximation; CFI, Comparative Fit Index; TLI, Tucker-Lewis Index; SRMR, Standardized Root Mean-square Residual; Δ, differences between fit index.

Table S20 - Measurement invariance testing of CBCL models across educational level groups

| Sample in each group | Model | Invariance | RMSEA | CFI | TLI | SRMR | Δ CFI | Δ RMSEA | Δ SRMR | Decision |
|---|---|---|---|---|---|---|---|---|---|---|
| Primary school = 2,892<br>Secondary school = 692 | Achenbach 2S | Configural | 0.025 | 0.939 | 0.935 | 0.088 | | | | |
| | | Scalar | 0.024 | 0.944 | 0.943 | 0.092 | 0.005 | 0.001 | 0.004 | Invariant |
| | Achenbach 8S | Configural | 0.024 | 0.924 | 0.921 | 0.092 | | | | |
| | | Scalar | 0.023 | 0.928 | 0.927 | 0.093 | 0.004 | 0.001 | 0.001 | Invariant |
| | Moore 3S | Configural | 0.032 | 0.924 | 0.919 | 0.082 | | | | |
| | | Scalar | 0.030 | 0.930 | 0.928 | 0.084 | 0.006 | 0.002 | 0.002 | Invariant |

| | | | | | | | | | |
|---|---|---|---|---|---|---|---|---|---|
| Moore 4S | Configural | 0.031 | 0.929 | 0.925 | 0.081 | | | | |
| | Scalar | 0.029 | 0.934 | 0.932 | 0.082 | 0.005 | 0.002 | 0.001 | Invariant |
| McElroy | Configural | 0.033 | 0.918 | 0.913 | 0.079 | | | | |
| | Scalar | 0.031 | 0.926 | 0.923 | 0.080 | 0.008 | 0.002 | 0.001 | Invariant |
| Deutz GP | Configural | 0.027 | 0.929 | 0.926 | 0.079 | | | | |
| | Scalar | 0.026 | 0.934 | 0.933 | 0.081 | 0.005 | 0.001 | 0.002 | Invariant |
| Deutz and Haltigan DP | Configural | 0.041 | 0.945 | 0.938 | 0.070 | | | | |
| | Scalar | 0.038 | 0.950 | 0.947 | 0.071 | 0.005 | 0.003 | 0.001 | Invariant |
| Haltigan GP | Configural | 0.025 | 0.927 | 0.924 | 0.086 | | | | |

| | | | | | | | | | |
|---|---|---|---|---|---|---|---|---|---|
| | Scalar | 0.024 | 0.932 | 0.930 | 0.087 | 0.005 | 0.001 | 0.001 | Invariant |
| Clark 2S | Configural | 0.024 | 0.910 | 0.907 | 0.092 | | | | |
| | Scalar | 0.023 | 0.914 | 0.913 | 0.093 | 0.004 | 0.001 | 0.001 | Invariant |
| Clark 3S | Configural | 0.021 | 0.927 | 0.924 | 0.086 | | | | |
| | Scalar | 0.021 | 0.931 | 0.930 | 0.087 | 0.004 | 0.000 | 0.001 | Invariant |
| Clark 4S | Configural | 0.022 | 0.923 | 0.920 | 0.088 | | | | |
| | Scalar | 0.021 | 0.927 | 0.925 | 0.089 | 0.004 | 0.001 | 0.001 | Invariant |

Note: The models were estimated using samples that contained information on educational level (HBN, BHRC and NKI). Invariance decision is based on ΔCFI < 0.010 supplemented by ΔRMSEA < 0.015 or ΔSRMR < 0.010. CBCL, Child Behavior Checklist; RMSEA, Root Mean Square Error of Approximation; CFI, Comparative Fit Index; TLI, Tucker-Lewis Index; SRMR, Standardized Root Mean-square Residual; Δ, differences between fit index.

Table S21 - Measurement invariance testing of CBCL models across IQ groups

| Sample in each group | Model | Invariance | RMSEA | CFI | TLI | SRMR | Δ CFI | Δ RMSEA | Δ SRMR | Decision |
|---|---|---|---|---|---|---|---|---|---|---|
| IQ < 90 = 1,419<br>IQ ≥ 90 and < 110 = 2,646<br>IQ ≥ 110 = 1,350 | Achenbach 2S | Configural | 0.027 | 0.947 | 0.943 | 0.085 | | | | |
| | | Scalar | 0.024 | 0.954 | 0.953 | 0.086 | 0.007 | 0.003 | 0.001 | Invariant |
| | Achenbach 8S | Configural | 0.026 | 0.930 | 0.927 | 0.087 | | | | |
| | | Scalar | 0.024 | 0.935 | 0.934 | 0.088 | 0.005 | 0.002 | 0.001 | Invariant |
| | Moore 3S | Configural | 0.035 | 0.929 | 0.925 | 0.076 | | | | |
| | | Scalar | 0.032 | 0.937 | 0.935 | 0.077 | 0.008 | 0.003 | 0.001 | Invariant |
| | Moore 4S | Configural | 0.033 | 0.936 | 0.932 | 0.074 | | | | |

| | | | | | | | | | |
|---|---|---|---|---|---|---|---|---|---|
| | Scalar | 0.031 | 0.942 | 0.940 | 0.075 | 0.006 | 0.002 | 0.001 | Invariant |
| McElroy | Configural | 0.036 | 0.923 | 0.918 | 0.075 | | | | |
| | Scalar | 0.033 | 0.933 | 0.931 | 0.077 | 0.010 | 0.003 | 0.002 | Invariant |
| Deutz GP | Configural | 0.030 | 0.934 | 0.931 | 0.073 | | | | |
| | Scalar | 0.027 | 0.941 | 0.940 | 0.075 | 0.007 | 0.003 | 0.002 | Invariant |
| Deutz and Haltigan DP | Configural | 0.044 | 0.949 | 0.943 | 0.066 | | | | |
| | Scalar | 0.040 | 0.953 | 0.951 | 0.067 | 0.004 | 0.004 | 0.001 | Invariant |
| Haltigan GP | Configural | 0.027 | 0.931 | 0.927 | 0.082 | | | | |
| | Scalar | 0.026 | 0.938 | 0.936 | 0.083 | 0.007 | 0.001 | 0.001 | Invariant |

| | | | | | | | | | |
|---|---|---|---|---|---|---|---|---|---|
| Clark 2S | Configural | 0.027 | 0.912 | 0.909 | 0.087 | | | | |
| | Scalar | 0.026 | 0.917 | 0.916 | 0.088 | 0.005 | 0.001 | 0.001 | Invariant |
| Clark 3S | Configural | 0.024 | 0.931 | 0.928 | 0.080 | | | | |
| | Scalar | 0.023 | 0.936 | 0.935 | 0.081 | 0.005 | 0.001 | 0.001 | Invariant |
| Clark 4S | Configural | 0.024 | 0.929 | 0.927 | 0.080 | | | | |
| | Scalar | 0.023 | 0.934 | 0.933 | 0.081 | 0.005 | 0.001 | 0.001 | Invariant |

Note: The models were estimated using samples that contained information on IQ (HBN, BHRC and NKI). Invariance decision is based on ΔCFI < 0.010 supplemented by ΔRMSEA < 0.015 or ΔSRMR < 0.010. CBCL, Child Behavior Checklist; RMSEA, Root Mean Square Error of Approximation; CFI, Comparative Fit Index; TLI, Tucker-Lewis Index; SRMR, Standardized Root Mean-square Residual; Δ, differences between fit index.

Table S22 - Measurement invariance testing of CBCL models across mental health condition status (presence vs. no condition)

| Sample in each group | Model | Invariance | RMSEA | CFI | TLI | SRMR | Δ CFI | Δ RMSEA | Δ SRMR | Decision |
|---|---|---|---|---|---|---|---|---|---|---|
| Yes = 3,637<br>No = 2,340 | Achenbach 2S | Configural | 0.028 | 0.931 | 0.927 | 0.083 | | | | |
| | | Scalar | 0.027 | 0.935 | 0.933 | 0.086 | 0.004 | 0.001 | 0.003 | Invariant |
| | Achenbach 8S | Configural | 0.027 | 0.900 | 0.896 | 0.087 | | | | |
| | | Scalar | 0.026 | 0.905 | 0.903 | 0.088 | 0.005 | 0.001 | 0.001 | Invariant |
| | Moore 3S | Configural | 0.035 | 0.901 | 0.896 | 0.079 | | | | |
| | | Scalar | 0.034 | 0.907 | 0.904 | 0.081 | 0.006 | 0.001 | 0.002 | Invariant |
| | Moore 4S | Configural | 0.034 | 0.909 | 0.904 | 0.078 | | | | |

| | | | | | | | | | |
|---|---|---|---|---|---|---|---|---|---|
| | Scalar | 0.033 | 0.913 | 0.911 | 0.079 | 0.004 | 0.001 | 0.001 | Invariant |
| McElroy | Configural | 0.036 | 0.901 | 0.895 | 0.077 | | | | |
| | Scalar | 0.035 | 0.906 | 0.903 | 0.079 | 0.005 | 0.001 | 0.002 | Invariant |
| Deutz GP | Configural | 0.030 | 0.918 | 0.913 | 0.074 | | | | |
| | Scalar | 0.029 | 0.919 | 0.917 | 0.076 | 0.001 | 0.001 | 0.002 | Invariant |
| Deutz and Haltigan DP | Configural | 0.043 | 0.935 | 0.927 | 0.069 | | | | |
| | Scalar | 0.043 | 0.934 | 0.930 | 0.072 | 0.001 | 0.000 | 0.003 | Invariant |
| Haltigan GP | Configural | 0.028 | 0.905 | 0.901 | 0.083 | | | | |
| | Scalar | 0.027 | 0.909 | 0.907 | 0.084 | 0.004 | 0.001 | 0.001 | Invariant |

| | | | | | | | | | |
|---|---|---|---|---|---|---|---|---|---|
| Clark 2S | Configural | 0.028 | 0.875 | 0.872 | 0.088 | | | | |
| | Scalar | 0.027 | 0.880 | 0.878 | 0.089 | 0.005 | 0.001 | 0.001 | Invariant |
| Clark 3S | Configural | 0.025 | 0.903 | 0.900 | 0.080 | | | | |
| | Scalar | 0.024 | 0.907 | 0.906 | 0.081 | 0.004 | 0.001 | 0.001 | Invariant |
| Clark 4S | Configural | 0.025 | 0.900 | 0.897 | 0.081 | | | | |
| | Scalar | 0.024 | 0.905 | 0.903 | 0.082 | 0.005 | 0.001 | 0.001 | Invariant |

Note: The models were estimated using samples that contained information on mental health condition status (HBN, BHRC and NKI). Invariance decision is based on ΔCFI < 0.010 supplemented by ΔRMSEA < 0.015 or ΔSRMR < 0.010. CBCL, Child Behavior Checklist; RMSEA, Root Mean Square Error of Approximation; CFI, Comparative Fit Index; TLI, Tucker-Lewis Index; SRMR, Standardized Root Mean-square Residual; Δ, differences between fit index.

Table S23 - Measurement invariance testing of CBCL models across study waves

| Sample in each group | Model | Invariance | RMSEA | CFI | TLI | SRMR | Δ CFI | Δ RMSEA | Δ SRMR | Decision |
|---|---|---|---|---|---|---|---|---|---|---|
| Wave 1 = 3,064<br>Wave 2 = 2,354<br>Wave 3 = 1,086 | Achenbach 2S | Configural | 0.024 | 0.951 | 0.948 | 0.078 | | | | |
| | | Scalar | 0.023 | 0.954 | 0.953 | 0.082 | 0.003 | 0.001 | 0.004 | Invariant |
| | Achenbach 8S | Configural | 0.022 | 0.940 | 0.938 | 0.082 | | | | |
| | | Scalar | 0.021 | 0.944 | 0.943 | 0.083 | 0.004 | 0.001 | 0.001 | Invariant |
| | Moore 3S | Configural | 0.030 | 0.939 | 0.936 | 0.077 | | | | |
| | | Scalar | 0.028 | 0.945 | 0.943 | 0.078 | 0.006 | 0.002 | 0.001 | Invariant |

| | | | | | | | | | |
|---|---|---|---|---|---|---|---|---|---|
| Moore 4S | Configural | 0.028 | 0.945 | 0.942 | 0.075 | | | | |
| | Scalar | 0.026 | 0.950 | 0.949 | 0.076 | 0.005 | 0.002 | 0.001 | Invariant |
| McElroy | Configural | 0.031 | 0.938 | 0.934 | 0.073 | | | | |
| | Scalar | 0.029 | 0.944 | 0.942 | 0.075 | 0.006 | 0.002 | 0.002 | Invariant |
| Deutz GP | Configural | 0.026 | 0.945 | 0.942 | 0.073 | | | | |
| | Scalar | 0.024 | 0.948 | 0.947 | 0.075 | 0.003 | 0.002 | 0.002 | Invariant |
| Deutz and Haltigan DP | Configural | 0.038 | 0.957 | 0.952 | 0.067 | | | | |
| | Scalar | 0.036 | 0.960 | 0.958 | 0.069 | 0.003 | 0.002 | 0.002 | Invariant |
| Haltigan GP | Configural | 0.024 | 0.941 | 0.938 | 0.078 | | | | |

| | | | | | | | | | |
|---|---|---|---|---|---|---|---|---|---|
| | Scalar | 0.022 | 0.945 | 0.944 | 0.080 | 0.004 | 0.002 | 0.002 | Invariant |
| Clark 2S | Configural | 0.022 | 0.924 | 0.922 | 0.085 | | | | |
| | Scalar | 0.021 | 0.930 | 0.929 | 0.085 | 0.006 | 0.001 | 0.000 | Invariant |
| Clark 3S | Configural | 0.020 | 0.935 | 0.933 | 0.079 | | | | |
| | Scalar | 0.019 | 0.941 | 0.940 | 0.080 | 0.006 | 0.001 | 0.001 | Invariant |
| Clark 4S | Configural | 0.021 | 0.933 | 0.931 | 0.080 | | | | |
| | Scalar | 0.020 | 0.938 | 0.938 | 0.081 | 0.005 | 0.001 | 0.001 | Invariant |

Note: The models were estimated using samples that contained longitudinal information (BHRC, NKI and CCNP). Invariance decision is based on ΔCFI < 0.010 supplemented by ΔRMSEA < 0.015 or ΔSRMR < 0.010. CBCL, Child Behavior Checklist; RMSEA, Root Mean Square Error of Approximation; CFI, Comparative Fit Index; TLI, Tucker-Lewis Index; SRMR, Standardized Root Mean-square Residual; Δ, differences between fit index.

Table S24 - Measurement invariance testing of CBCL models across informants (parent-CBCL, teacher-TRF and self-YSR)

| Sample in each group | Model | Invariance | RMSEA | CFI | TLI | SRMR | Δ CFI | Δ RMSEA | Δ SRMR | Decision |
|---|---|---|---|---|---|---|---|---|---|---|
| Parent (CBCL) = 3,270<br>Teacher (TRF) = 960<br>Self (YSR) = 1,154 | Achenbach 2S | Configural | 0.031 | 0.931 | 0.926 | 0.092 | | | | |
| | | Scalar | 0.032 | 0.922 | 0.920 | 0.096 | 0.009 | 0.001 | 0.004 | Invariant |
| | Achenbach 8S | Configural | 0.031 | 0.896 | 0.891 | 0.096 | | | | |
| | | Scalar | 0.031 | 0.890 | 0.888 | 0.099 | 0.006 | 0.000 | 0.003 | Invariant |
| | Moore 3S | Configural | 0.037 | 0.908 | 0.902 | 0.086 | | | | |
| | | Scalar | 0.037 | 0.900 | 0.898 | 0.090 | 0.008 | 0.000 | 0.004 | Invariant |
| | Moore 4S | Configural | 0.035 | 0.916 | 0.911 | 0.083 | | | | |

| | | | | | | | | | |
|---|---|---|---|---|---|---|---|---|---|
| | Scalar | 0.036 | 0.908 | 0.906 | 0.087 | 0.008 | 0.001 | 0.004 | Invariant |
| McElroy | Configural | 0.036 | 0.909 | 0.903 | 0.086 | | | | |
| | Scalar | 0.038 | 0.897 | 0.894 | 0.093 | 0.012 | 0.002 | 0.007 | Invariant |
| Deutz GP | Configural | 0.033 | 0.903 | 0.898 | 0.089 | | | | |
| | Scalar | 0.034 | 0.893 | 0.891 | 0.093 | 0.010 | 0.001 | 0.004 | Invariant |
| Deutz and Haltigan DP | Configural | 0.042 | 0.940 | 0.933 | 0.075 | | | | |
| | Scalar | 0.045 | 0.927 | 0.924 | 0.082 | 0.013 | 0.003 | 0.007 | Invariant |
| Haltigan GP | Configural | 0.031 | 0.912 | 0.907 | 0.091 | | | | |
| | Scalar | 0.032 | 0.900 | 0.898 | 0.096 | 0.012 | 0.001 | 0.005 | Invariant |

| | | | | | | | | | |
|---|---|---|---|---|---|---|---|---|---|
| Clark 2S | Configural | 0.033 | 0.873 | 0.869 | 0.096 | | | | |
| | Scalar | 0.034 | 0.865 | 0.864 | 0.099 | 0.008 | 0.001 | 0.003 | Invariant |
| Clark 3S | Configural | 0.029 | 0.904 | 0.900 | 0.087 | | | | |
| | Scalar | 0.030 | 0.894 | 0.892 | 0.092 | 0.010 | 0.001 | 0.005 | Invariant |
| Clark 4S | Configural | 0.029 | 0.904 | 0.900 | 0.087 | | | | |
| | Scalar | 0.030 | 0.896 | 0.894 | 0.091 | 0.008 | 0.001 | 0.004 | Invariant |

Note: The models were estimated using samples that contained information from different raters (HBN). To match to CBCL (parent-informed), we used teachers ratings assessed with the Teacher's Report Form (TRF) and self-reports using the Youth Self-Report (YSR) and. CBCL, TRF and YSR have 100 out of 120 matched items. CBCL unmatched items were excluded from all models. Invariance decision is based on ΔCFI < 0.010 supplemented by ΔRMSEA < 0.015 or ΔSRMR < 0.010. ASEBA, Achenbach System of Empirically Based Assessment; CBCL, Child Behavior Checklist; RMSEA, Root Mean Square Error of Approximation; CFI, Comparative Fit Index; TLI, Tucker-Lewis Index; SRMR, Standardized Root Mean-square Residual; Δ, differences between fit index.

Table S25 - Correlation between p-factors derived from 11 CBCL bifactor models

| Model | | Achenbach 2S | Achenbach 8S | Moore 3S | Moore 4S | McElroy | Deutz GP | Deutz DP | Haltigan GP | Clark 2S | Clark 3S |
|---|---|---|---|---|---|---|---|---|---|---|---|
| Achenbach 8S | | 0.929 | | | | | | | | | |
| | CI | [0.925; 0.932] | | | | | | | | | |
| | p-value | < 0.001 | | | | | | | | | |
| Moore 3S | | 0.938 | 0.982 | | | | | | | | |
| | CI | [0.935; 0.941] | [0.981; 0.982] | | | | | | | | |
| | p-value | < 0.001 | < 0.001 | | | | | | | | |
| Moore 4S | | 0.934 | 0.982 | 0.997 | | | | | | | |
| | CI | [0.931; 0.937] | [0.981; 0.983] | [0.997; 0.997] | | | | | | | |

| | | | | | | | |
|---|---|---|---|---|---|---|---|
| | p-value | < 0.001 | < 0.001 | < 0.001 | | | |
| McElroy | | 0.960 | 0.97 | 0.979 | 0.975 | | |
| | CI | [0.958; 0.962] | [0.969; 0.972] | [0.978; 0.98] | [0.973; 0.976] | | |
| | p-value | < 0.001 | < 0.001 | < 0.001 | < 0.001 | | |
| Deutz GP | | 0.962 | 0.949 | 0.946 | 0.945 | 0.942 | |
| | CI | [0.96; 0.963] | [0.947; 0.951] | [0.944; 0.949] | [0.942; 0.947] | [0.939; 0.945] | |
| | p-value | < 0.001 | < 0.001 | < 0.001 | < 0.001 | < 0.001 | |
| Deutz DP | | 0.932 | 0.956 | 0.967 | 0.965 | 0.989 | 0.91 |
| | CI | [0.929; 0.935] | [0.954; 0.958] | [0.966; 0.969] | [0.963; 0.966] | [0.988; 0.989] | [0.906; 0.914] |

| | | | | | | | | | |
|---|---|---|---|---|---|---|---|---|---|
| p-value | < 0.001 | < 0.001 | < 0.001 | < 0.001 | < 0.001 | < 0.001 | | | |
| Haltigan GP | 0.949 | 0.98 | 0.981 | 0.978 | 0.980 | 0.977 | 0.958 | | |
| CI | [0.946; 0.951] | [0.979; 0.981] | [0.98; 0.981] | [0.977; 0.979] | [0.979; 0.981] | [0.976; 0.978] | [0.956; 0.96] | | |
| p-value | < 0.001 | < 0.001 | < 0.001 | < 0.001 | < 0.001 | < 0.001 | < 0.001 | | |
| Clark 2S | 0.878 | 0.982 | 0.968 | 0.966 | 0.947 | 0.920 | 0.935 | 0.967 | |
| CI | [0.872; 0.883] | [0.981; 0.983] | [0.966; 0.97] | [0.964; 0.968] | [0.945; 0.95] | [0.916; 0.923] | [0.932; 0.938] | [0.966; 0.969] | |
| p-value | < 0.001 | < 0.001 | < 0.001 | < 0.001 | < 0.001 | < 0.001 | < 0.001 | < 0.001 | |
| Clark 3S | 0.911 | 0.984 | 0.978 | 0.975 | 0.960 | 0.943 | 0.948 | 0.975 | 0.984 |
| CI | [0.907; 0.915] | [0.983; 0.985] | [0.977; 0.979] | [0.974; 0.976] | [0.958; 0.962] | [0.941; 0.946] | [0.946; 0.951] | [0.974; 0.976] | [0.983; 0.984] |

| | | | | | | | | | | | |
|---|---|---|---|---|---|---|---|---|---|---|---|
| | p-value | < 0.001 | < 0.001 | < 0.001 | < 0.001 | < 0.001 | < 0.001 | < 0.001 | < 0.001 | < 0.001 | |
| Clark 4S | | 0.924 | 0.987 | 0.974 | 0.977 | 0.957 | 0.958 | 0.938 | 0.978 | 0.978 | 0.991 |
| | CI | [0.921; 0.928] | [0.986; 0.988] | [0.973; 0.975] | [0.976; 0.978] | [0.955; 0.959] | [0.956; 0.96] | [0.935; 0.941] | [0.977; 0.979] | [0.977; 0.979] | [0.991; 0.991] |
| | p-value | < 0.001 | < 0.001 | < 0.001 | < 0.001 | < 0.001 | < 0.001 | < 0.001 | < 0.001 | < 0.001 | < 0.001 |

Note: nS, number of specific factors in the model; GP, General psychopathology model; DP, dysregulation profile model; CI, 95% confidence interval.

Table S26 - Correlation between specific factors derived from 11 CBCL bifactor models

| | Int. (Achenbach 2S) | Anxiety-dep. (Achenbach 8S) | Withdrawn-dep. (Achenbach 8S) | Som. (Achenbach 8S) | Int. (Moore 3S) | Int. (Moore 4S) | Som. (Moore 4S) | Int. (McElroy) | Int. (Deutz GP) | Anxiety-dep. (Deutz DP) | Int. (Haltigan GP) | Int. (Clark 2S) | Int. (Clark 3S) | Int. (Clark 4S) | Som. (Clark 4S) | Ext. (Achenbach 2S) | Rule-Breaking (Achenbach 8S) | Aggressive (Achenbach 8S) | Ext. (Moore 3S) | Ext. (Moore 4S) | Ext. (McElroy) | Ext. (Deutz GP) | Aggressive (Deutz DP) | Ext. (Haltigan GP) | Ext. (Clark 2S) | Ext. (Clark 3S) | Ext. (Clark 4S) | Att. (Achenbach 8S) | Att. (Moore 3S) | Att. (Moore 4S) | Att. (McElroy) | Att. (Deutz DP) | Att. (Haltigan GP) | Att. (Clark 3S) | Att. (Clark 4S) | Social (Achenbach 8S) | Tho. (Achenbach 8S) | Tho. (Haltigan GP) |
|---|---|---|---|---|---|---|---|---|---|---|---|---|---|---|---|---|---|---|---|---|---|---|---|---|---|---|---|---|---|---|---|---|---|---|---|---|---|---|
| Int. (Achenbach 2S) | | | | | | | | | | | | | | | | | | | | | | | | | | | | | | | | | | | | | | |
| Anxiety-dep. (Achenbach 8S) | 0.463 | | | | | | | | | | | | | | | | | | | | | | | | | | | | | | | | | | | | | |
| Withdrawn-dep. (Achenbach 8S) | 0.466 | 0.186 | | | | | | | | | | | | | | | | | | | | | | | | | | | | | | | | | | | | |
| Som. (Achenbach 8S) | 0.765 | 0.18 | 0.089 | | | | | | | | | | | | | | | | | | | | | | | | | | | | | | | | | | | |
| Int. (Moore 3S) | 0.933 | 0.544 | 0.596 | 0.715 | | | | | | | | | | | | | | | | | | | | | | | | | | | | | | | | | | |
| Int. (Moore 4S) | 0.625 | 0.592 | 0.847 | 0.136 | 0.76 | | | | | | | | | | | | | | | | | | | | | | | | | | | | | | | | | |
| Som. (Moore 4S) | 0.752 | 0.156 | 0.091 | 0.978 | 0.72 | 0.131 | | | | | | | | | | | | | | | | | | | | | | | | | | | | | | | | |
| Int. (McElroy) | 0.953 | 0.624 | 0.504 | 0.71 | 0.961 | 0.718 | 0.689 | | | | | | | | | | | | | | | | | | | | | | | | | | | | | | | |
| Int. (Deutz GP) | 0.952 | 0.422 | 0.449 | 0.83 | 0.935 | 0.575 | 0.822 | 0.921 | | | | | | | | | | | | | | | | | | | | | | | | | | | | | | |
| Anxiety-dep. (Deutz DP) | 0.569 | 0.96 | 0.259 | 0.209 | 0.621 | 0.669 | 0.185 | 0.722 | 0.489 | | | | | | | | | | | | | | | | | | | | | | | | | | | | | |
| Int. (Haltigan GP) | 0.942 | 0.575 | 0.478 | 0.779 | 0.966 | 0.666 | 0.763 | 0.977 | 0.967 | 0.644 | | | | | | | | | | | | | | | | | | | | | | | | | | | | |

| | | | | | | | | | | | | | | | | | | | | | |
|---|---|---|---|---|---|---|---|---|---|---|---|---|---|---|---|---|---|---|---|---|---|
| Int. (Clark 2S) | 0.814 | 0.597 | 0.525 | 0.687 | 0.923 | 0.691 | 0.674 | 0.908 | 0.866 | 0.635 | 0.932 | | | | | | | | | | |
| Int. (Clark 3S) | 0.867 | 0.609 | 0.548 | 0.688 | 0.95 | 0.727 | 0.669 | 0.942 | 0.89 | 0.667 | 0.951 | 0.956 | | | | | | | | | |
| Int. (Clark 4S) | 0.554 | 0.805 | 0.626 | 0.143 | 0.703 | 0.882 | 0.141 | 0.699 | 0.54 | 0.819 | 0.664 | 0.728 | 0.752 | | | | | | | | |
| Som. (Clark 4S) | 0.753 | 0.164 | 0.084 | 0.993 | 0.708 | 0.13 | 0.977 | 0.695 | 0.824 | 0.192 | 0.77 | 0.68 | 0.685 | 0.149 | | | | | | | |
| Ext. (Achenbach 2S) | -0.154 | -0.453 | -0.311 | -0.17 | -0.307 | -0.356 | -0.12 | -0.307 | -0.19 | -0.392 | -0.299 | -0.428 | -0.389 | -0.436 | -0.141 | | | | | | |
| Rule-Breaking (Achenbach 8S) | -0.137 | -0.305 | -0.112 | -0.094 | -0.162 | -0.168 | -0.056 | -0.201 | -0.112 | -0.264 | -0.175 | -0.22 | -0.208 | -0.237 | -0.058 | 0.593 | | | | | |
| Aggressive (Achenbach 8S) | -0.314 | -0.351 | -0.316 | -0.224 | -0.377 | -0.376 | -0.186 | -0.408 | -0.289 | -0.414 | -0.36 | -0.429 | -0.426 | -0.381 | -0.201 | 0.801 | 0.318 | | | | |
| Ext. (Moore 3S) | -0.181 | -0.395 | -0.278 | -0.145 | -0.275 | -0.311 | -0.095 | -0.289 | -0.173 | -0.363 | -0.252 | -0.367 | -0.339 | -0.367 | -0.107 | 0.926 | 0.646 | 0.829 | | | |
| Ext. (Moore 4S) | -0.237 | -0.397 | -0.251 | -0.23 | -0.317 | -0.29 | -0.182 | -0.331 | -0.231 | -0.366 | -0.3 | -0.397 | -0.373 | -0.349 | -0.192 | 0.922 | 0.638 | 0.834 | 0.994 | | |
| Ext. (McElroy) | -0.118 | -0.369 | -0.249 | -0.135 | -0.237 | -0.274 | -0.085 | -0.239 | -0.149 | -0.317 | -0.225 | -0.357 | -0.33 | -0.352 | -0.104 | 0.935 | 0.614 | 0.841 | 0.95 | 0.94 | |
| Ext. (Deutz GP) | -0.236 | -0.47 | -0.313 | -0.168 | -0.331 | -0.387 | -0.118 | -0.366 | -0.192 | -0.453 | -0.308 | -0.394 | -0.384 | -0.427 | -0.139 | 0.943 | 0.561 | 0.853 | 0.912 | 0.914 | 0.91 |

| | | | | | | | | | | | | | | | | | | | | | | | | | | | | | | | |
|---|---|---|---|---|---|---|---|---|---|---|---|---|---|---|---|---|---|---|---|---|---|---|---|---|---|---|---|---|---|---|---|
| Aggressive (Deutz DP) | -0.1 | -0.296 | -0.198 | -0.17 | -0.226 | -0.211 | -0.132 | -0.216 | -0.151 | -0.256 | -0.217 | -0.372 | -0.326 | -0.296 | -0.148 | 0.839 | 0.371 | 0.888 | 0.858 | 0.85 | 0.919 | 0.796 | | | | | | | | | |
| Ext. (Haltigan GP) | -0.212 | -0.416 | -0.288 | -0.139 | -0.285 | -0.34 | -0.087 | -0.315 | -0.172 | -0.4 | -0.257 | -0.347 | -0.343 | -0.379 | -0.104 | 0.923 | 0.62 | 0.87 | 0.953 | 0.951 | 0.956 | 0.97 | 0.842 | | | | | | | | |
| Ext. (Clark 2S) | -0.252 | -0.314 | -0.281 | -0.107 | -0.266 | -0.322 | -0.07 | -0.296 | -0.186 | -0.337 | -0.231 | -0.22 | -0.292 | -0.314 | -0.081 | 0.717 | 0.567 | 0.749 | 0.813 | 0.816 | 0.789 | 0.808 | 0.649 | 0.856 | | | | | | | |
| Ext. (Clark 3S) | -0.197 | -0.379 | -0.225 | -0.131 | -0.25 | -0.285 | -0.08 | -0.294 | -0.146 | -0.378 | -0.232 | -0.299 | -0.273 | -0.293 | -0.081 | 0.875 | 0.656 | 0.807 | 0.953 | 0.951 | 0.886 | 0.898 | 0.776 | 0.932 | 0.818 | | | | | | |
| Ext. (Clark 4S) | -0.298 | -0.398 | -0.281 | -0.243 | -0.352 | -0.336 | -0.19 | -0.387 | -0.258 | -0.399 | -0.338 | -0.396 | -0.385 | -0.33 | -0.195 | 0.893 | 0.638 | 0.827 | 0.953 | 0.961 | 0.888 | 0.916 | 0.78 | 0.937 | 0.825 | 0.979 | | | | | |
| Att. (Achenbach 8S) | 0.023 | -0.164 | -0.018 | -0.102 | -0.115 | -0.08 | -0.1 | -0.13 | -0.029 | -0.146 | -0.158 | -0.281 | -0.17 | -0.146 | -0.1 | 0.1 | -0.025 | -0.039 | -0.054 | -0.063 | -0.024 | 0.038 | 0.036 | -0.087 | -0.262 | -0.061 | -0.049 | | | | |
| Att. (Moore 3S) | 0.155 | -0.141 | 0.017 | -0.024 | -0.011 | -0.008 | -0.018 | 0.000 | 0.058 | -0.064 | -0.057 | -0.214 | -0.085 | -0.116 | -0.022 | 0.115 | -0.022 | -0.095 | -0.04 | -0.059 | 0.016 | 0.005 | 0.077 | -0.097 | -0.306 | -0.093 | -0.094 | 0.937 | | | |
| Att. (Moore 4S) | 0.101 | -0.153 | 0.038 | -0.106 | -0.058 | 0.005 | -0.101 | -0.049 | 0.000 | -0.078 | -0.109 | -0.258 | -0.129 | -0.111 | -0.104 | 0.128 | -0.012 | -0.074 | -0.024 | -0.034 | 0.028 | 0.02 | 0.093 | -0.082 | -0.293 | -0.076 | -0.07 | 0.94 | 0.996 | | |
| Att. (McElroy) | 0.173 | -0.129 | 0.043 | -0.016 | 0.007 | 0.011 | -0.011 | 0.01 | 0.077 | -0.055 | -0.045 | -0.188 | -0.071 | -0.094 | -0.016 | 0.113 | -0.016 | -0.104 | -0.039 | -0.057 | 0.007 | 0.002 | 0.07 | -0.107 | -0.289 | -0.086 | -0.086 | 0.903 | 0.93 | 0.925 | |
| Att. (Deutz DP) | 0.22 | -0.067 | 0.132 | -0.021 | 0.074 | 0.115 | -0.024 | 0.082 | 0.116 | 0.031 | 0.013 | -0.108 | 0.009 | -0.004 | -0.023 | 0.056 | -0.028 | -0.193 | -0.1 | -0.111 | -0.055 | -0.058 | 0.003 | -0.169 | -0.331 | -0.143 | -0.144 | 0.87 | 0.911 | 0.909 | 0.982 |

| | | | | | | | | | | | | | | | | | | | | | | | | | | | | | | | | | | | | | |
|---|---|---|---|---|---|---|---|---|---|---|---|---|---|---|---|---|---|---|---|---|---|---|---|---|---|---|---|---|---|---|---|---|---|---|---|---|---|
| Att. (Haltigan GP) | 0.082 | -0.207 | -0.014 | -0.041 | -0.071 | -0.078 | -0.029 | -0.093 | 0.039 | -0.165 | -0.109 | -0.248 | -0.14 | -0.163 | -0.034 | 0.165 | 0.027 | -0.016 | 0.03 | 0.014 | 0.049 | 0.103 | 0.098 | -0.016 | -0.204 | 0.005 | 0.006 | 0.921 | 0.904 | 0.903 | 0.976 | 0.939 | | | | | |
| Att. (Clark 3S) | 0.149 | -0.121 | 0.063 | -0.028 | 0.006 | 0.022 | -0.022 | -0.001 | 0.074 | -0.064 | -0.044 | -0.17 | -0.038 | -0.055 | -0.02 | 0.067 | -0.052 | -0.121 | -0.094 | -0.109 | -0.042 | -0.02 | 0.014 | -0.132 | -0.322 | -0.077 | -0.098 | 0.93 | 0.957 | 0.954 | 0.893 | 0.876 | 0.88 | | | | |
| Att. (Clark 4S) | 0.019 | -0.183 | -0.026 | -0.143 | -0.131 | -0.077 | -0.13 | -0.139 | -0.053 | -0.141 | -0.179 | -0.305 | -0.189 | -0.133 | -0.13 | 0.162 | 0.01 | -0.017 | -0.003 | -0.011 | 0.043 | 0.081 | 0.093 | -0.038 | -0.234 | 0.002 | 0.015 | 0.961 | 0.96 | 0.965 | 0.896 | 0.865 | 0.901 | 0.966 | | | |
| Social (Achenbach 8S) | 0.028 | -0.034 | 0.053 | -0.052 | 0.019 | 0.059 | -0.029 | 0.023 | 0.028 | 0.012 | 0.025 | -0.033 | -0.07 | 0.016 | -0.053 | 0.066 | 0.035 | 0.035 | 0.115 | 0.118 | 0.111 | 0.07 | 0.123 | 0.092 | 0.02 | 0.023 | 0.097 | -0.049 | -0.052 | -0.045 | -0.002 | -0.006 | 0.001 | -0.14 | -0.077 | | |
| Tho. (Achenbach 8S) | 0.159 | 0.07 | 0.046 | 0.013 | 0.068 | 0.078 | 0.001 | 0.143 | -0.088 | 0.155 | -0.006 | -0.032 | 0.023 | -0.028 | 0.006 | -0.029 | -0.074 | -0.146 | -0.127 | -0.136 | -0.022 | -0.258 | 0.006 | -0.209 | -0.219 | -0.233 | -0.212 | 0.042 | 0.161 | 0.152 | 0.162 | 0.18 | 0.003 | 0.092 | 0.06 | -0.065 | |
| Tho. (Haltigan GP) | 0.218 | 0.03 | 0.059 | 0.097 | 0.128 | 0.09 | 0.093 | 0.189 | 0.003 | 0.127 | 0.077 | 0.017 | 0.094 | -0.038 | 0.1 | 0.01 | -0.03 | -0.124 | -0.058 | -0.074 | 0.032 | -0.199 | 0.049 | -0.128 | -0.169 | -0.162 | -0.182 | 0.006 | 0.128 | 0.114 | 0.136 | 0.146 | 0.005 | 0.052 | 0.003 | -0.019 | 0.874 |

Note: Int., Internalizing; Ext., Externalizing; Som., Somatic; -dep., Depression; Att., Attention; Tho., Thought; nS, number of specific factors in the model; GP, General psychopathology model; DP, dysregulation profile model.
